# Multimodal approach to identify neuropsychophysiological subgroups in myalgic encephalomyelitis/chronic fatigue syndrome and their relevance for rehabilitation: protocol for a mechanistic cross-sectional and longitudinal study

**DOI:** 10.64898/2026.06.05.26354983

**Authors:** Ynse Dooms, Lixin Qiu, Iris Coppieters, Elfi Vergaelen, Stephan Claes, Patrick Dupont, Melina Hehl, Koen Cuypers, Harald Engler, Kirsten Dombrowski, Kristin Verbeke, Omer Van den Bergh, Jeroen Raes, Lukas Van Oudenhove, Maaike Van Den Houte, Katleen Bogaerts

**Author notes:** Corresponding author: Maaike Van den Houte, ON2 Herestraat 49 - bus 70, KU Leuven, 3000 Leuven, Belgium. Co-senior authorship (Lukas Van Oudenhove, Maaike Van Den Houte, Katleen Bogaerts). **Contact details:** Ynse Dooms, Lixin Qiu, Iris Coppieters, Elfi Vergaelen, Stephan Claes, Patrick Dupont, Melina Hehl, Koen Cuypers, Harald Engler, Kirsten Dombrowski, Kristin Verbeke, Omer Van den Bergh, Jeroen Raes, Katleen Bogaerts, Lukas Van Oudenhove, Maaike Van Den Houte.

## Abstract

**Introduction:** Myalgic Encephalomyelitis (ME)/Chronic Fatigue Syndrome (CFS) is a debilitating condition characterised by severe fatigue and post-exertional malaise (PEM). Reported neuropsychophysiological abnormalities suggest ME/CFS is multifactorial, but current knowledge remains fragmented. This study protocol outlines a multimodal investigation designed to (1) compare neuropsychophysiological mechanisms between ME/CFS patients and healthy participants, (2) test an integrative model of ME/CFS, (3) identify neuropsychophysiological subgroups within the patient population, and (4) identify predictors of symptom response during rehabilitation.

**Methods and analysis:** This study will enroll 115 ME/CFS patients and 55 healthy participants. Groups will be comparable in age, sex, and education level, with a larger patient sample enabling subgroup and longitudinal analyses. A cross-sectional assessment at baseline will be carried out in both groups. Patients will then be evaluated longitudinally throughout a standardized cognitive-behavioral therapy rehabilitation program delivered as routine care. Baseline measures include systemic inflammation and general health biomarkers, measures of autonomic and central nervous system function, neuroinflammation (magnetic resonance spectroscopy, [^18^F]DPA714 PET in a subsample), serum short-chain fatty acid levels, gut microbiota composition and function, and neuroendocrine and self-reported responses to psychosocial stress. Fatigue severity (physical and cognitive) and PEM will be assessed through validated questionnaires, ecological momentary assessment, and laboratory tasks. These will be re-evaluated during therapy, and all non-neuroimaging measures will be repeated after the rehabilitation program. Statistical analyses will comprise multivariate analysis of variance, general linear models, classification algorithms, structural equation models, least absolute shrinkage selection operator principal component regression (LASSO-PCR), cluster analysis and latent class growth analysis (LCGA).

## 1. Introduction

Myalgic Encephalomyelitis/Chronic Fatigue Syndrome (ME/CFS) is a complex and debilitating condition predominantly defined by persistent, intense, and impairing fatigue and by post-exertional malaise (PEM), a worsening of symptoms following physical or cognitive exertion (16,141, 170). This fatigue cannot be alleviated by rest and limits daily activities significantly (16, 32). Besides fatigue, patients often experience various other symptoms, such as cognitive problems, orthostatic intolerance, muscle or joint pain, headaches, and unrefreshing sleep (32). Several sets of diagnostic criteria exist (16, 32). Although the prognosis for ME/CFS varies widely, only less than 10% of cases return to the same levels of functioning as before disease onset without treatment (3). Therapies focusing primarily on symptom management, such as cognitive behavioral therapy (CBT) and energy management, have shown effectiveness in improving wellbeing and quality of life (1, 6, 47). Analgesics and other pharmacological treatments are also frequently used for symptomatic relief (13, 23).

Although the precise etiology of ME/CFS remains unclear, ME/CFS is believed to have a multifactorial origin (2, 30), where underlying mechanisms are suggested to interact in a self-intensifying “vicious cycle” disrupting the body’s homeostasis, resulting in deterioration of symptoms over time (71).

The resemblance of ME/CFS symptoms to inflammation-induced sickness behaviors, such as fatigue, altered sleep and malaise, has sparked interest in the role of chronic low-grade **inflammation** in ME/CFS (63). Large-scale case-control studies, systematic reviews, and a meta-analysis on cytokine profiles reported significantly increased levels of CRP, IL-1, IL-4, IL-6, IL-8, IL-10, IL-12, TGF-β, TNF-β, and TNF-α in ME/CFS compared to healthy participants (18, 40, 65, 76, 79, 161, 166, 167). At the same time, several findings have shown decreased levels of IL-1β, IL-6, IL-8, IL-10, IL-17A, and IFN-γ (18, 41, 79, 168). However, other recent studies have failed to consistently replicate these significant differences (73, 162, 169), pointing to considerable divergence in the literature and suggesting that immune dysregulation in ME/CFS may be more complex. For instance, recent studies investigating cellular immunity in ME/CFS reported natural killer (NK) cell dysfunction (40, 154, 155), including decreased NK cell cytotoxicity (66), lower antibody-dependent cell-mediated cytotoxicity (156), distinct T-cell phenotypes (160–162), and T-cell exhaustion (36, 164).

Tilt-table testing, sympathetic skin response tests, heart rate variability (HRV), and cortisol measurements suggest malfunctions in the **neuroendocrine stress response system** (SRS) in ME/CFS patients (43, 51, 52, 61). During stress, the hypothalamic-pituitary-adrenal (HPA)-axis and the autonomic nervous system (ANS) are activated to preserve homeostasis (163). The majority of the studies showed sympathetic nervous system (SNS) predominance in ME/CFS patients compared to healthy participants (43, 51, 61), implying a lack of adaptability to changing environmental demands (60). The preponderance of studies reports that ME/CFS is marked by lower morning cortisol levels, suggesting hypocortisolism and HPA-axis downregulation (52). Additional evidence includes a blunted cortisol response to physical and psychological challenges in some, but not all, studies (52).

The evidence for chronic low-grade systemic inflammation, along with the prominent neuropsychological symptoms observed in ME/CFS patients, suggests a potential role for **neuroinflammation** in the pathogenesis and persistence of the symptoms, hence the term myalgic encephalomyelitis/chronic fatigue syndrome (ME/CFS) (99, 165). However, direct evidence for this hypothesis remains sparse (99, 165). Currently, the gold standard for quantifying neuroinflammation in vivo in humans is positron emission tomography (PET) targeting the 18-kDa translocator protein (TSPO) (99). Increases in TSPO expression in the brain are indicative of increased density of microglia and astrocytes, the primary inflammatory cells in the brain (12, 81). A small case-control study reported increased neuroinflammation in the cingulate cortex, hippocampus, amygdala, thalamus, midbrain, and pons in ME/CFS patients (72). However, a more recent, but equally small, study using gold-standard arterial input-based kinetic modeling to quantify the PET signal found no evidence of altered TSPO binding in patients (82). Both studies used PK11195, a first-generation TSPO radiotracer which has a poor blood-brain barrier permeability and high non-specific binding (62, 99). To more robustly evaluate neuroinflammation in ME/CFS, there is a need for studies using second-generation TSPO tracers and substantially larger samples (165). A less direct but less costly and invasive way to study neuroinflammation is via magnetic resonance spectroscopy (MRS) (159). Single-voxel and whole-brain MRS studies discovered metabolite abnormalities in the brains of individuals with ME/CFS (99). Significant group differences for choline (CHO) and myo-inositol (MI) in the anterior cingulate cortex (ACC) suggested neuroinflammation and glial dysfunction in ME/CFS patients (33, 67).

Beyond neuroinflammation, a range of other **central nervous system** changes have been investigated using MRI (93, 165). The majority of the studies investigating blood oxygenation level-dependent (BOLD) signals described that patients with ME/CFS had a stronger BOLD response or engaged additional brain regions during cognitive tasks as opposed to healthy participants, despite having comparable cognitive performances (85, 93). This may reflect the increased need of neural resources for a given cognitive effort, leading to elevated fatigue, which is demonstrated by the significant associations between increased brain activity and fatigue during cognitive tasks in ME/CFS patients (17). This result is reinforced by absence of BOLD adaptation, potentially reflecting energy efficiency, in patients with ME/CFS during sustained cognitive tasks (93). Further, functional connectivity findings demonstrate alterations across a number of resting-state networks, including the default mode network and salience network (87, 92). Scores of self-reported fatigue are found to be associated with altered functional connectivity patterns (69, 92).

Alongside these observations in the central nervous system, a growing body of evidence points to the **gut microbiome** dysbiosis in ME/CFS (158). Microorganisms and their metabolites prevent (bacterial) antigens from entering the system where they may drive immune and metabolic reactions (91). Alterations in ME/CFS patients included reduced α-diversity and β-diversity as well as a diminished Firmicutes/Bacteroidetes ratio (102). This microbiome dysbiosis may cause a reduced production of beneficial microbial metabolites like short-chain fatty acids (SCFA) (157). Decreased SCFA levels are suggested to impact cognitive function, fatigue severity, impaired stress resilience, depressive feelings, and unrefreshing sleep via the gut-brain axis (21), highlighting their possible involvement in ME/CFS.

While this body of work in different neuropsychophysiological domains has substantially advanced our understanding of ME/CFS, the current evidence is constrained by several important limitations.

*First*, disabling fatigue and PEM, though a defining feature in all ME/CFS patients, are subjective and ambiguous, and their perception varies widely across patients (70). Therefore, the concept of fatigue cannot be captured by unidimensional scales, underscoring the need for multidimensional assessment, covering both physical and cognitive dimensions of fatigue (80, 84).

*Second*, despite the evidence that multiple neuropsychophysiological mechanisms may contribute to the pathophysiology of ME/CFS, the interactions between these mechanisms and how they relate to the symptom experience are understudied. For instance, the bidirectional interaction between the immune pathway and the HPA-axis (94), the causal cascades of peripheral inflammation to neuroinflammation and their effect on behavioral symptoms (97), the link between fatigue sensations, sympathetic hyperactivity and diminished parasympathetic activity (64), and the gut-brain communication via bidirectional signalling mechanisms, with the mediating function of microbial metabolites on cognition, fatigue severity, and depressive symptoms (21, 95) have been shown in healthy participants but not yet in ME/CFS. Therefore, an integrative, multimodal approach, measuring neuropsychophysiological functioning in different bodily systems in the same set of patients, is needed.

*Third*, existing studies have concentrated primarily on case-control differences despite the fact that ME/CFS is known to be a heterogeneous disorder, possibly contributing to the contradictory findings in the literature (142). Therefore, the identification of inter-individual differences within the ME/CFS patient group and delineating mechanistic subgroups within the ME/CFS population is a critical priority (142).

*Fourth*, although it is often assumed that neuropsychophysiological dysfunctions precede the development or exacerbation of ME/CFS symptoms, the temporal direction of these relationships remains unclear due to a lack of longitudinal studies (134, 142).

This paper describes the protocol of a comprehensive, multimodal study designed to investigate the central, systemic, and peripheral neuropsychophysiological mechanisms underlying ME/CFS, and their interactions. Given the disabling fatigue and disruptions reported among the immune, (neuro)endocrine, autonomic, gastrointestinal, and neural function in ME/CFS (51, 52, 63, 93, 99, 158), the first objective of our study is to compare these mechanisms between a well-characterized, large sample of ME/CFS patients and healthy participants. Secondly, we want to test the interactions between the stress response system, the immune system, the brain, and the gut microbiota and their metabolites in ME/CFS patients, as such interactions have been shown in healthy participants (21, 64, 94, 95). Third, we want to identify subgroups of ME/CFS patients based on inter-individual differences in pathophysiological characteristics, including multidimensional measurement of fatigue and fatigability severity (142). Lastly, we want to investigate to what extent the pathophysiological profile at baseline predicts response to a standardized ME/CFS-specific CBT-based rehabilitation program, and to investigate to what extent this pathophysiological profile is changed after treatment, in line with previous outcomes linking physiological characteristics to symptom severity (65, 69, 72). See **Table 1** and **Supplementary Table 1** for an overview of objectives, research questions, measured variables, and planned analysis methods.

**Table 1.**
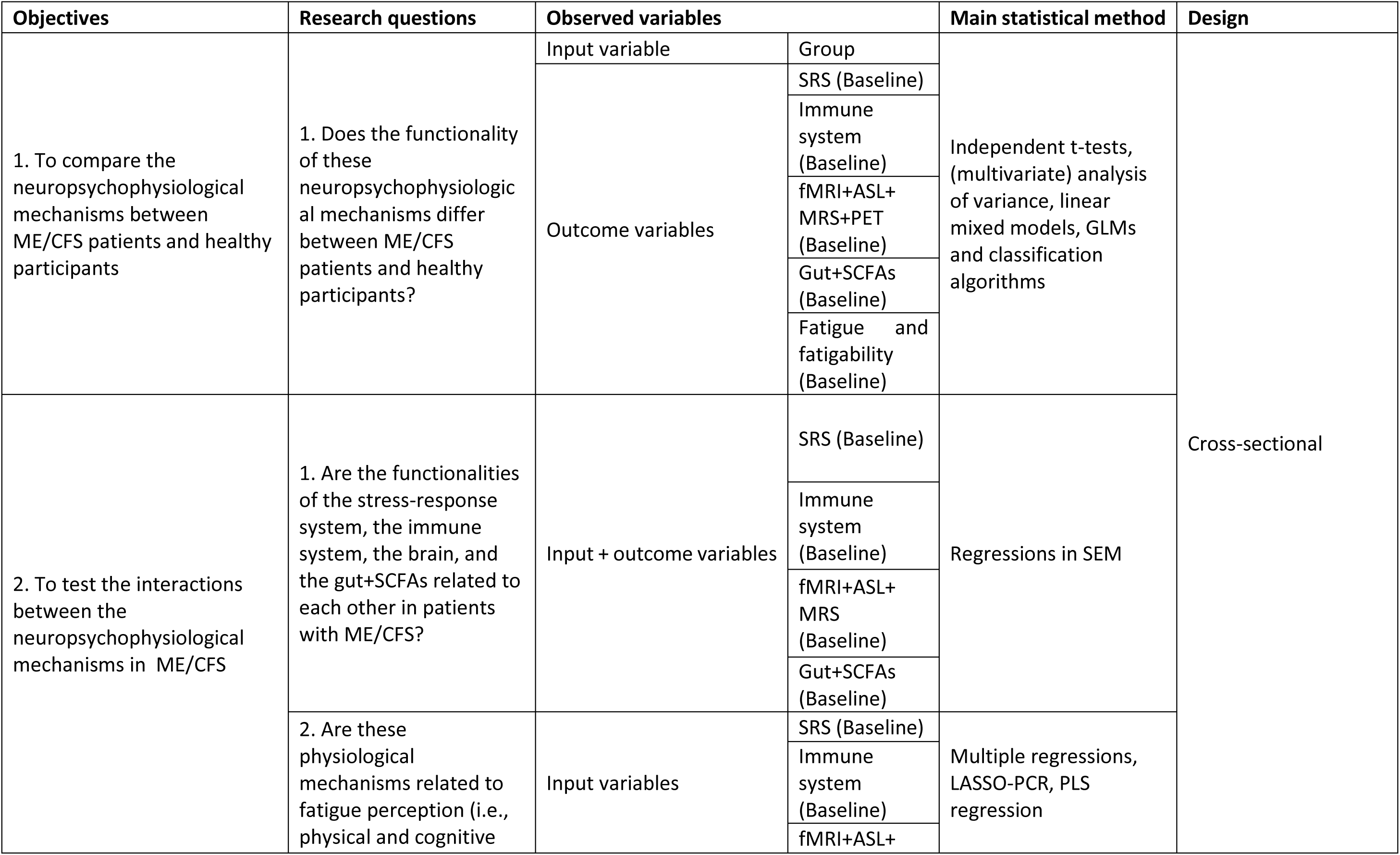
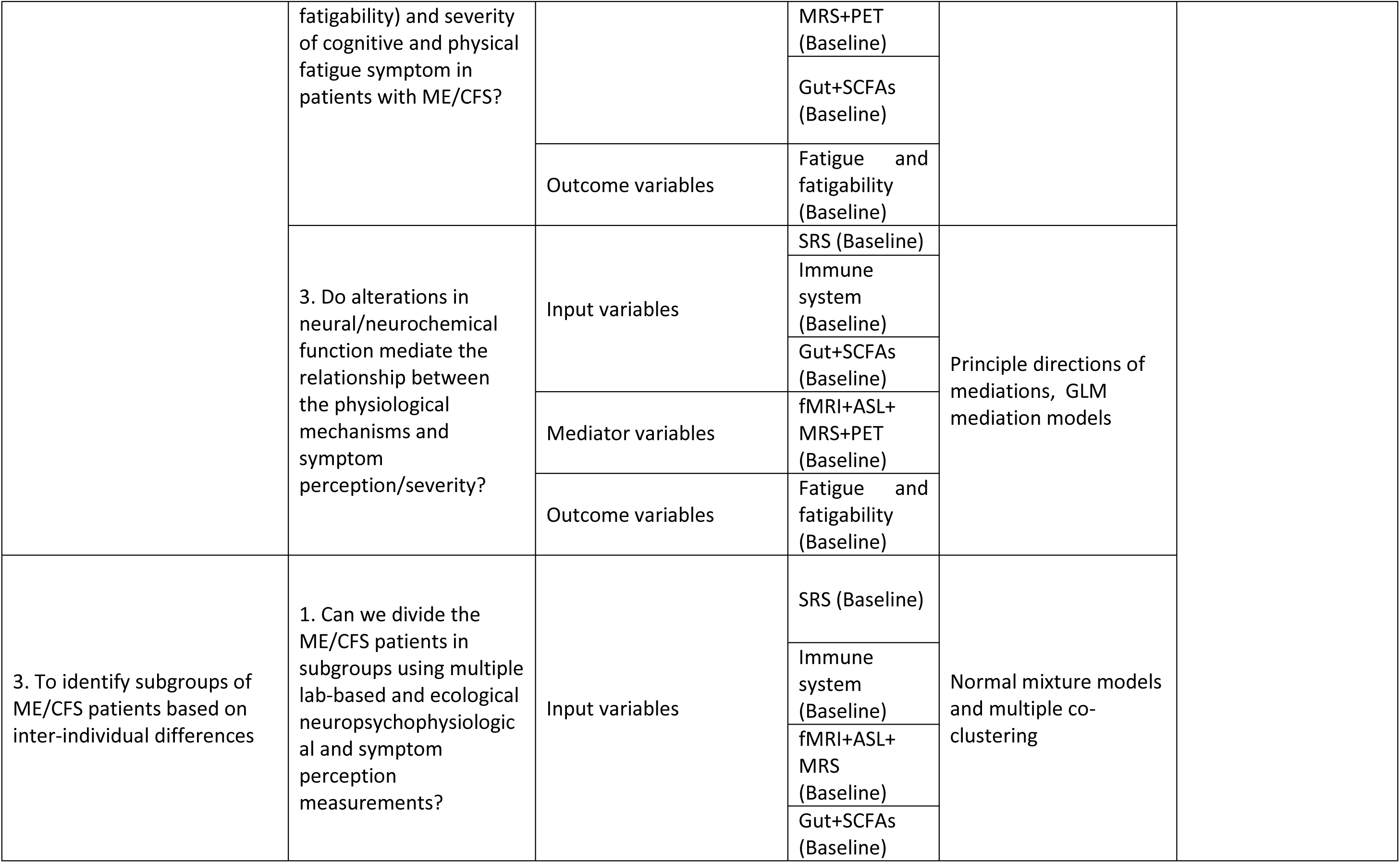
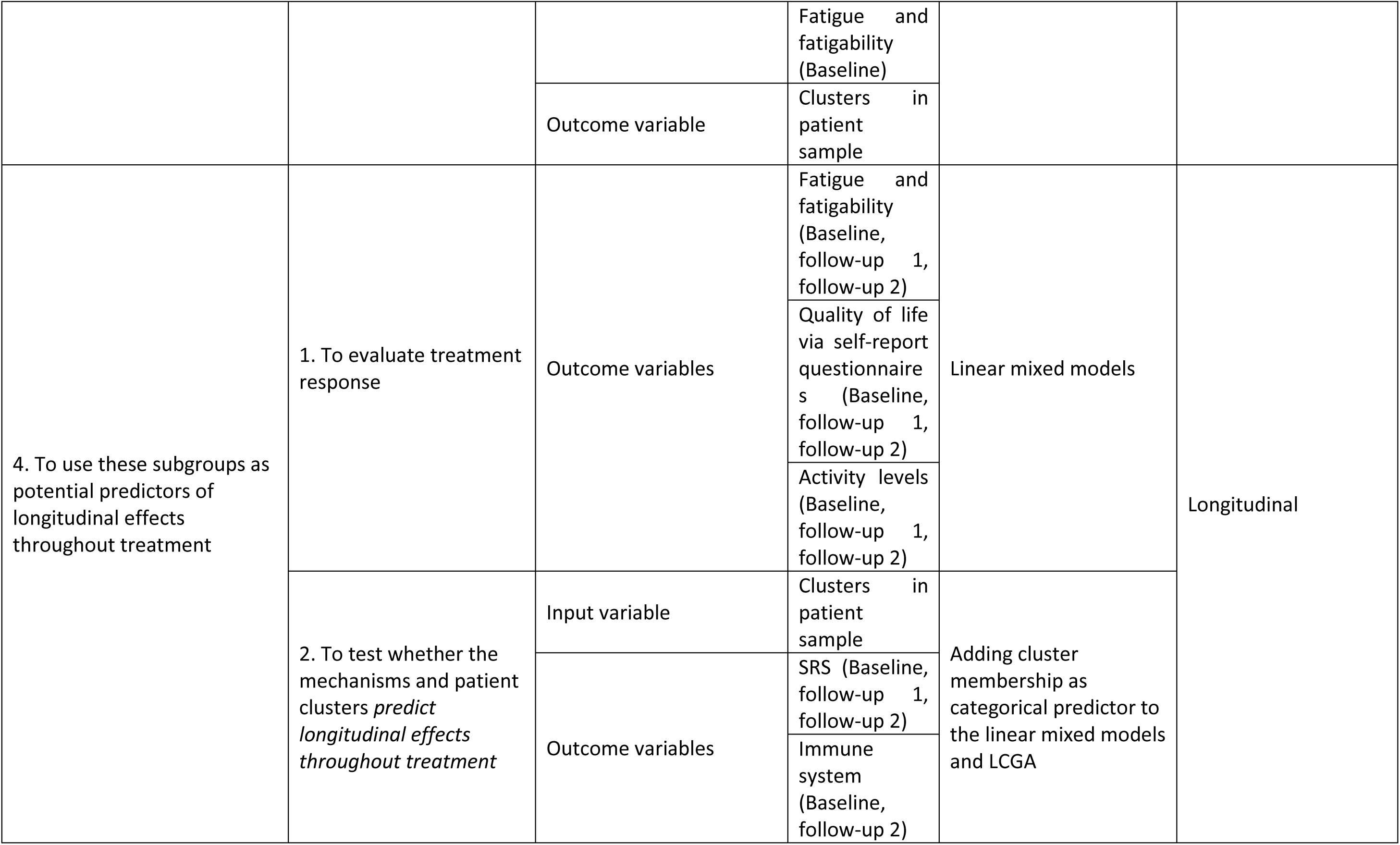
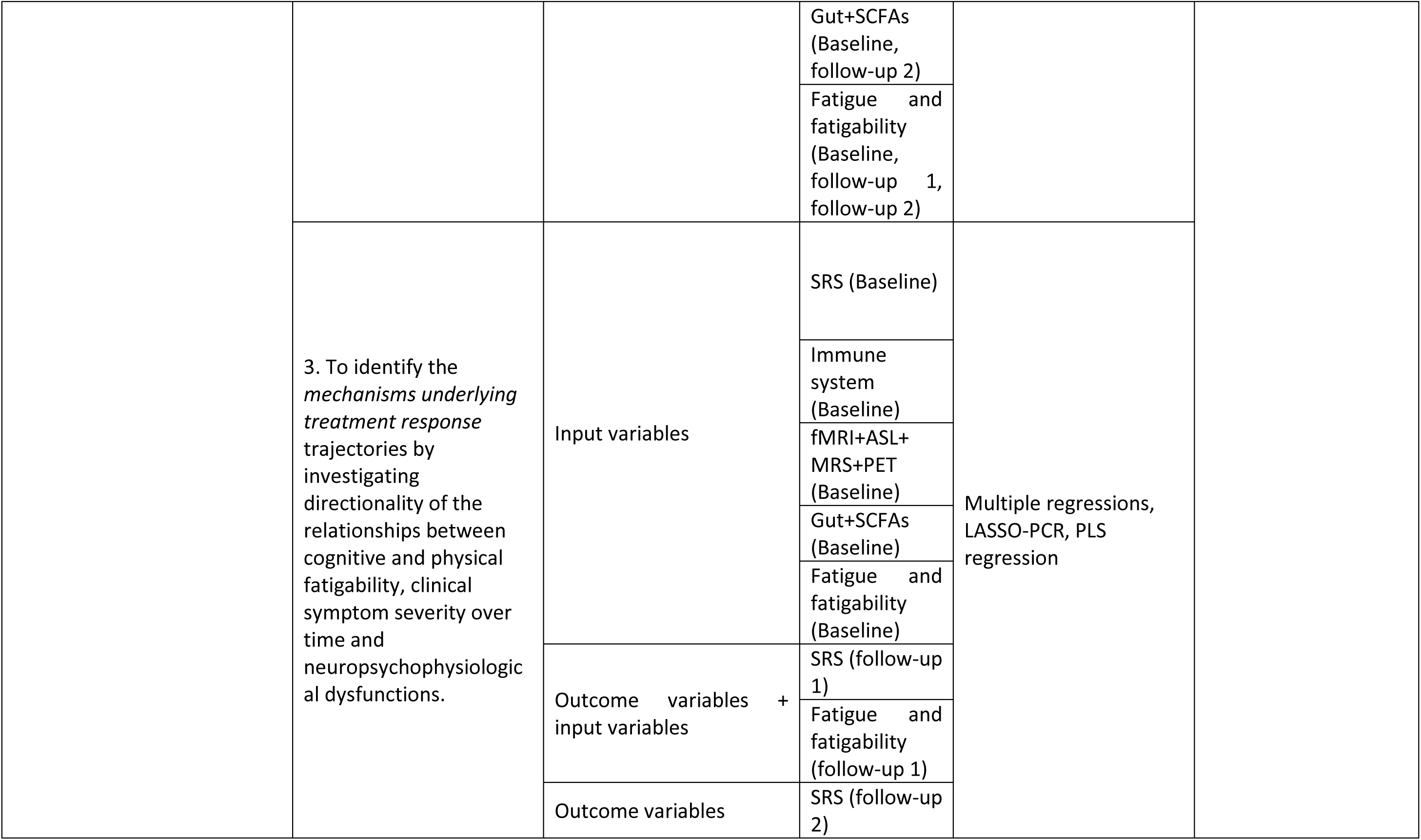
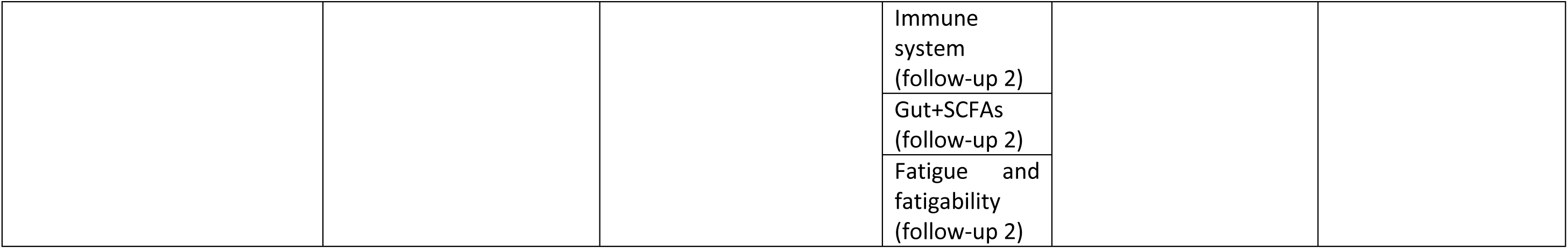
ME/CFS= myalgic encephalomyelitis/chronic fatigue syndrome, SRS= stress response system, SCFA= short chain fatty acids, GLM=general linear model, SEM=structural equation model, LASSO-PCR= Least Absolute Shrinkage and Selection Operator-Regularized Principal Component Regression, LCGA= Latent class growth analysis, PLS= Partial Least Squares

## 2. Methods

### 2.1 Experimental design

This mechanistic study combines a cross-sectional case-control part *(“baseline”)* and a longitudinal part in which patients are followed-up during and after routine care (i.e. a standardized CBT rehabilitation program) *(“follow-up”)* **(Figure 1)**. The first three objectives will be addressed using baseline data, while the fourth objective will be addressed using baseline and follow-up data. Patients with ME/CFS and healthy participants will be first assessed at baseline throughout a clinical assessment visit (T1), a brain imaging visit (T2), a lab-based testing moment (T3) and home assessments (**Figure 1**). Patients will then be followed throughout standardized clinical care, a CBT rehabilitation program. Follow-up measurements will take place after ten and fifteen therapy sessions. During both follow-up moments, patients will again perform the home assessments. After fifteen sessions, a lab-based testing moment (T4), similar to T3, will be performed (**Figure 2**). To minimize the effects of the circadian rhythm on the neuro-endocrine stress response system and to standardize blood collection, T3 and T4 will always take place in the afternoon (between 12:00 and 18:00). Participants will be instructed to abstain from alcohol consumption and strenuous exercise for 24 hours prior to each test session and ensure adequate rest the night before. They will also be asked to avoid caffeine and smoking in the morning of T3 and T4 and to not brush their teeth or chew gum within two hours of its start. All measurements will be conducted at University Hospitals Leuven (Leuven, Belgium) or at the participant’s home. Ethical approval was obtained from the Ethical Committee Research UZ/KU Leuven (Ref. S66452). Data collection started in February 2023 and is expected to finish in December 2026.

**Figure 1.**
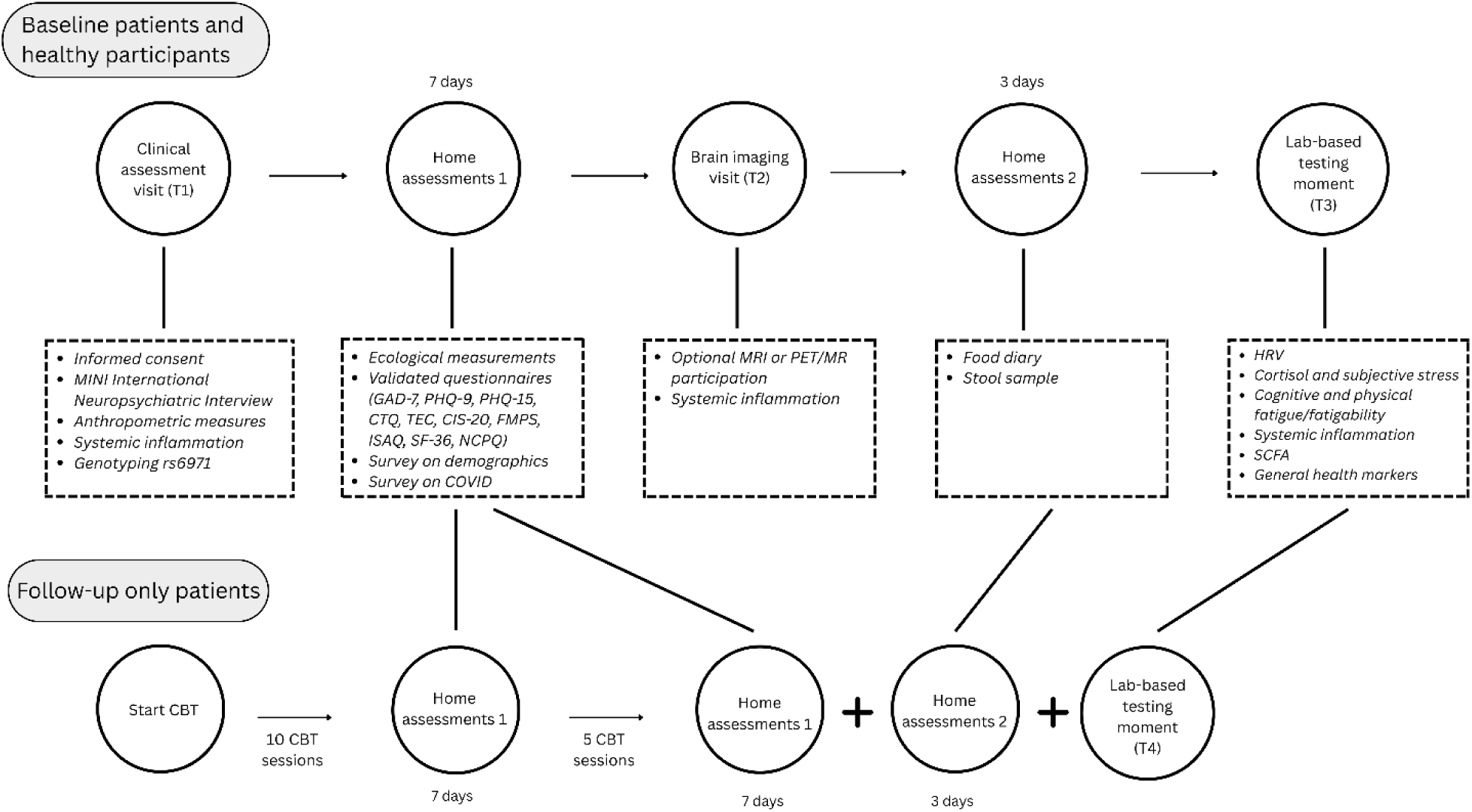
Overview of the cross-sectional and longitudinal parts of the study design. MRI=magnetic resonance imaging, PET/MR= positron emission tomography/magnetic resonance, HRV= heart rate variability, SCFA=short chain fatty acids, CBT=cognitive behavioral therapy, GAD-7=Generalized Anxiety Disorder-7, PHQ-9= Patient Health Questionnaire-9, PHQ-15=Patient Health Questionnaire-15, CTQ=Childhood Trauma Questionnaire, TEC=Traumatic Experience Checklist, CIS-20=Checklist Individual Strength-20, FMPS=Frost Multidimensional Perfectionism Scale, ISAQ= Interoceptive Sensitivity and Attention Questionnaire, SF-36=36-Item Short Form Health Survey, NCP-Q= Need for Controllability and Predictability questionnaire.

**Figure 2.**
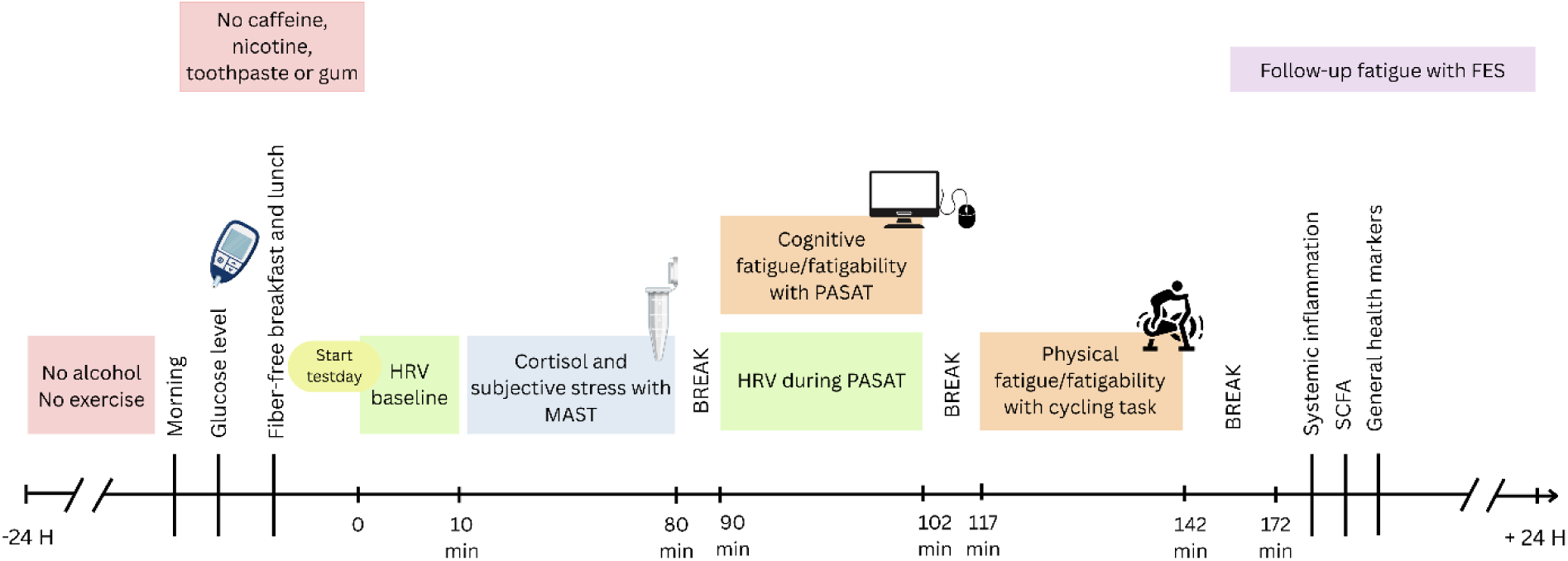
Outline of the lab-based testing moments (T3 and T4). HRV= heart rate variability, MAST= Maastricht Acute Stress Task, PASAT= Paced Auditory Serial Attention Task, SCFA= short-chain fatty acids, FES=fatigue and energy scale.

### 2.2 Standardized rehabilitation program

The CBT rehabilitation program is part of the standard clinical care of CFS patients in Belgium delivered by licensed cognitive behavioral therapists according to the [program guidelines](https://www.riziv.fgov.be/SiteCollectionDocuments/cognitief_gedragstherapeut_cvs_therapieprotocol.pdf). It is reimbursed and bound by convention by the Belgian National Institute for Health and Invalidity Insurance (RIZIV/INAMI). More details on the program are in Supplement.

### 2.3 Participants

We will recruit 115 patients diagnosed with ME/CFS after a comprehensive assessment by internists and psychiatrists with specialized expertise in fatigue evaluation. Patients will be recruited in the Multidisciplinary Diagnostic Center for ME/CFS of the University Hospitals Leuven, the only multidisciplinary center for ME/CFS recognized by the Belgian National Institute for Health and Invalidity Insurance (RIZIV/INAMI). Until February 29th, 2024, the Multidisciplinary Diagnostic Center employed the 1994 Center of Disease Control criteria (32) for diagnosis; from March 1st, 2024, onwards, the NICE criteria (34) are employed. Patients suffering from major depressive disorder or patients reporting pain, rather than fatigue, as their primary symptom, do not receive the ME/CFS diagnosis. Patients diagnosed outside the Multidisciplinary Diagnostic Center will be considered for inclusion only if their diagnostic procedure is comparable to that employed at the Multidisciplinary Diagnostic Center, and provided that at least 10 years have passed since the completion of their standard clinical care program for ME/CFS. They will not be included in the longitudinal study. Fifty-five healthy controls (HC) will be recruited via local outreach. HC will be selected in a manner that ensures their sex, age, and educational degree distributions are comparable to those of the patients. Details about the exclusion criteria can be found in Supplement.

### 2.4 Sample size

The aspired participant numbers reflect expected feasibility based on patient flow in the hospital’s Multidisciplinary Diagnostic Center and the available budget. The sample size represents the intended number of participants completing baseline measurements. In case of dropouts, additional participants will be recruited to achieve the targeted sample size. The aimed sample size yields 80% power to detect group differences in physiological, neurological and symptom parameters, and to detect 4 equally sized clusters (44). The sample size is also sufficient to perform latent class growth analysis (LCGA) (44), and least absolute shrinkage selection operator principal component regression (LASSO-PCR) (50). More information on power calculations is in Supplement.

### 2.5 Measures

#### 2.5.1 Primary outcome variables

##### 2.5.1.1 Systemic inflammation

To assess systemic inflammation, 15 mL venous blood will be drawn from the participant’s antecubital vein into serum-separator tubes (Vacutainers; Becton-Dickinson, Franklin Lakes, NJ). The serum will be separated and stored at −80°C until analysis. Serum levels of TNF-α, IL-1β, IL-4, IL-6, IL-8, IL-10 and IFN-γ will be quantified by multiplex electrochemiluminescence immunoassay using the V-PLEX Human Proinflammatory Panel 1 kit, and TGF-β will be measured using the U-PLEX Human TGF-β1 Assay, both from Meso Scale Diagnostics (Rockville, MD), following the manufacturer’s instructions. Analyses will be performed at the Institute of Medical Psychology and Behavioral Immunobiology (University Hospital Essen, Germany). High sensitivity CRP (hs-CRP) will be measured using a latex-enhanced immunoturbidimetry test (COBAS 8000, Hitachi, Tokyo, Japan/Roche Diagnostics, IN, USA) at the Laboratory Medicine Department (UZ Leuven). Details on used assays are in Supplement.

##### 2.5.1.2 General health biomarkers

A 4 mL blood sample will be collected from the participant’s antecubital vein into EDTA tubes (Vacutainers, Becton-Dickinson, Franklin Lakes, NJ) for the assessment of complete blood counts, a lipid profile, inflammatory markers and a metabolic, liver, and renal panel. An overview of general health biomarkers can be found in Supplement. Blood analyses will be performed immediately upon collection at the Laboratory Medicine Department (UZ Leuven). Additionally, fasting blood glucose will be determined with a glucometer (GlucoMen areo, Menarini Diagnostics, Florence, Italia).

##### 2.5.1.3 Heart rate (HR) and heart rate variability (HRV)

Electrocardiogram (ECG) recordings will be made with a two-channel ECG monitor (Firstbeat Bodyguard 3, Firstbeat Technologies Oy, Jyväskylä, Finland). Electrodes (Kendall electrodes, Medi-Trace, Belgium) will be positioned similar to RA and LL positions. Patients will be monitored for ten consecutive minutes in rest and during a mental arithmetic task (PASAT, see “2.5.1.7 Fatigue and fatigability lab measures”). Extracted interbeat intervals will be visually inspected and processed offline using ARTiiFACT software (74). HR and time domain indices of HRV will be calculated by assessing the root mean squares of successive RR interval differences (RMSSD). RMSSD is the primary time-domain measure in research as it provides a highly relevant and accurate measure of short-term autonomic nervous system activity (86, 101, 103).

##### 2.5.1.4 Gut microbiota composition

Participants will be instructed to collect a stool sample at home and to determine stool consistency by means of the provided Bristol Stool Score chart (96, 109). Participants will be instructed to store the sample in their −18°C home freezer until delivery to the lab. Cooling elements and cooler bags will be used for transport. After receipt, fecal samples will be stored at −80°C. Fecal samples will be analysed via 1) shotgun metagenomic sequencing/analysis, transcriptomic analysis, metabolomic analysis, 16S/18S/ITS amplicon sequencing analysis, 2) isolation and cultivating gut microorganisms and 3) bacterial cell count using flow cytometry after dilution of the stool samples aliquots. Additionally, fecal calprotectin will be analysed as a measure of gut-inflammation via ELISA as well as moisture content (a proxy for transit time) through lyophilizing the aliquots. Analyses will be performed by the Raes lab (VIB-KU Leuven Center for Microbiology, Leuven, Belgium).

##### 2.5.1.5 Short chain fatty acids (SCFAs)

To assess SCFAs levels, 4 ml venous blood will be collected in serum-separator tubes (Vacutainers, Becton-Dickinson, Franklin Lakes, NJ). The serum will be separated and stored at −80°C until analysis. Acetate, butyrate and propionate concentrations will be determined using gas chromatography-mass spectrometry (GC-MS) (Trace 1300 GC and DSQ II XL, Thermo Electron Corporation, Waltham, MA, USA) following derivatization with 2,4-difluoroaniline and extraction in ethyl acetate (98). Details in Supplement.

##### 2.5.1.6 Salivary cortisol response to psychosocial stress

The Maastricht Acute Stress Test (MAST) (88) includes a 5-min instruction phase, a 10-min stress induction phase, and a 55-minute recovery period (**Figure 3**). The stress induction phase consists of alternations between cold water hand immersion trials (6°C) and mental arithmetic trials, with trial durations varying between 45 and 90 s. Social stressors are added by giving negative feedback after mistakes and mock videotaping. The MAST elicits a robust physiological and subjective stress response (88). For the assessment of the salivary cortisol response, saliva samples will be collected with Salivette Cortisol (Sarstedt AG & Co., Nümbrecht, Germany) before the instruction phase and seven times after the stress induction phase (t+00, t+05, t+15, t+25, t+35, t+45, t+55) (**Figure 3**). Samples will be centrifuged immediately after collection and stored at −20°C until enzyme-linked immunosorbent assay (ELISA) analysis (TECAN, RE52611) (assay details in Supplement) at the Institute of Medical Psychology and Behavioral Immunobiology (University Hospital Essen, Germany) according to the manufacturer’s guidelines. Participants will rate subjective stress at baseline, during instructions, and in the middle of and after the stress induction phase by scoring the stressfulness, painfulness and unpleasantness of the test on a visual analogue scale (VAS) ranging from 0: not at all, to 10: extremely.

**Figure 3.**
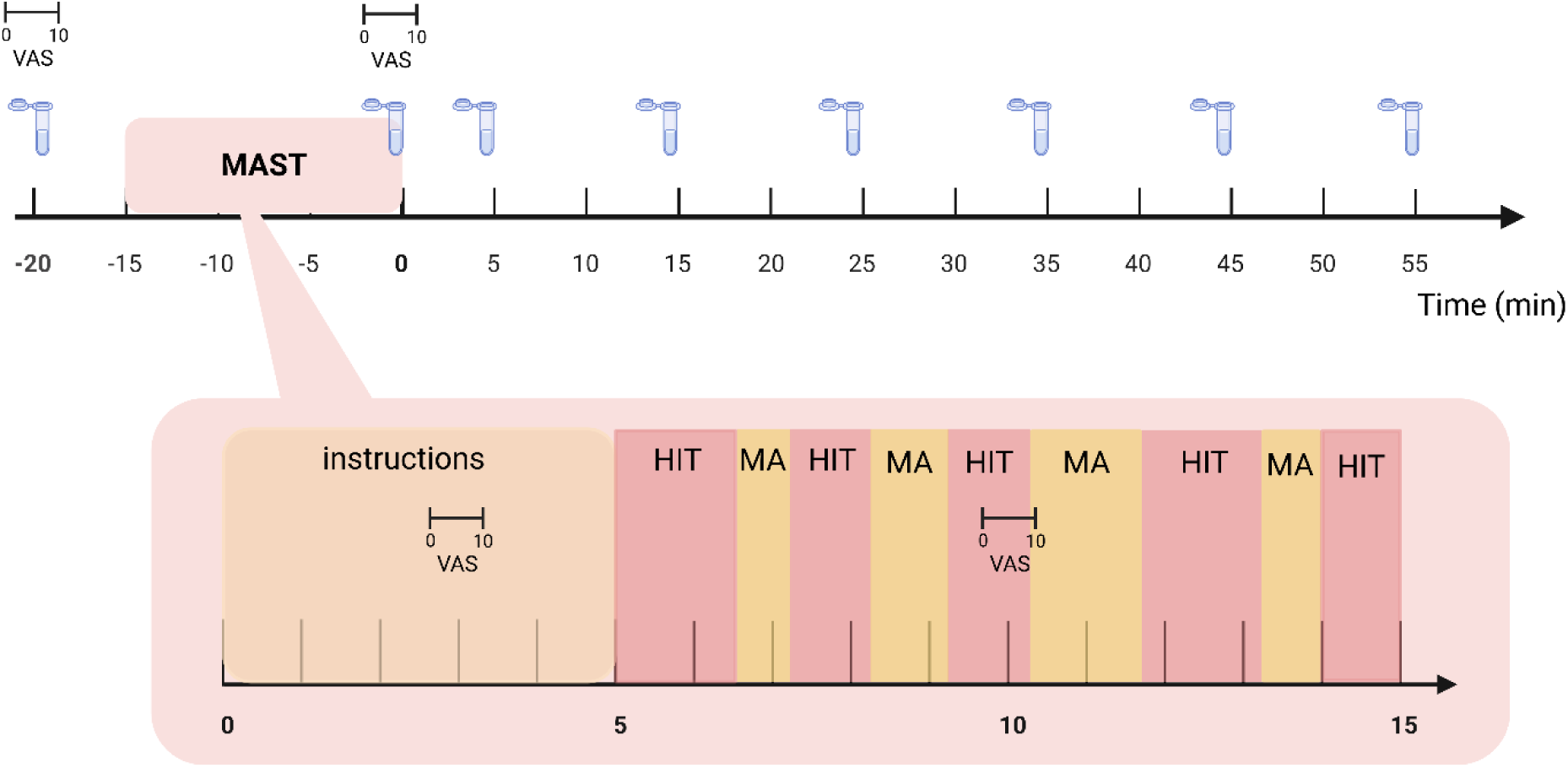
Design of the Maastricht Acute Stress Test (MAST). HIT= hand immersion trial, MA= mental arithmetic task, VAS=visual analogue scale.

##### 2.5.1.7 Fatigue and fatigability lab measures

To measure increases in and recovery from cognitive fatigue following a cognitive challenge, participants will perform a modified version of the Paced Auditory Serial Attention Task (PASAT), a task requiring working-memory retrieval, shown to significantly increase cognitive fatigue in ME/CFS patients (17). Participants will listen to a pre-recorded sequence of numbers between one and nine, and are instructed to press a button every time two consecutive numbers add up to ten. Each number will be presented for 1500 ms and separated by 500 ms. Additionally, participants have to pay attention to a screen displaying rapidly (every 500 ms) and randomly changing numbers, increasing the attentional demands of the auditory task. The task will be presented three times for three minutes with 30-s breaks in between (17). State fatigue is measured with a Dutch translation of the Fatigue and Energy Scale (FES), a validated scale capturing the post-exertional aggravation of physical and mental fatigue in response to activities in the lab or in real life (42). Participants have to fill in the FES prior to the task, at both 30-s breaks, immediately after the task, and 1 h, 4 h and 24 h after finishing the task to monitor their fatigue.

To measure physical fatigability, a 25-min cycling task will be performed on an arm-leg cycling ergometer (AirBike inSPORTline Pro, SEVEN SPORT s.r.o., Prague, Czech Republic) similar to the procedure described in Keech et al. (2015) (42). This test is shown to induce significant physical fatigue in ME/CFS patients. The cycling rate will be individually calibrated by monitoring the heart rate by a chest band (Polar T34, Polar Electro, Kempele, Finland). Initially, participants will be asked to increase their pedaling rate until they reach 70% of their age-predicted maximal heart rate (=208 − (0.7 ∗ age)). For the remainder of the test, the pedaling rate will be regularly adjusted to maintain this heart rate within a 3 beats per minute error margin. Before, immediately after, and 1 h, 4h, and 24 h after the task, participants will score perceived physical fatigue with the FES (42).

Fatigability in both tasks will be quantified as the increase in fatigue from baseline to after the task, while recovery failure will be investigated by comparing ratings before and immediately after vs 1, 4 and 24h after task completion.

##### 2.5.1.8 Ecological measurements

Via the experience sampling method (ESM), we will be able to measure dynamic experiences in the daily life of the patients (108). During seven consecutive days, patients will be requested to fill in a brief, momentary survey ten times per day (68,110). Requests will be sent through the m-Path smartphone application, a GDPR-compliant platform. Self-reported data about levels of stress and nervosity (9, 15, 153), levels of mental and physical fatigue (42), levels of arousal and valence (8, 39, 48), levels of physical and mental activity and behavioral responses to it (11, 19, 77), rest, somatic symptoms (107, 149), and use of substances (20) will be collected in the person’s natural environment (see **Supplementary Table 2** for details on the questions and answer options). The requests will be sent in a semi-randomized way, with 8 one-hour blocks evenly distributed across the participant’s entire daily wakefulness period. Within the 1 hour blocks, the surveys will be sent at a random moment (68) (**Supplementary Figure 1**). In addition, respondents will also fill out a daily morning questionnaire (10, 14, 150), assessing sleep duration and perceived sleep quality, and an evening questionnaire, evaluating general feelings of fatigue and mood throughout the day (19, 22, 42). At the end of the week, a debriefing survey will be sent to get some insights in how the study went from the participant’s point of view (e.g. asking questions such as ‘Did the ESM period influence your mood?’) (68).

Additionally, during the same seven days, participants will wear a device (Chill+, Imec, Leuven, Belgium) day and night except during strenuous physical activities or water-related tasks. After implementing a signal quality indicator and algorithms for physiological feature selection, heart rate and pulse, electrodermal activity (EDA), skin temperature, and motion/activity levels (via 3D accelerometry) can be quantified.

After synchronization, the data from the wearable and the responses to the daily life questionnaires can together provide an overview of the functionality of the SRS in a non-laboratory setting. Additionally, physical fatigability in daily life can be assessed by modeling changes in momentary fatigue levels, measured with ESM, following challenging situations identified through ambulatory physiological data, actigraphy, and ESM responses.

##### 2.5.1.9 Neuroimaging

###### 2.5.1.9.1 MRI scan protocol

MRI scanning will be carried out on a 3T Philips Achieva DStream scanner with a 32-channel head coil. MRI acquisition will start with a T1-weighted 3D turbo-field-echo sequence anatomical scan. Next, brain metabolites will be examined with 1H-MRS using a Point Resolved Spectroscopy (PRESS) sequence. The sequence will allow us to non-invasively investigate levels of myo-inositol and choline. Based on significant case-control findings from previous MRS studies in ME/CFS (33, 67), single-voxel MRS will be performed in the pregenual anterior cingulate cortex (pACC). Details on positioning of the voxel are in Supplement. Afterwards, neural responses to psychosocial stress will be quantified using the Montreal Imaging Stress Test (MIST; (24)) during a functional MRI scan (blood oxygen level dependent (BOLD)) conducted with a Gradient Echo (EPI - Echo Planar Imaging) acquisition sequence. The MIST consists of four runs in which three conditions [rest, control, and experimental (i.e. stress)] are alternated in a block design (**Figure 4**). Next, a resting-state fMRI scan with a Gradient Echo (EPI) acquisition sequence will be acquired in order to assess functional connectivity. Finally, cerebral blood perfusion will be quantified with a Pseudo-Continuous Arterial Spin Labeling (pCASL) sequence. The MRI procedure will last approximately 70 minutes. Details on the scanning protocol are in Supplement.

**Figure 4.**
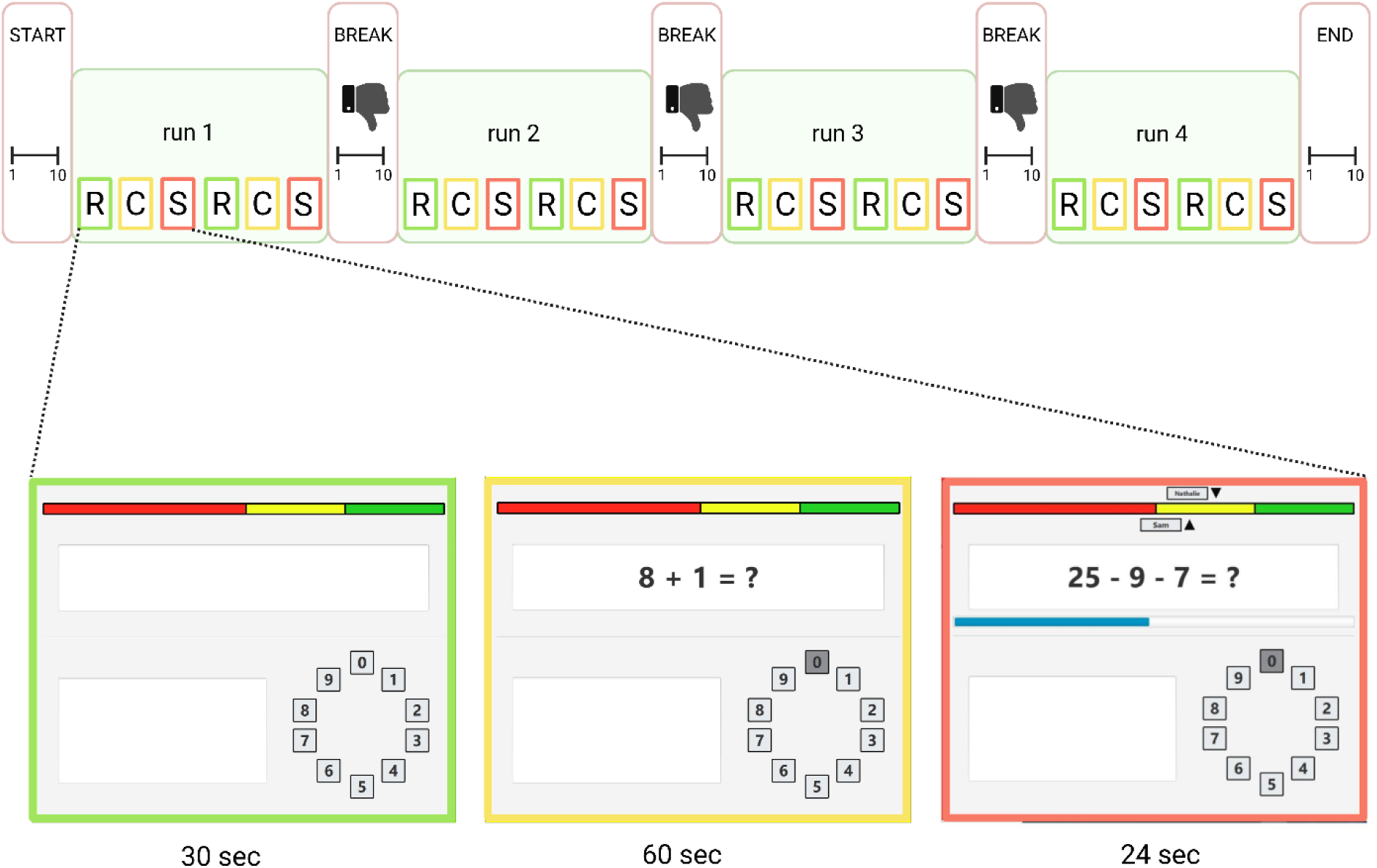
Montreal Imaging Stress Task (MIST) protocol. R=rest condition, C=control condition, S=stress (experimental) condition.

###### 2.5.1.9.2 PET/MR scan protocol

A subset of the sample (ME/CFS: N = 30, HC: N = 20) will undergo simultaneous MRI and PET scanning with a General Electric (GE, Milwaukee, WI, USA) Signa PET/MR equipped with time-of-flight (TOF) PET and a 3-Tesla MRI system with an 8-channel head coil. MRI sequences include (1) 3D volumetric T1-weighted BRAVO scan, (2) MRS scan with Probe-P (JPRESS) sequence, (3) Zero Echo Time (ZTE) scan for PET attenuation correction, (4) four fMRI MIST runs with gradient echo pulse sequence, (5) a resting-state fMRI scan with gradient echo pulse sequence, (6) ASL scan with 3D pCASL sequence. Simultaneously, dynamic 3D PET scans will be acquired for 60 min after a single intravenous injection of 120-150 MBq ^18^F-DPA714. Twenty 2 mL and five 5 mL arterial blood samples will be collected during the scan to allow for full kinetic modeling. Radioactivity in the 20 blood samples will be determined with a gamma counter that is cross-calibrated with the PET-MR scanner. The full scanning protocol at the PET/MR lasts approximately 75 min. Details are in Supplement.

#### 2.5.2 Participant characterization

##### 2.5.2.1 TSPO binding affinity

To enable reliable quantification of tracer binding, 10 mL of blood will be collected from the antecubital vein for genotyping in PET-eligible participants. A polymorphism in the human 18kDa TSPO, rs6971, gives rise to three distinct binding affinity profiles: high-, medium-, and low-affinity binders. Only individuals identified as high- or medium-affinity binders will be eligible for inclusion in the PET procedure, and binding affinity will be controlled for in all PET analysis. Analysis comprises DNA extraction and genetic sequencing carried out by the KU Leuven Center for Human Genetics and Genomics Core as described by Schroyen et al. (2021) (112).

##### 2.5.2.2 Psychiatric comorbidity

Psychiatric comorbidity will be evaluated using the Mini International Neuropsychiatric Interview Simplified (MINI-S) for the Diagnostic and Statistical Manual of Mental Disorders 5 (DSM-5) (78), a semi-structured interview evaluating the DSM-V criteria for the seventeen most common axis-I disorders.

##### 2.5.2.3 Self-report questionnaires

Participants completed questionnaires screening for the 1994 CDC criteria (32) and 2015 IOM criteria for ME/CFS (16), the 2010 American College of Rheumatology criteria for fibromyalgia (105), and the Rome IV diagnostic criteria for irritable bowel syndrome (IBS) (26). Furthermore, they completed the Generalized Anxiety Disorder-7 (GAD-7) (89), Patient Health Questionnaire-9 (PHQ-9) (45), Patient Health Questionnaire-15 (PHQ-15) (46), Childhood Trauma Questionnaire (CTQ) (5), Positive and Negative Affect Scale (PANAS) Trait version (25), Traumatic Experiences Checklist (TEC) (75), Checklist individual strength (CIS-20) (100), Frost Multidimensional Perfectionism Scale (FMPS) (31), Interoceptive Sensitivity and Attention Questionnaire (ISAQ) (7), The Medical Outcome Study Short Form – 36 items (SF-36) (27,28), Need for Controllability and Predictability questionnaire (NCPQ) (83), COVID-related questions. Details on the questionnaires and scoring are in Supplement.

##### 2.5.2.4 Demographics

We will record sex, age, socioeconomic information, lifestyle factors (e.g., smoking, substance use, fitness, dietary history), medication use, education level, diet, clinical history (e.g., ME/CFS-related: duration of symptoms, time since diagnosis), general personal and family medical history, and treatment history via a standardized self-created survey. Anthropometric measures such as height and weight will be measured in the lab during the study visit.

##### 2.5.2.5 Dietary intake

Participants will monitor their daily dietary intake for three days using the [MyFitnessPal](https://www.myfitnesspal.com/) app. They will enter each food item consumed, along with the quantity of each item.

### 2.6 Neuroimaging data

#### 2.6.1 Preprocessing

##### 2.6.1.1 Task-based fMRI (Montreal Imaging Stress Test)

Source data from the scanner will be converted from DICOM format into [The Brain Imaging Data Structure (BIDS)](https://bids.neuroimaging.io/) specification prior to preprocessing. [Mriqc version 0.16.0](https://mriqc.readthedocs.io/) will be used for quality control purposes. (f)MRI Preprocessing will be performed using fMRIPrep 20.2.6 (RRID:SCR_016216) (111, 143). Quality control and preprocessing details can be found in Supplement.

##### 2.6.1.2 Functional connectivity

After preprocessing using fMRIPrep 20.2.6 (RRID:SCR_016216) (111, 143), analyses of resting-state fMRI data will be performed using CONN (57) (RRID:SCR_009550) release 22.v2407 (58) and SPM (59) (RRID:SCR_007037) release 12.7771. Details on data preprocessing and denoising can be found in Supplement.

##### 2.6.1.3 PET

Preprocessing will be done with in-house scripts using MATLAB (R2021a, Natick, MA: The MathWorks Inc.) based on SPM12 (v7771, Wellcome Centre for Human Neuroimaging, University College London). The volume of distribution (V_T_) of [^18^F]-DPA714 will be quantified using a plasma-input based 2 tissue 4K model with an additional component describing irreversible endothelial binding and taking into account a fraction of whole blood in predefined ROIs and in parcels of the Canlab 2024 combined atlas (**Supplementary Table 3**). V_T_ is the primary outcome measure. For the ROI and parcel based analysis we will also estimate K_1_ and use this as a proxy for blood-brain barrier integrity (90).

##### 2.6.1.4 MRS

Data will be processed in the Osprey Toolbox v.2.9.0 (53) in MATLAB (R2021a, Natick, MA: The MathWorks Inc.) following standardized procedures for preprocessing and linear-combination modeling. Choline (CHO) and myo-inositol (MI) quantification will be performed as markers of neuroinflammation. Total N-Acetylaspartate (tNAA), a proxy marker of neuronal health (106), and glutamate/glutamine (Glx), amino acids important in neuronal regulation (152), concentrations will be assessed in an explorative way. Metabolite concentrations (institutional units (i.u.)) will be water-scaled using the unsuppressed water as a reference signal. Details on preprocessing and quality parameters can be found in Supplement. **Figure 5** shows an exemplary voxel position and spectra after model fit.

**Figure 5.**
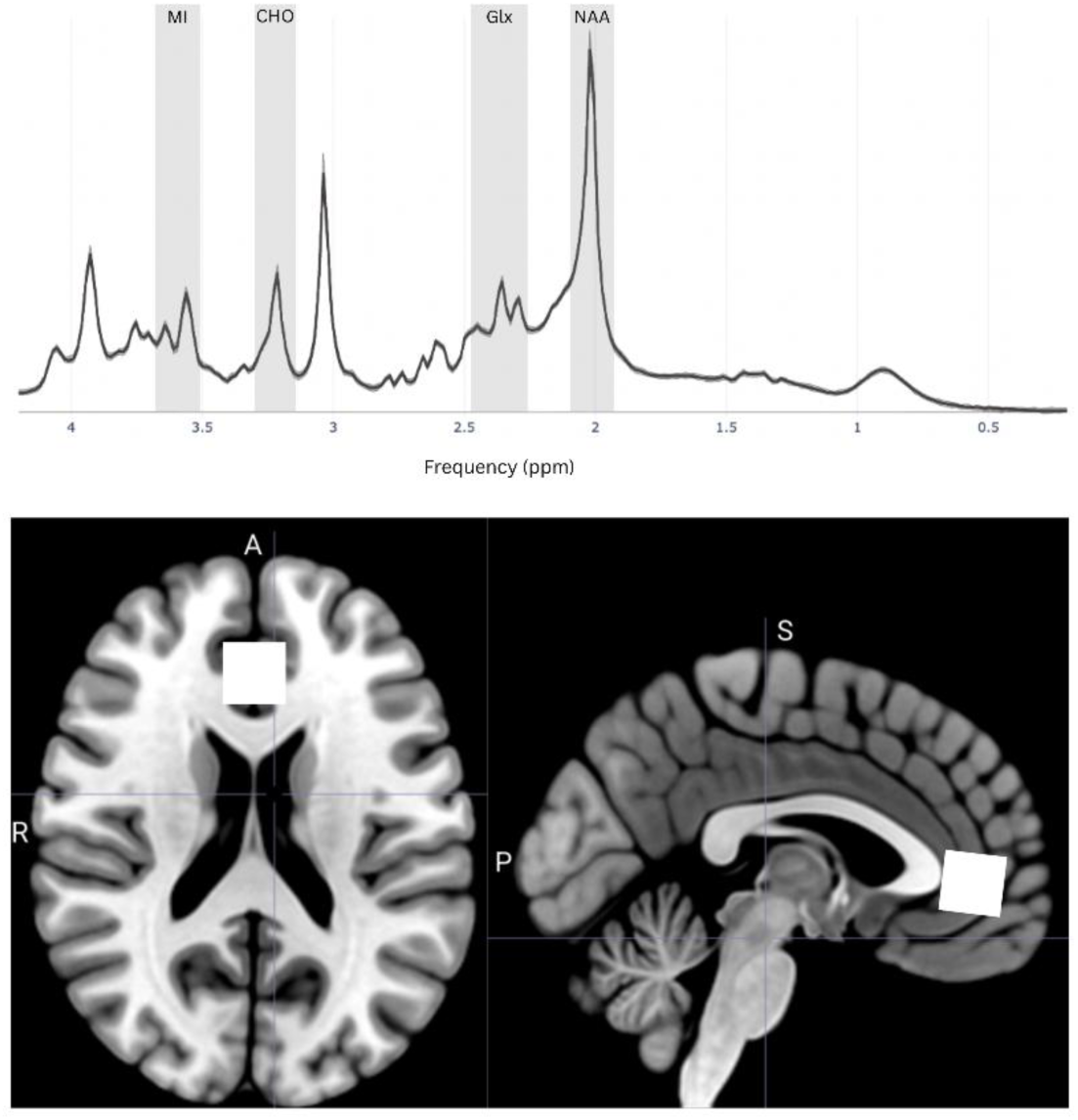
Representation of exemplary magnetic resonance spectroscopy (MRS) raw spectra and voxel positions. The myo-inositol (MI), choline (CHO), glutamate/glutamine (Glx), N-acetyl aspartate (NAA) peaks are highlighted in gray. Exemplary voxel position for the pregenual anterior cingulate cortex (pACC) is highlighted in white. Spectra adapted from Clarke et al. (2025) (29, 54). Brain adapted from MRIcroGL (v1.2.20220720) (55).

#### 2.6.2 ROI definition

##### 2.6.2.1 Task-based fMRI (Montreal Imaging Stress Test)

ROIs for task-based analyses (MIST) were selected based on their consistent association with stress across prior meta-analyses (116, 125, 130, 132) (details in Supplement). An overview of the included regions is illustrated in **Figure 6**.

**Figure 6.**
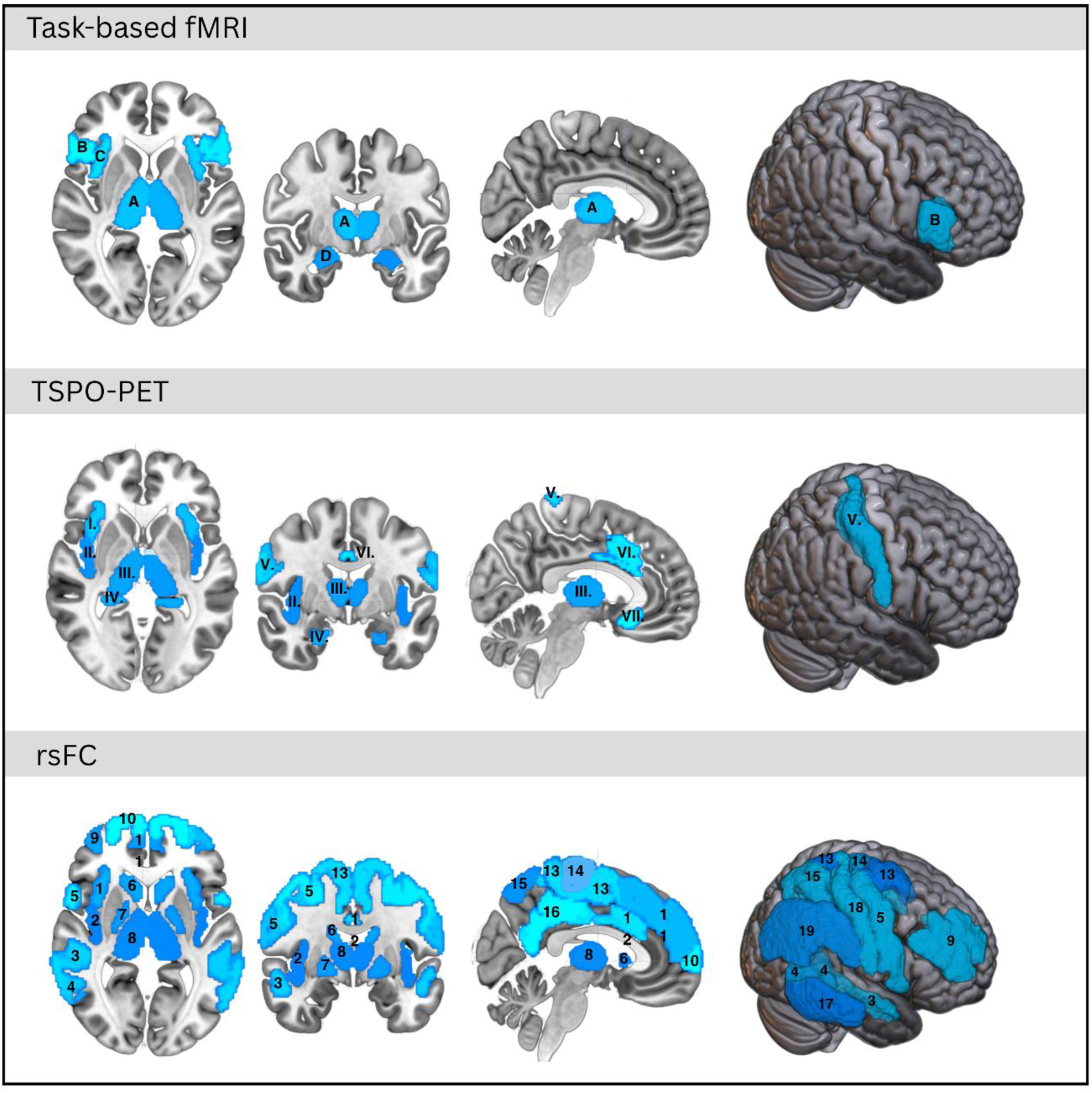
Overview of the regions of interest (ROIs) included in the neuroimaging analyses. ROIs for fMRI during the Montreal Imaging Stress Task (MIST): A= thalamus, B= inferior frontal gyrus, C= anterior insula, D= amygdala. ROIs for PET : I.= anterior insula, II.= posterior insula, III.= thalamus, IV.= hippocampus, V.= primary somatosensory cortex (S1), VI.= anterior midcingulate cortex (aMCC), VII.= subgenual cingulate cortex (sgACC). ROIs for resting-state fMRI (rsfMRI): 1= anterior insula, 2= posterior insula, 3= auditory association cortex, 4= temporal parietal occipital junction (TPOJ), 5= premotor cortex, 6= caudate, 7= globus pallidus, 8= thalamus, 9= dorsal lateral prefrontal cortex (dlPFC), 10= frontal pole, 11= dorsal medial prefrontal cortex (dmPFC), 12= anterior mid-cingulate cortex (aMCC), 13= somatomotor paracentral cortex, 14= primary motor cortex (M1), 15= superior parietal lobe (SPL), 16= precuneus, 17= middle temporal gyrus (MTG), 18= primary somatosensory cortex (S1), 19= inferior parietal lobe (IPL). ROI for the Magnetic Resonance Spectroscopy (MRS): pregenual anterior cingulate cortex (pACC). ROIs are always bilateral.

##### 2.6.2.2 Functional connectivity

ROIs for functional connectivity analysis were defined based on previous case-control and intervention studies in ME/CFS populations (69, 117, 118, 124, 127, 137, 139, 140) (details in Supplement). Included ROIs are presented in **Figure 6**.

##### 2.6.2.3 PET

ROIs were selected based on prior TSPO PET studies in affective and functional somatic disorders (148), including major depressive disorder (119–123, 126, 133, 135, 136), fibromyalgia (113, 131), chronic pain (4, 114, 115, 120, 128, 138), ME/CFS (72) and functional somatic syndromes (129) (See **Figure 6**) Details are explained in Supplement.

### 2.7 Statistical analysis plan

Except stated otherwise, statistics will be performed using SAS Analytics Software 9.4 (SAS Institute, Cary, NC, USA), JMP® Pro 18 (JMP Statistical Discovery LLC, Cary, NC, USA), JASP (Version 0.95.4) and MATLAB R2023a (Mathworks, Natick, MA, USA).

#### 2.7.1 Objective 1: comparison of neuropsychophysiological measures between ME/CFS patients and healthy participants

##### 2.7.1.1 SRS, SCFAs, fatigue/fatigability, immune markers

For objective 1 (see **Table 1**), we will compare SRS, SCFAs, immune and fatigability outcomes between patients with ME/CFS and healthy participants using (multivariate) one-way analysis of variance [(M)ANOVA] for the different (groups of) outcomes. For outcomes measured repeatedly during the test session (i.e. ratings and physiological measures in response to tasks), linear mixed models will be used, with time and group (ME/CFS patients versus healthy participants) as the fixed factors. When appropriate, multiple testing corrections will be applied using positive false discovery rates (FDR; q_FDR_) (37, 38) (i.e. when the estimated number of true significances by the bootstrap and/or spline procedure > 0), otherwise standard Benjamini-Hochberg FDR (p_FDR_) will be used (49) and covariates will be controlled (ANCOVA) when appropriate.

##### 2.7.1.2 Gut microbiome

Analysis for the gut microbiome data will happen in R primarily through the packages *vegan*, *phyloseq*, and *coda.base*, which offer key functionalities for gut microbiome data analysis. Statistical analyses will include computing and comparing diversity metrics, evaluating overall microbial composition via ordination methods, and identifying group differences and associations with host metadata. Generalized linear models will be used to account for confounding factors, including metabolic health. More details are in Supplement.

##### 2.7.1.3 Brain task-based fMRI

Second-level fixed effects analysis, comparing patients and healthy participants, will be performed on first-level stress minus control contrasts using GLMs implemented in MATLAB scripts calling CANlab and SPM12 functions.

Whole-brain voxel-wise GLMs using a gray matter mask will be performed, using a voxel-wise threshold of q_FDR_ < 0.05. The whole-brain analyses will be complemented by ROI-based (M)ANOVAs with positive FDR correction (q_FDR_ < 0.05) (see “2.6.2 ROI definition” for ROI selection and justification). GLM analyses will be corrected for scanner type (MR vs. PET/MR) (fixed effect). Whole brain multivariate mediation analysis will be performed using the “principal directions of mediation” (PDM) method (144,145) in order to study brain patterns mediating case-control differences in self-reported NA during the MIST. Analyses will be done using the CANlab mediation toolbox and will be thresholded voxel-wise at q_FDR_ < 0.05. Finally, multi-voxel pattern analysis (MVPA) will be employed with The Decoding Toolbox (35) to train and cross-validate (k-fold) a Support Vector Machines classifier to distinguish ME/CFS patients from healthy controls based on their brain response to stress. Statistical significance (voxel-wise p-values) will be estimated using 5,000 permutation tests, preserving group distribution in each fold during the label shuffling. Details are in Supplement.

##### 2.7.1.4 Brain resting-state fMRI

Resting-state functional connectivity will be determined via ROI-to-ROI connectivity among 38 ROIs (see “2.6.2 ROI definition” for ROI selection and justification) analysis using multivariate GLM (with CONN (57) (RRID:SCR_009550) release 22.v2407 (58) and SPM (59) (RRID:SCR_007037) release 12.7771). Results will be thresholded using a combination of uncorrected p < 0.01 at connection-level threshold and familywise error corrected (p_FWE_ < 0.05) at ROI-level (ROI mass/intensity). Graph Theoretical Analysis will be performed with the GraphVar Toolbox (146, 147) version 2.03a. Group comparisons will be performed with one-way ANOVAs and MANOVAs. Positive FDR correction (q_FDR_ < 0.05) will be applied when appropriate, otherwise standard Benjamini-Hochberg FDR will be used (49). Furthermore, Partial Least Squares (PLS) - Discriminant Analysis (DA) using the nonlinear iterative partial least squares (NIPALS) method implemented in JMP® Pro 18 software will be implemented to test how well the graph measures are able to distinguish ME/CFS patients from healthy participants. Furthermore, Functional Connectivity Multivariate Pattern Analysis (fc-MVPA) will be performed (104). Voxel-level results will be determined using multivariate parametric statistics with random-effects across subjects and sample covariance estimation across measurements. Cluster-level inferences (groups of adjacent voxels) will be derived from nonparametric statistics using Threshold Free Cluster Enhancement (TFCE), with 5000 residual-randomization iterations. Results will be familywise error corrected (p_FWE_ < 0.05). More details in Supplement.

##### 2.7.1.5 Brain PET

Case-control comparisons for ROI data will be performed using MANOVA with (positive) FDR correction (<0.05). In addition, PLS-DA will be used to test how well the ME/CFS patients can be distinguished from healthy participants based on PET outcomes (V_T_ and K_1_). To account for differences in genotypes in the PET analyses, we will use the residuals, after regressing the dependent variables on binding-affinity (high or medium affinity binding), in the analyses.

##### 2.7.1.6 Brain MRS

Group differences for MI and CHO concentrations in the ACC will be analyzed using a one-way MANOVA with q_FDR_ correction (<0.05). Other metabolites (tNAA, Glx) will be evaluated exploratorily.

#### 2.7.2 Objective 2: Test the interactions between the neuropsychophysiological mechanisms in ME/CFS

For objective 2, structural equation modeling (SEM) will be used to investigate the hypothesized relationships among the SRS, immune functioning, gut-microbiome, SCFAs, and brain-related outcomes in the patient sample. A path diagram illustrating the hypothesized model is shown in **Figure 7**. Model identification and fit will be checked to ensure that all parameters can be uniquely estimated from the available data. To assess the indirect effects, 5000 bootstrap samples will be used, providing robust estimates of the confidence intervals for the indirect effects.

**Figure 7.**
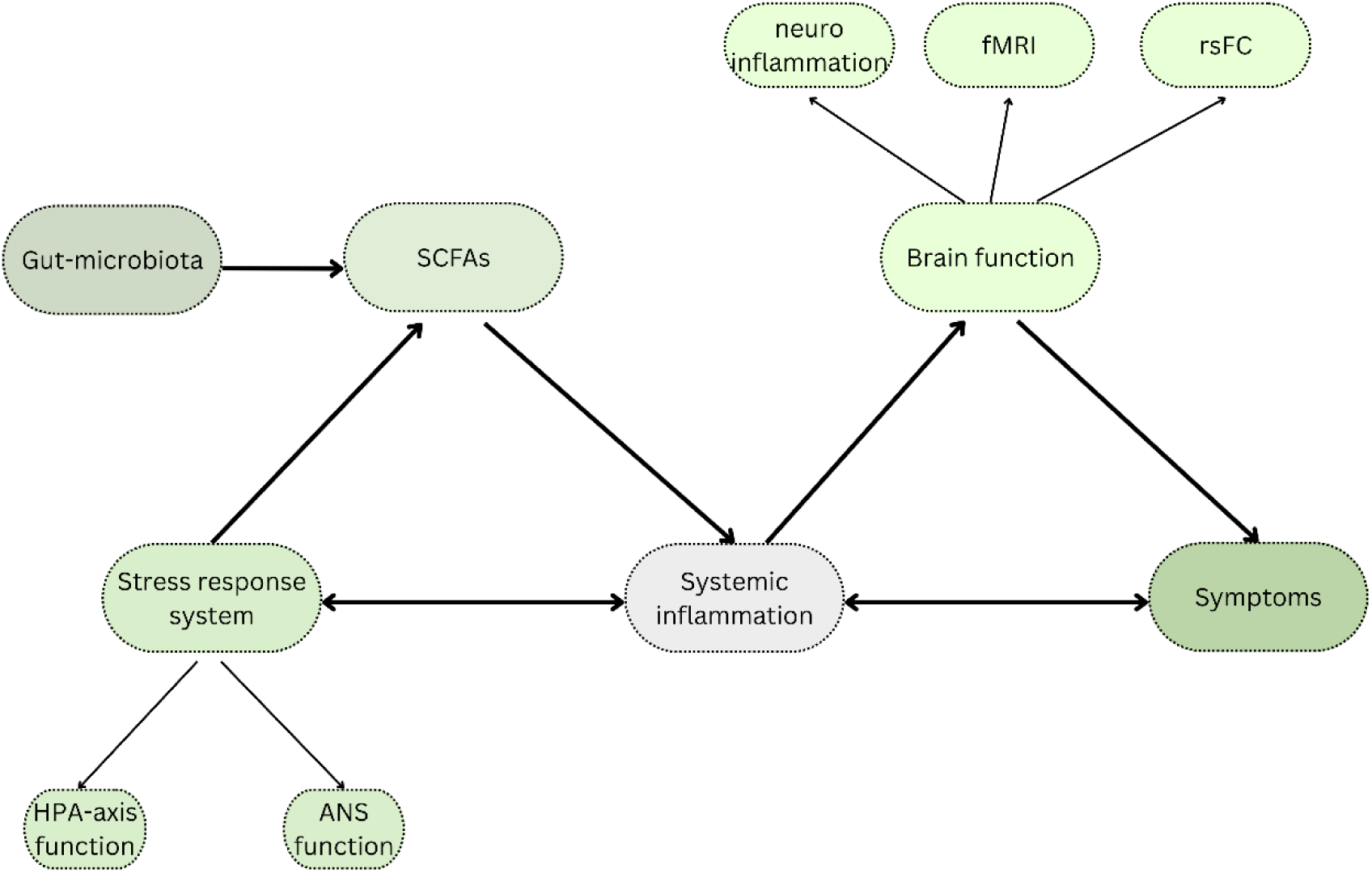
Path model visualizing the hypothesized neuropsychophysiological relationships in the ME/CFS patient sample. SCFAs= short-chain fatty acids, HPA-axis= hypothalamic-pituitary-adrenal axis, ANS= autonomic nervous system, fMRI= functional magnetic resonance imaging, rsFC=resting-state functional connectivity.

For research question 2 (see **Table 1**), each set of input variables will be examined for associations with fatigue and fatigability outcomes using multiple regressions. Voxel-wise neural and neurochemical measures will additionally be assessed using least absolute shrinkage and selection operator principal component regression (LASSO-PCR). Because we expect substantial intercorrelations among the input variables, we will also use partial least squares (PLS) regression to assess how combinations of these measures collectively relate to fatigue and fatigability.

Furthermore, we will test whether brain functioning mediates the associations between the input variables and fatigue/fatigability using multivariate mediation analyses using the PDM method.

#### 2.7.3 Objective 3: To identify subgroups of ME/CFS patients based on inter-individual differences

We will test the hypothesis that data-driven multimodal CFS subtypes can be identified based on a combination of mechanisms. Unsupervised clustering algorithms, like Gaussian mixture models, and co-clustering methods (56, 151) will be applied on data from all neuropsychophysiological levels.

#### 2.7.4 Objective 4: Use these subgroups as potential predictors of longitudinal effects throughout treatment

For objective 4, linear mixed models will be used to analyze the evolution of outcome variables such as symptom severity, fatigability, quality of life and activity levels (self-reported, lab-based, and ESM-based) in order to examine treatment response. In addition, latent class growth analysis (LCGA) will be used to assess individual differences in treatment responses by identifying subgroups based on treatment response trajectories. LCGA clusters participants based on both the baseline level of functioning (intercept) and its trajectory over time (slope, linear and higher-order depending on model fit). Fit statistics, such as the Bayesian Information Criterion (BIC) and entropy, will be applied to decide the number of subgroups resulting from the LCGA. Given our sample size, we will not choose a LCGA solution in which one of the clusters contains less than 10% of the sample. Additionally, by adding subgroup membership as categorical predictors to the linear mixed models and LCGAs we will test whether the subgroups defined in objective 3 are predictors of treatment trajectories. Further, multiple regressions, LASSO-PCR, and PLS will be used in a similar way as objective 2.2 to determine mechanisms underlying treatment response.

## 3. Ethics Approval

We will conduct the trial in compliance with the principles of the Declaration of Helsinki (version, 2013), and in accordance with all applicable regulatory requirements. This protocol and related documents were reviewed and approved by the Ethics Committee Research UZ/KU Leuven (Herestraat 49, 3000 Leuven), ref. S66452 in September 2022. Signed informed consent form (ICF) for all participants will be applied prior to their enrollment and participation in the study. Privacy rights of human subjects have been acknowledged.

## CRediT authorship contribution statement

**Ynse Dooms**: Writing – original draft, Visualization, Methodology, Investigation, Formal analysis, Data curation, Project administration. **Lixin Qiu**: Writing – original draft, Visualization, Methodology, Formal analysis, Data curation. **Iris Coppieters**: Methodology, Investigation, Writing – review and editing. **Elfi Vergaelen**: Conceptualization, Resources, Writing – review and editing. **Stephan Claes**: Conceptualization, Recourses, Writing – review and editing. **Patrick Dupont**: Formal analysis, Writing – review and editing. **Melina Hehl**: Formal analysis, Writing – review and editing. **Koen Cuypers**: Formal analysis, Writing – review and editing. **Harald Engle**r: Formal analysis, Writing – review and editing. **Kirsten Dombrowski**: Formal analysis. **Kristin Verbeke**: Formal analysis, Writing – review and editing. **Omer Van den Bergh**: Conceptualization, Writing – review and editing. **Jeroen Raes**: Formal analysis, Writing – review and editing. **Lukas Van Oudenhove**: Conceptualization, Methodology, Formal analysis, Data curation, Resources, Funding Acquisition, Writing – review and editing. **Maaike Van Den Houte**: Conceptualization, Methodology, Investigation, Formal analysis, Data curation, Resources, Supervision, Writing – review and editing. **Katleen Bogaerts**: Conceptualization, Methodology, Resources, Funding Acquisition, Writing – review and editing.

## Data availability

The code supporting this work is available in open Github repositories. The in-house scripts used for the PET preprocessing and VT and K1 calculations can be downloaded from the public repository https://github.com/labgas/LaBGAScore/tree/main/pet/scripts

The custom scripts to calculate first-level contrasts during the MIST can be downloaded from https://github.com/labgas/LaBGAScore/tree/main/firstlevel Scripts on the second-level fixed effects analysis can be found at https://github.com/labgas/CANlab_help_examples

Mediation analyses will be conducted with the use of https://github.com/canlab/MediationToolbox The ethics protocol and datasets that will be generated and/or analysed during the current study will be available from the corresponding author upon reasonable request.

## Acknowledgments

We would like to thank Greet Vandermeulen, Sophie Wieczorek, Marta Walentynowicz and Master students Julie Stegen, Silvana Sokolji, Ditte Coopmans, Emily Van der Schueren, Bente Ponsaerts, Karim Tahri, and Caressa Haerts.

## Author Contributions

LVO, KB and MVDH were involved in the project’s funding acquisition and supervision. LVO, MVDH, KB, LQ, EV, SC, PD, MH, KC, KV, OVDB, JR, and YD were all involved in the conceptualisation, methodology and/or data analysis plan of the project. YD, MVDH, and IC collected the data of the study project. YD and LQ wrote the original draft of this manuscript. All authors revised the manuscript and agreed to be accountable for all aspects of the work.

