## Supplementary files for "Multimodal approach to identify neuropsychophysiological subgroups in myalgic encephalomyelitis/chronic fatigue syndrome and their relevance for rehabilitation: protocol for a mechanistic cross-sectional and longitudinal study"

### Supplementary Text

#### 2. Methods

**2.2 Standardized rehabilitation program**

According to the NICE guidelines (106) and previous evidence (105), CBT is recommended as a supportive therapy to alleviate symptoms, coping with the agony of having a chronic disorder, and ameliorate functioning. It has shown to be moderately effective in alleviating symptoms and enhancing wellbeing, albeit not in all patients (105).

**2.3 Participants**

Exclusion criteria for all participants are (1) pregnancy or lactation, (2) use of medications affecting stress physiology, e.g. corticoids, beta blockers, opiates, other anti-inflammatory treatment, and use of more than one antidepressant type or more than two antidepressants at low doses, assessed on a case-by-case basis, (3) alcohol or drug dependency, (4) consumption of antibiotics within three months prior to baseline, (5) presence of a severe medical condition (e.g. malignancy, severe heart disease, kidney disease, neurological deficit) and/or psychiatric disorders (except secondary depression, social and generalised anxiety disorder, panic disorder, agoraphobia, post-traumatic stress disorder, acute stress disorder and obsessive compulsive disorder for patients), (6) being younger than 18 and older than 66 at baseline visit, and (7) having the ability to understand and speak the Dutch language. Participants partaking in brain imaging must not (1) experience claustrophobia, (2) have back problems making it impossible to complete a 75-minute brain scan, and (3) have a condition or implant that interferes with magnetic resonance imaging (MRI) (e.g. cardiac pacemaker, neural stimulator, metal fragments in the body). Participants undergoing the PET/MR scan, in addition to the previous criteria, cannot (1) be exposed to more than 1 mSv of ionizing radiation in the past twelve months, (2) be allergic to the radioactive contrast agent, and (3) be low affinity binders of the TSPO radiotracer - participants will be screened for the genetic TSPO polymorphism rs6971 as recommended (6). All participants will need to submit written informed consent to participation before data collection.

**2.4 Sample size**

The aimed sample size yields 80% power to detect group differences with small effect sizes ($f<0.25$) for up to 7 variables and medium effect sizes ($f=0.31$) for up to 175 variables in one-way analyses of variance (ANOVAs) with Holm correction. Highly interrelated dependent variables (e.g. serum inflammatory markers) will be assessed with one-way multivariate analyses of variance (MANOVAs). The included sample size yields 80% power to detect small-to-medium effects ($f^2=0.13$) for 10 response variables and medium effects ($f^2=0.23$) for up to 50 variables by MANOVAs with Holm correction. Power for cluster analysis is dependent on cluster size and separation. A study with 116 adolescent ME/CFS patients investigating autonomic, immunological, neuroendocrine, cognitive and sensory processing functionalities was able to identify six clusters (108). Conservatively assuming no, small, or medium effect sizes for the individual variables (multivariate effect size $\Delta=4$), $n=115$ yields 80% power to detect up to 4 equally sized clusters (109). To maximize power, we will use dimensionality reduction (Principal Components Analysis (PCA), multidimensional scaling) and- powerful finite mixture models as recommended (109). Due to expected drop-out for the neuroimaging measures, e.g. by not fulfilling the extra inclusion criteria, we intend to include 105 ME/CFS patients and 50 healthy people for the

MR scans. This subsample is powered to detect medium effect sizes (f=0.27) by one-way ANOVAs with Holm correction for the 10 regions of interest (ROIs). Additionally to the expected drop-outs, participant inclusion for the PET measures will be affected by scanner availability and costs. We anticipate a PET subsample of 30 ME/CFS patients and 20 HC. Participants will be included on a first-come, first-served basis. 19 HC are already available, historical HC, who underwent an identical PET imaging protocol on the same General Electric Signa PET/MR scanner. Historical HC data will be derived from three studies: (I) Van Weehaeghe et al. (2020) amyotrophic lateral sclerosis study (N = 4, September 2017 - October 2018) (112), (II) 22q22 deletion syndrome study (N = 5, June 2017 - May 2018; unpublished), (III) Schroyen et al. (2022) early-stage breast cancer study (N = 10, May 2019 - August 2020) (111). This sample is powered to detect large effect sizes (f=0.49) by one-way ANOVAs with Holm correction for the 14 ROIs. In addition, this sample size is sufficient to determine whether patient clusters predict treatment trajectories with latent class growth analysis (LCGA) (109). Furthermore, treatment responses can be evaluated with medium effect sizes ($f^2$=0.18) for 10 key predictors by multiple linear regressions. The sample size is also sufficient for least absolute shrinkage selection operator principal component regression (LASSO-PCR) (28). All outcomes are obtained with sensitivity power calculations in GPower 3.1.9.7 (110)).

**2.5 Measures**

2.5.1 Primary outcome variables

2.5.1.1 Systemic inflammation

Serum levels of TNF-α, IL-1β, IL-4, IL-6, IL-8, IL-10 and IFN-γ will be quantified by multiplex electrochemiluminescence immunoassay using the V-PLEX Human Proinflammatory Panel 1 kit, and TGF-β will be measured using the U-PLEX Human TGF-β1 Assay. The sensitivity (LLOQ) of these assays are 0.69 pg/ml (TNF-α), 0.646 pg/ml (IL-1β), 0.218 pg/ml (IL-4), 0.633 pg/ml (IL-6), 0.591 pg/ml (IL-8), 0.298 pg/ml (IL-10), 1.76 pg/ml (IFN-γ), and 5.0 pg/mL (TGF-β). Intra-assay coefficients of variation (CVs) are typically less than 7% for V-plex and below 5.7% for U-plex kits. Inter-assay CVs are generally below 15% in V-plex and 16.8% in U-plex assays for low controls based on the manufacturer's specifications. All panels will be analyzed on the Meso QuickPlex SQ 120 platform (Meso Scale Diagnostics LLC, Maryland, USA). A standard curve will be established for each plate. A control will be prepared using only the diluent.

2.5.1.2 General health biomarkers

Markers include alanine aminotransferase (ALT), aspartate aminotransferase (AST), albumin, bilirubin, creatine kinase (CK), hemoglobin, cholesterol, gamma-glutamyl transferase (GGT), triglycerides, urea and uric acid analysed via colorimetry (COBAS 8000, Hitachi, Tokyo, Japan/Roche Diagnostics, IN, USA and XN-1000/XN-2000, Sysmex, Kobe, Hyogo, Japan), ferritin and vitamin B12 analysed via immunoassays (COBAS 8000, Hitachi, Tokyo, Japan/Roche Diagnostics, IN, USA), platelet characteristics like platelet count, mean platelet volume (MPV), platelet-large cell ratio (P-LCR), plateletcrit (PCT), platelet distribution width (PDW) measured with XN-1000 and XN-2000 (Sysmex, Kobe, Hyogo, Japan), hematocrit, mean corpuscular hemoglobin (concentration) (MCH(C)), mean corpuscular volume (MCV), macro and micro erythrocytes, red blood cell count, relative distribution width (RDW) quantified with electrical impedance (XN-1000 and XN-2000, Sysmex, Kobe, Hyogo, Japan), erythrocyte sedimentation rate (ESR) via aggregation capacity of the erythrocyte measured with capillary photometry (Test1 SDL, Alifax, Polverara, Italy), neutrophil granular intensity (NEUT-GI), neutrophil reactive intensity (NEUT-RI), white blood cell differential, and white blood cell count analysed with specific lysis, light scattering, and fluorescence (XN-1000 and XN-2000, Sysmex, Kobe, Hyogo, Japan), glycohemoglobin (HbA1c) measured with high-performance liquid chromatography (HLC-723G8, TOSOH, Tokyo, Japan), creatinin concentration quantified with the enzymatic coupled creatininase peroxidase method (COBAS 8000, Hitachi, Tokyo, Japan/Roche Diagnostics, IN, USA), and proteins after electrophoresis (albumin, alpha 1-globulin, alpha 2-globulin, beta-globulin, gamma-globulin) analysed with capillary zone electrophoresis (Capillarys 3 Octa, Sebia, Lisses, France).

2.5.1.5 Short chain fatty acids (SCFAs)

The lowest analyte concentrations measurable with a CV < 20% with this technique are 10 μM, 0.5 μM and 1 μM for acetate, butyrate and propionate respectively.

2.5.1.6 Salivary cortisol response to psychosocial stress

Samples will be analyzed using enzyme-linked immunosorbent assay (ELISA) analysis (Cortisol Saliva ELISA, RE52611, TECAN/IBL International, Hamburg, Germany). The mean intra-assay CV is typically 4.3% and the mean inter-assay CV is typically 13.2%. The LoQ is 0.138 nmol/l.

2.5.1.9 Neuroimaging

2.5.1.9.1 MRI scan protocol

MRI scanning will be carried out on a 3T Philips Achieva DStream scanner with a 32- channel head coil. MRI acquisition will start with a T1-weighted 3D turbo-field-echo sequence anatomical scan (182 sagittal slices; field of view (FOV): 256 (FH) x 242 (AP) x 182 (RL); matrix: 256x240x182; flip angle: 8°; Echo Time [TE]: 4.6 ms; Repetition Time [TR]: 9.7 ms; 1x1.01x1 $mm^3$ voxels). Next, brain metabolites will be examined with 1H-magnetic resonance spectroscopy (MRS) using PRESS sequence (TE: 35 ms; TR: 2 s; spectral bandwidth: 2kHz; 20x20x20 $mm^3$ voxel; Number of Averaged Spectra (NA): 128; automatic second-order pencil-beam (PB) shimming; MOIST water suppression (96)). The voxel of interest (VOI) will be positioned individually based on anatomical landmarks (in the sagittal plane, anterior to the anterior cingulate cortex, with its inferior border aligned along the anterior commissure–posterior commissure (AC–PC) line) identified on each participant's T1-weighted MRI scan and in accordance with the Canlab 2024 combined atlas (imported into CONN). Afterwards, neural responses to psychosocial stress will be acquired using the Montreal Imaging Stress Test (MIST; 107)) during a functional MRI scan (blood oxygen level dependent (BOLD)). 4 fMRI MIST runs will be conducted with a Gradient Echo (EPI - Echo Planar Imaging) acquisition sequence (60 transverse slices; FOV: 224 (RL, AP), 132 (FH); matrix: 112x109x60; flip angle: 90°; TE: 30 ms; TR: 1.8 s; 2x2x2 $mm^3$ voxels). The MIST consists of four runs in which three conditions (rest, control, and experimental) are alternated; the runs are presented in sequential blocks as a block paradigm (**Figure 4 in main text**). Each run consists of two rest (15 frames), two control (30 frames) and two experimental (60 frames) blocks, and lasts a total of seven minutes and thirteen seconds. In the rest condition, participants look at a static computer screen on which no task is shown. In the control condition, the computer screen displays a series of mathematical equations of varying difficulty, without a time-restraint or feedback. Participants submit their answer by selecting the correct answer using a response box. In the stress condition, the computer manipulates the difficulty and time limit of the tasks to be just beyond the individual's mental capacity. Manipulation of both the time limit and difficulty level results in better performance triggering harder-to-solve equations so that approximately 50% of responses are correct on average. Finally, to add a social stress component to the task, participants are told they will compete against another participant who they briefly meet before the scan, and whose performance they can follow during the task, and who always outperforms the participant. In reality, the opponent is a stooge and will not engage in the task itself. In between runs, participants will receive scripted negative verbal feedback and will rate, on a scale from 1 to 10, how much they feel judged, pressured, gloomy, insecure, anxious, guilty, relaxed, cheerful, and whether they feel they meet expectations. Scores will be summed up (reverse scored where necessary) to create a general 'negative affect' rating after each MIST run. After the 4 runs, a resting-state fMRI scan with a Gradient Echo (EPI) acquisition sequence (42 transverse slices; FOV: 240 (RL, AP), 146 (FH); matrix: 112x110x42; flip angle: 80°; TE: 33 ms; TR: 1000 ms; 2.15x2.14x3 $mm^3$ voxels) will be acquired in order to assess functional connectivity. Finally, cerebral blood perfusion will be quantified with a Pseudo-Continuous Arterial Spin Labeling (pCASL) sequence (20 transverse slices; FOV: 240 (RL, AP), 120 (FH); matrix: 64x60x20; 3.75x3.75x6 $mm^3$ voxels). pCASL demonstrated a higher signal-to-noise ratio (SNR) than pulsed ASL (PASL) and greater tagging efficiency compared to continuous ASL (CASL) (2). The MRI procedure will last approximately 70 minutes. Pencil-beam (PB) shimming will be applied to optimize magnetic field homogeneity.

2.5.1.9.2 PET/MR scan protocol

A subset of the sample (ME/CFS: N = 30, HC: N = 20) will undergo simultaneous MRI and PET scanning with a General Electric (GE, Milwaukee, WI, USA) Signa PET/MR equipped with time-of-flight (TOF) PET and a 3-Tesla MRI system with an 8-channel head coil. MRI sequences include (1) 3D volumetric T1-weighted BRAVO scan (TE: 3.4 ms, TR: 8.8 ms, TI: 450 ms, matrix: 256x256x170, flip angle: 12°; 1x1x1 $mm^3$ voxels), (2) MRS scan with Probe-P (JPRESS) sequence (TE: 35 ms; TR: 1.5 s; spectral bandwidth: 6.94kHz; 20x20x20 $mm^3$ voxel; Number of Averaged Spectra (NA): 128; automatic second-order shimming; CHESS water suppression (97)), (3) Zero Echo Time (ZTE) scan for PET attenuation correction (flip angle: 0.8°, matrix: 110x110x116, 2.4x2.4x2.4 $mm^3$ voxels, number of averages 4, pixel bandwidth: 976.562 Hz), (4) 4 fMRI MIST runs with gradient echo pulse sequence (42 oblique slices, TE: 30 ms, TR: 1.8 s, flip angle: 80°; matrix: 96x96x42; 2.3x2.3x3 $mm^3$ voxels), (5) a resting-state fMRI scan with gradient echo pulse sequence (42 axial slices, TE: 30 ms; TR: 1.212 s, flip angle: 70°, matrix: 64x64x42, 3.4x3.4x4 $mm^3$ voxels), (6) ASL scan a 3D PCASL (TE: 11.3, TR: 4.879s, matrix: 512x8x36, 0.5x30x4 $mm^3$ voxels). Simultaneously, dynamic 3D PET scans will be acquired for 60 min after a single intravenous injection of 120-150 MBq $^{18}$F-DPA714. Twenty 2 mL arterial blood samples will be collected during the scan (10s, 20s, 30s, 40s, 50s, 60s, 70s, 80s, 90s, 100s, 120s, 140s, 60s, 180s, 4min, 5min, 10min, 20min, 40min and 60min) after tracer injection to derive the arterial input curve (all samples) and parent free fraction (five additional samples at the final five timepoints assessed with 5mL arterial blood) to allow for full kinetic modeling. Radioactivity in the 20 blood samples will be determined (both in whole blood samples and plasma samples derived from the arterial blood samples) with a gamma counter that is cross-calibrated with the PET-MR scanner. The full scanning protocol at the PET/MR lasts approximately 75 min. Automatic shimming will be applied for the MRI parts.

2.5.2 Participant characterization

2.5.2.3 Self-report questionnaires

2.5.2.3.1 Functional syndrome checklist

Participants completed questionnaires screening for the 1994 CDC criteria (119) and 2015 IOM criteria for ME/CFS (120), the 2010 American College of Rheumatology criteria for fibromyalgia (121), and the Rome IV diagnostic criteria for irritable bowel syndrome (IBS) (122).

2.5.2.3.2 Generalized Anxiety Disorder-7 (GAD-7)

(Generalized) anxiety severity is measured with the GAD-7 (123). The GAD-7 consists of seven anxiety symptoms, each rated based on their frequency during the last two weeks, ranging from 0 'not at all' to 4 'almost every day'. The questionnaire has cut-off points for mild, moderate, and severe anxiety.

2.5.2.3.3 Patient Health Questionnaire-9 (PHQ-9)

The PHQ-9 is a depression assessment module that detects the presence and evaluates the severity of depressive symptoms. Respondents are asked to rate how often they experienced each of the nine depressive symptoms in the last two weeks on a 4-point Likert scale (0: not at all, 4: almost every day). The questionnaire has cut-off levels for mild, moderate, moderately severe, and severe depressive symptoms (124).

2.5.2.3.4 Patient Health Questionnaire-15 (PHQ-15)

The PHQ-15 evaluates the occurrence and severity of fifteen common somatic symptoms. Respondents are required to indicate on a 3-point Likert scale to what extent they are affected by the symptoms in the past four weeks (0: not affected at all, 2: affected a lot). Cut-off scores can be used to define mild, moderate, and severe somatic symptom severity (125) .

2.5.2.3.5 Childhood Trauma Questionnaire (CTQ)

The CTQ will be used to assess early adverse experiences. The self-report questionnaire consists of 25 items regarding childhood that are rated on a scale ranging from 0 'never true' to 5 'very often true'. In addition to a total trauma sum score, the CTQ can examine trauma across the subtypes physical abuse, emotional abuse, sexual abuse, physical neglect and emotional neglect (126).

2.5.2.3.6 Positive and Negative Affect Scale (PANAS) Trait version

The PANAS is a scale consisting of 20 items, measuring negative and positive emotions, that each need to be scored on a 5 point Likert scale estimating how often they are experienced in daily life (127).

2.5.2.3.7 Traumatic Experiences Checklist (TEC)

The TEC is used to identify potentially traumatic events in a person's life. Respondents need to indicate for 29 statements if they were exposed to a specific event / experience, to what extent the corresponding event impacted them (0: no impact, 5: very severe impact), and their age at the onset and end of the event. The TEC trauma presence score is calculated by summing the number of traumatic events a participant was exposed to. The TEC trauma impact score is calculated by summing the impact scores (128).

2.5.2.3.8 Checklist individual strength (CIS-20)

Self-reported feelings of fatigue and behavioral aspects related to these feelings are assessed via a list of 20 statements, applying to the last two weeks, each scored on a 7-point Likert scale ranging from 'yes, that's correct' to 'no, that's not correct'. Additionally to a total score, subscores will be calculated for subjective fatigue, concentration, motivation, and physical activity (129).

2.5.2.3.9 Frost Multidimensional Perfectionism Scale (FMPS)

The Frost Multidimensional Perfectionism Scale measures perfectionism. Respondents have to rate 35 items on a 5 point Likert scale going from 1: totally not true, to 5: totally true. Six different subscales can be distinguished (1) concern over mistakes, (2) personal standards, (3) parental criticism, (4) parental expectations, (5) doubts about actions, and (6) organization (130).

2.5.2.3.10 Interoceptive Sensitivity and Attention Questionnaire (ISAQ)
The ISAQ is a self-assessment tool designed to evaluate interoception using 17 items. Each item is scored on a 5-point scale, ranging from 1 (totally disagree) to 5 (totally agree). The ISAQ is composed of three subscales: (1) sensitivity to neutral bodily sensations, (2) attention to unpleasant bodily sensations, and (3) difficulty disengaging from unpleasant bodily sensations (131).

2.5.2.3.11 The Medical Outcome Study Short Form – 36 items (SF-36)
Health-related quality of life will be measured by means of the SF-36. The questionnaire contains a series of subscales covering different dimensions of perceived health: physical functioning, social functioning, role limitations due to physical health, role limitations due to emotional problems, bodily pain, emotional wellbeing, energy/fatigue, health change and general health perceptions (113). A Physical Composite Summary and Mental Composite Summary score will be calculated (117, 118).

2.5.2.3.12 Need for Controllability and Predictability questionnaire (NCPQ)
An individual's need for control and predictability is assessed via the 15-item NCPQ. Responders need to rate how much the item applies to them by giving a score between 1 'not typical' and 5 'very typical' (132).

2.5.2.3.13 COVID-related questions
Data will be collected about (1) date of COVID infection(s), (2) presence and severity of symptoms during and after the COVID infections (a.o., neurologic, psychiatric, neuroendocrinological, ANS and immune-related) on a NRS from 0: not at all to 10: extreme/ unbearable, (3) physical condition and health before and after COVID, (4) subjective severity of the COVID infection(s), (5) influence of the COVID infection(s) on daily life now, and (6) date and type of COVID vaccination.

**2.6 Neuroimaging data**
2.6.1 Preprocessing
2.6.1.0 Anatomical data MR
Mriqc version 0.16.0 will be used for quality control purposes. Runs will be excluded when (a) >15% of the volumes identified as spikes based on the FD and DVARS thresholds set in fMRIprep (details below) (b) Less than 2 fMRI MIST runs are available. In addition, the EPI signal will be visually inspected to check for signal dropout or other anatomical abnormalities that could compromise data quality.

Preprocessing will be performed using fMRIPrep 20.2.6 (RRID:SCR_016216) (30, 31) which is based on Nipype 1.7.0 (RRID:SCR_002502) (33, 34). Many internal operations of fMRIPrep use Nilearn 0.6.2 (RRID:SCR_001362) (26), mostly within the functional processing workflow. The fMRIPrep pipeline uses a combination of tools from well-known software packages, including FSL, ANTs, FreeSurfer and AFNI. This pipeline was designed to provide the best software implementation for each state of preprocessing and will be updated as newer and better neuroimaging software become available. Visual inspection of the data and fMRIPrep reports will be performed at each stage to ensure the quality of the data. For more details of the pipeline can be found here.

A total of 1 T1-weighted (T1w) images will be found within the input BIDS dataset. The T1-weighted (T1w) image will be corrected for intensity non-uniformity (INU) with N4BiasFieldCorrection (1), distributed with ANTs 2.3.3 ( RRID:SCR_004757) (27), and used as T1w-reference throughout the workflow. The T1w-reference will be then skull-stripped with a Nipype implementation of the antsBrainExtraction.sh workflow (from ANTs), using OASIS30ANTs as target template. Brain tissue segmentation of cerebrospinal fluid (CSF), white-matter (WM) and gray-matter (GM) will be performed on the brain-extracted T1w using fast (FSL 5.0.9, RRID:SCR_002823) (44). Volume-based spatial normalization to one standard space (MNI152NLin2009cAsym) will be performed through nonlinear registration with antsRegistration (ANTs 2.3.3), using brain-extracted versions of both T1w reference and the T1w template. The following template will be selected for spatial normalization: ICBM 152 Nonlinear Asymmetrical template version 2009c [RRID:SCR_008796; TemplateFlow ID: MNI152NLin2009cAsym] (32).

2.6.1.1 Task-based fMRI (Montreal Imaging Stress Test)
Mriqc version 0.16.0 will be used for quality control purposes. Runs will be excluded when (a) >15% of the volumes identified as spikes based on the FD and DVARS thresholds set in fMRIprep (details below) (b) Less than 2 fMRI MIST runs are available. In addition, the EPI signal will be visually inspected to check for signal dropout or other anatomical abnormalities that could compromise data quality.

Preprocessing will be performed using fMRIPrep 20.2.6 (RRID:SCR_016216) (30, 31) which is based on Nipype 1.7.0 (RRID:SCR_002502) (33, 34). Many internal operations of fMRIPrep use Nilearn 0.6.2 (RRID:SCR_001362) (26), mostly within the functional processing workflow. The fMRIPrep pipeline uses a combination of tools from well-known software packages, including FSL, ANTs, FreeSurfer and AFNI. This pipeline was designed to provide the best software implementation for each state of preprocessing and will be updated as newer and better neuroimaging software become available. Visual inspection of the data and fMRIPrep reports will be performed at each stage to ensure the quality of the data. More details of the pipeline can be found here.

For each of the 4 BOLD runs found per subject (4 task-based MIST- runs), the following preprocessing will be performed. First, a reference volume and its skull-stripped version will be generated using a custom methodology of fMRIPrep. A deformation field to correct for susceptibility distortions will be estimated based on fMRIPrep's fieldmap-less approach. The deformation field is that resulting from co-registering the BOLD reference to the same-subject T1w-reference with its intensity inverted (36, 43). Registration is performed with antsRegistration (ANTs 2.3.3), and the process regularized by constraining deformation to be nonzero only along the phase-encoding direction, and modulated with an average fieldmap template (42). Based on the estimated susceptibility distortion, a corrected EPI (echo-planar imaging) reference will be calculated for a more accurate co-registration with the anatomical reference. The BOLD reference will then be co-registered to the T1w reference using flirt (FSL 5.0.9) (38) with the boundary-based registration (35) cost-function. Co-registration will be configured with nine degrees of freedom to account for distortions remaining in the BOLD reference. Head-motion parameters with respect to the BOLD reference (transformation matrices, and six corresponding rotation and translation parameters) are estimated before any spatiotemporal filtering using mcflirt (FSL 5.0.9) (37). BOLD runs will be slice-time corrected to 0.476s (0.5 of slice acquisition range 0s-0.952s) using 3dTshift from AFNI 20160207 (RRID:SCR_005927) (29). The BOLD time-series (including slice-timing correction when applied) will be resampled onto their original, native space by applying a single, composite transform to correct for head-motion and susceptibility distortions. These resampled BOLD time-series will be referred to as preprocessed BOLD in original space, or just preprocessed BOLD. The BOLD time-series will be resampled into standard space, generating a preprocessed BOLD run in MNI152NLin2009cAsym space. First, a reference volume and its skull-stripped version will be generated using a custom methodology of fMRIPrep. Several confounding time-series will be calculated based on the preprocessed BOLD: framewise displacement (FD), DVARS and three region-wise global signals. FD will be computed using two formulations following Power (absolute sum of relative motions) (40) and Jenkinson (relative root mean square displacement between affines) (37). FD and DVARS are calculated for each functional run, both using their implementations in Nipype (following the definitions by Power et al. (2014) (40)). The three global signals are extracted within the CSF, the WM, and the whole-brain masks. Additionally, a set of physiological regressors will be extracted to allow for component-based noise correction (CompCor) (17). Principal components are estimated after high-pass filtering the preprocessed BOLD time-series (using a discrete cosine filter with 128s cut-off) for the two CompCor variants: temporal (tCompCor) and anatomical (aCompCor). tCompCor components are then calculated from the top 2% variable voxels within the brain mask. For aCompCor, three probabilistic masks (CSF, WM and combined CSF+WM) are generated in anatomical space. The implementation differs from that of Behzadi et al. (2007) in that instead of eroding the masks by 2 pixels on BOLD space, the aCompCor masks are subtracted a mask of pixels that likely contain a volume fraction of GM. This mask is obtained by thresholding the corresponding partial volume map at 0.05, and it ensures components are not extracted from voxels containing a minimal fraction of GM. Finally, these masks are resampled into BOLD space and binarized by thresholding at 0.99 (as in the original implementation). Components are also calculated separately within the WM and CSF masks. For each CompCor decomposition, the k components with the largest singular values are retained, such that the retained components' time series are sufficient to explain 50 percent of variance across the nuisance mask (CSF, WM, combined, or temporal). The remaining components are dropped from consideration. The head-motion estimates calculated in the correction step will also be placed within the corresponding confounds file. The confound time series derived from head motion estimates and global signals will be expanded with the inclusion of temporal derivatives and quadratic terms for each (41). Frames that exceeded a threshold of 0.9 mm FD or 2.0 standardised DVARS will be annotated as motion outliers. All resamplings can be performed with a single interpolation step by composing all the pertinent transformations (i.e. head-motion transform matrices, susceptibility distortion correction when available, and co-registrations to anatomical and output spaces). Gridded (volumetric) resamplings will be performed using antsApplyTransforms (ANTs), configured with Lanczos interpolation to minimize the smoothing effects of other kernels (39). Non-gridded (surface) resamplings will be performed using mri_vol2surf (FreeSurfer).

2.6.1.2 Functional connectivity

Mriqc version 0.16.0 will be used for quality control purposes. Runs will be excluded when (a) >15% of the volumes identified as spikes based on the FD and DVARS thresholds set in fMRIprep (details below) (b) Less than 2 fMRI MIST runs are available. In addition, the EPI signal will be visually inspected to check for signal dropout or other anatomical abnormalities that could compromise data quality.

Preprocessing will be performed using fMRIPrep 20.2.6 (RRID:SCR_016216) (30, 31) which is based on Nipype 1.7.0 (RRID:SCR_002502) (33, 34). Many internal operations of fMRIPrep use Nilearn 0.6.2 (RRID:SCR_001362) (26), mostly within the functional processing workflow. The fMRIPrep pipeline uses a combination of tools from well-known software packages, including FSL, ANTs, FreeSurfer and AFNI. This pipeline was designed to provide the best software implementation for each state of preprocessing and will be updated as newer and better neuroimaging software become available. Visual inspection of the data and fMRIPrep reports will be performed at each stage to ensure the quality of the data. More details of the pipeline can be found here.

For each BOLD run found per subject (1 resting-state functional connectivity run), the following preprocessing will be performed. First, a reference volume and its skull-stripped version will be generated using a custom methodology of fMRIPrep. A deformation field to correct for susceptibility distortions will be estimated based on fMRIPrep's fieldmap-less approach. The deformation field is that resulting from co-registering the BOLD reference to the same-subject T1w-reference with its intensity inverted (36, 43). Registration is performed with antsRegistration (ANTs 2.3.3), and the process regularized by constraining deformation to be nonzero only along the phase-encoding direction, and modulated with an average fieldmap template (42). Based on the estimated susceptibility distortion, a corrected EPI (echo-planar imaging) reference will be calculated for a more accurate co-registration with the anatomical reference. The BOLD reference will then be co-registered to the T1w reference using flirt (FSL 5.0.9) (38) with the boundary-based registration (35) cost-function. Co-registration will be configured with nine degrees of freedom to account for distortions remaining in the BOLD reference. Head-motion parameters with respect to the BOLD reference (transformation matrices, and six corresponding rotation and translation parameters) are estimated before any spatiotemporal filtering using mcflirt (FSL 5.0.9) (37). BOLD runs will be slice-time corrected to 0.476s (0.5 of slice acquisition range 0s-0.952s) using 3dTshift from AFNI 20160207 (RRID:SCR_005927) (29). The BOLD time-series (including slice-timing correction when applied) will be resampled onto their original, native space by applying a single, composite transform to correct for head-motion and susceptibility distortions. These resampled BOLD time-series will be referred to as preprocessed BOLD in original space, or just preprocessed BOLD. The BOLD time-series will be resampled into standard space, generating a preprocessed BOLD run in MNI152NLin2009cAsym space. First, a reference volume and its skull-stripped version will be generated using a custom methodology of fMRIPrep. Several confounding time-series will be calculated based on the preprocessed BOLD: framewise displacement (FD), DVARS and three region-wise global signals. FD will be computed using two formulations following Power (absolute sum of relative motions) (40) and Jenkinson (relative root mean square displacement between affines) (37). FD and DVARS are calculated for each functional run, both using their implementations in Nipype (following the definitions by Power et al. (2014) (40)). The three global signals are extracted within the CSF, the WM, and the whole-brain masks. Additionally, a set of physiological regressors will be extracted to allow for component-based noise correction (CompCor) (17). Principal components are estimated after high-pass filtering the preprocessed BOLD time-series (using a discrete cosine filter with 128s cut-off) for the two CompCor variants: temporal (tCompCor) and anatomical (aCompCor). tCompCor components are then calculated from the top 2% variable voxels within the brain mask. For aCompCor, three probabilistic masks (CSF, WM and combined CSF+WM) are generated in anatomical space. The implementation differs from that of Behzadi et al. (2007) in that instead of eroding the masks by 2 pixels on BOLD space, the aCompCor masks are subtracted a mask of pixels that likely contain a volume fraction of GM. This mask is obtained by thresholding the corresponding partial volume map at 0.05, and it ensures components are not extracted from voxels containing a minimal fraction of GM. Finally, these masks are resampled into BOLD space and binarized by thresholding at 0.99 (as in the original implementation). Components are also calculated separately within the WM and CSF masks. For each CompCor decomposition, the k components with the largest singular values are retained, such that the retained components' time series are sufficient to explain 50 percent of variance across the nuisance mask (CSF, WM, combined, or temporal). The remaining components are dropped from consideration. The head-motion estimates calculated in the correction step will also be placed within the corresponding confounds file. The confound time series derived from head motion estimates and global signals will be expanded with the inclusion of temporal derivatives and quadratic terms for each (41). Frames that exceeded a threshold of 0.9 mm FD or 2.0 standardised DVARS will be annotated as motion outliers. All resamplings can be performed with a single interpolation step by composing all the pertinent transformations (i.e. head-motion transform matrices, susceptibility distortion correction when available, and co-registrations to anatomical and output spaces). Gridded (volumetric) resamplings will be performed using antsApplyTransforms (ANTs), configured with Lanczos interpolation to minimize the smoothing effects of other kernels (39). Non-gridded (surface) resamplings will be performed using mri_vol2surf (FreeSurfer). Functional data will be smoothed using spatial convolution with a Gaussian kernel of 6 mm full width half maximum (FWHM) using CONN (11) (RRID:SCR_009550) release 22.v2407 (12) and SPM (13) (RRID:SCR_007037) release 12.7771.

In addition, functional data will be denoised using a standard denoising pipeline (14) including the regression of potential confounding effects characterized by filtered white matter timeseries (5 CompCor noise components), filtered CSF timeseries (5 CompCor noise components), motion parameters and their first order derivatives (12 factors) (15), session effects and their first order derivatives (2 factors), QC_motion_outlier regressors and their first order derivatives (58 components), and linear trends (2 factors) within each functional run, followed by bandpass frequency filtering of the BOLD timeseries (16) between 0.008 Hz and 0.09 Hz. CompCor (17,18) noise components within white matter and CSF will be estimated by computing the average BOLD signal as well as the largest principal components orthogonal to the BOLD average, motion parameters within each subject's eroded segmentation masks. From the number of noise terms included in this denoising strategy, the effective degrees of freedom of the BOLD signal after denoising will be estimated to range from 59.9 to 69.4 (average 69) across all subjects (19).

2.6.1.3 Positron emission tomography (PET)

Source data from the scanner will be converted to Nifti format from DICOM format and organized into Brain Imaging Data Structure (BIDS) specification prior to preprocessing. After quality control (checking the alignment between PET and the attenuation correction image (ZTE) and adjusting the origin and orientation (to make it more similar to the orientation of the template in MNI space) of T1, and the dynamic PET images), a summed image of the first 5min of PET acquisition will be calculated and each pair of two sequential frames will be realigned starting from the summed image and the next frame after 5min without resampling images to avoid accumulation of resampling errors. Then, co-registration of the summed PET to the T1w image (and applied to all individual frames of the dynamic PET image) will be performed, followed by a segmentation of T1-w in gray matter (GM), white matter (WM), and cerebrospinal fluid (CSF) and this step will also generate the deformation fields to warp the images from patient space to MNI. The warping parameters will be applied to the PET, GM, WM and

CSF images. A quality check will be performed by evaluating head-motion parameters, checking the coregistration, checking quality of segmentation, and checking the warping of PET and MRI data. This part will be performed using SPM12 tools.

The volume of distribution ($V_T$) of [$^{18}$F]-DPA714 will be quantified using a plasma-input based 2 tissue 4K model with an additional component describing irreversible endothelial binding and taking into account an estimated fraction of whole blood in predefined ROIs and in parcels of the Canlab 2024 combined atlas (**Supplementary table 3)**. Parametric images of $V_T$ will be calculated using a Logan graphical analysis (46) with t* = 31min (112). Total distribution volume ($V_T$) is the primary outcome measure. For the ROI and parcel based analysis we will also estimate $K_1$ and use this as a proxy for blood-brain barrier integrity (47). To calculate the plasma input function corrected for the intact fraction, we will fit a constrained Hill function using the data of the intact fraction in 5 samples over time. ( in-house scripts)

2.6.1.4 Magnetic resonance spectroscopy (MRS)
Data will be processed in the Osprey Toolbox (98) in MATLAB following standardized procedures for preprocessing and linear-combination modeling, as outlined in consensus guidelines. Choline (CHO) and myo-inositol (MI) quantification will be performed as markers of neuroinflammation. Total N-Acetylaspartate (tNAA), a proxy marker of neuronal health (48), and glutamate/glutamine (Glx), amino acids important in neuronal regulation (49), concentrations will be assessed exploratorily. Metabolite concentrations (institutional units (i.u.)) will be water-scaled using the unsuppressed water as a reference signal. The next preprocessing steps will be followed: First, robust spectral registration (rSR) is performed. Its pipeline consists of (1) filtering lipid contamination and residual water signals, (2) reordering free induction decays based on their similarity, (3) frequency and phase correction in the time domain using spectral registration, (4) subspectral alignment in the frequency domain, (5) weighted averaging of data (99). After subsequent Eddy current correction, a wide metabolite dataset will be defined and modeled with Osprey, including myo-inositol (MI) and choline (CHO). For MRS data acquired on the 3T Philips Achieva DStream scanner, spectral alignment, weighted averaging, and eddy current correction are performed during scanner reconstruction. Therefore, these preprocessing steps were omitted from the offline rSR pipeline for data acquired on this system. The modeled spectral range will span from 0.5 to 4 ppm for the metabolites and from 2.0 to 7.0 for water, and a baseline knot spacing (0.4 ppm) will be used. Next, the voxel mask will be coregistered to the T1-w image. Segmentation will be performed via SPM12 functions to determine fractions of the different tissue types (GM, WM and CSF) to account for tissue-specific effects of relaxation, tissue water content, and metabolite content. This is necessary as CHO and MI are shown to be significantly higher in gray compared to white matter (51, 52). Data will be quantified as [met]TissCorr (Gasparovic-method) using the unsuppressed water signal as a reference.

The quality parameters that will be applied are

(1) Visual inspection of each spectrum, and exclusion of any subject showing clear artifacts, like out-of-voxel echoes (OOVs), to ensure data quality.
(2) Calculation of the signal-to-noise ratio (SNR) to determine signal strength relative to background noise. Participants with SNR more than 3 standard deviations below the mean (calculated separately for both scanners used in the study) will be excluded.
(3) Checking the linewidth FWHM.

(4) Checking the voxel placement and voxel fractions. Participants in which the voxel contained a fraction of cerebrospinal fluid more than 3 standard deviations above the mean (calculated separately for both scanners used in the study) will be excluded.

2.6.2 Region of interest (ROI) definition

2.6.2.1 Task-based fMRI (Montreal Imaging Stress Test)

A brain region was included as an ROI if it was reported as significant (significant (de)activation during stress vs control/baseline) in more than 60% of relevant stress-related brain imaging meta-analyses (53, 54, 55, 56), including automated meta-analysis from Neurosynth. The following bilateral regions will be extracted from the Canlab 2024 atlas as ROIs for MIST analysis: anterior insula, ventrolateral prefrontal cortex, amygdala, and thalamus (**Supplementary table 4**).

2.6.2.2 Functional connectivity

To investigate resting-state functional connectivity (rs-FC), ROIs will be defined based on previous case-control and intervention studies in ME/CFS populations (57-63, 95). A brain region was included as an ROI if at least two studies reported significant rs-FC alterations in that region. The following bilateral ROIs will be extracted from the CANLab 2024 atlas to be used in rs-FC analysis: anterior mid-cingulate cortex, frontal pole, anterior and posterior insula, premotor cortex, somatomotor paracentral area, dorsolateral and dorsomedial prefrontal cortex, primary motor cortex (M1), primary somatosensory cortex (S1), posterior cingulate cortex/precuneus, superior and inferior parietal lobes, temporoparietal-occipital junction, auditory association cortex, middle temporal gyrus, globus pallidus, thalamus, and caudate (**Supplementary table 5**).

2.6.2.3 PET

To examine neuroinflammation, ROIs were selected based on prior translocator protein (TSPO) PET studies in affective and functional somatic disorders (64), including major depressive disorder (65-72), fibromyalgia (73, 74), chronic pain (3, 6, 7, 75-77), ME/CFS (5) and functional somatic syndromes (4). A region was included as an ROI if it was examined in at least three studies and reported as significantly different between patients and controls in at least 75% of those studies (i.e., frequency(significant)/frequency(defined) > 75%). The following bilateral ROIs will be extracted from the Canlab 2024 atlas: anterior mid-cingulate cortex, subgenual anterior cingulate cortex, anterior insula, posterior insula, hippocampus, S1, and thalamus (**Supplementary tables 6&7**). ROI analysis will be performed complementary to thresholded whole-brain analysis.

**2.7 Statistical analysis plan**

2.7.1 Objective 1: compare the neuropsychophysiological mechanisms between ME/CFS patients and healthy participants

2.7.1.2 Gut microbiome

Samples will be grouped into community types via the Dirichlet multinomial mixtures method as described by Holmes, Harris, Quince (2012) (103). To generate quantitative microbiome profiles (QMP), we will integrate sequencing data with microbial load estimates obtained through flow cytometry, as described in recent literature (104). This integration enhances interpretability into microbiome dynamics by capturing both the magnitude and direction of changes, while also overcoming the limitations associated with compositional data analysis, inherent in methods like *coda.base* and *robCompositions*. Where relevant, multiple testing corrections will be applied using the (positive) FDR method.

2.7.1.3 Brain task-based fMRI

Regarding the brain data, we will determine brain activation during the MIST by calculating the stress minus control contrast at the subject level (i.e., first level based on the general linear model (GLM) using custom scripts calling functions from the CANab toolbox and SPM12 (Statistical Parametric Mapping) in MATLAB. Using the beta images of these first-level contrasts, second-level fixed-effects analysis, comparing patients and healthy participants, will be conducted using GLMs implemented in MATLAB scripts calling CANlab and SPM12 functions. Whole-brain voxel-wise GLMs using a gray matter mask will be performed, using a voxel-wise threshold of $q_{FDR} < 0.05$. The whole-brain analyses will be complemented by ROI-based (M)ANOVAs with positive FDR correction ($q_{FDR} < 0.05$). (see "2.6.2 Region of interest (ROI) definition" for ROI selection and justification). GLM analyses will be corrected for scanner type (MR vs. PET/MR) (fixed effect). Whole brain multivariate mediation analysis will be performed using the "principal directions of mediation" (PDM) method (144, 145) in order to study brain patterns mediating case-control differences in self-reported NA during the MIST. Analyses will be done using the CANlab mediation toolbox and will be thresholded voxel-wise at $q_{FDR} < 0.05$. Finally, multi-voxel pattern analysis (MVPA) will be employed with The Decoding Toolbox (115) to train and cross-validate (k-fold) a Support Vector Machines classifier to distinguish ME/CFS patients from healthy controls based on their brain response to stress. Given the group size imbalance, the classification performance will be evaluated using the area under the curve (AUC), which is more robust for the imbalanced dataset than the commonly used accuracy metric (114). The searchlight procedure will be conducted within the grey matter, using an 8 mm-radius sphere, and AUC values will be assigned to the central voxel. Statistical significance (voxel-wise p-values) will be estimated using 5,000 permutation tests, preserving group distribution in each fold during the label shuffling.

2.7.1.4 Brain resting-state fMRI

2.7.1.4.1 ROI-to-ROI connectivity analyses

2.7.1.4.1.1 Multivariate GLM

2.7.1.4.1.1.1 R2R

ROI-to-ROI connectivity (RRC) matrices will be estimated characterizing the functional connectivity between each pair of regions among 38 ROIs. Functional connectivity strength will be represented by Fisher-transformed bivariate correlation coefficients from a general linear model (weighted-GLM (20)), estimated separately for each pair of ROIs, characterizing the association between their BOLD signal timeseries. Group-level analyses will be performed using a General Linear Model (GLM (21)). For each individual connection a separate GLM will be estimated, with first-level connectivity measures at this connection as dependent variables (one independent sample per subject), and group as independent variable. Connection-level hypotheses will be evaluated using multivariate parametric statistics with random-effects across subjects and sample covariance estimation across multiple measurements. Inferences will be performed at the level of individual ROIs. ROI-level inferences will be based on parametric multivariate statistics, combining the connection-level statistics across all connections from each individual ROI. Results will be thresholded using a combination of uncorrected $p < 0.01$ connection-level threshold and familywise error correction ($p_{FWE} < 0.05$) at ROI-level (ROI mass/intensity).

2.7.1.4.1.1.2 R2R Networks

ROI-to-ROI connectivity (RRC) matrices will be estimated characterizing the functional connectivity between each pair of regions among 32 High-Performance Computing-Independent Component Analysis (HPC-ICA) network ROIs (12). Functional connectivity strength will be represented by Fisher-transformed bivariate correlation coefficients from a general linear model (weighted-GLM (20)), estimated separately for each pair of ROIs, characterizing the association between their BOLD signal timeseries. Group-level analyses will be performed using a General Linear Model (GLM (21)). For each individual connection a separate GLM will be estimated, with first-level connectivity measures at this connection as dependent variables (one independent sample per subject), and group as independent variable. Connection-level hypotheses will be evaluated using multivariate parametric statistics with random-effects across subjects and sample covariance estimation across multiple measurements. Inferences will be performed at the level of individual clusters (groups of similar connections). Cluster-level inferences will be based on parametric statistics within- and between- each pair of networks (Functional Network Connectivity (22)), with networks identified using a complete-linkage hierarchical clustering procedure (23) based on ROI-to-ROI anatomical proximity and functional similarity metrics (24). Results will be thresholded using a combination of a $p < 0.05$ connection-level threshold and a $p_{FWE} < 0.05$ ROI-level threshold (ROI mass/intensity).

2.7.1.4.1.2 Graph Theoretical Analysis

First, functional connectivity networks (i.e. ROI-to-ROI connectivity matrices) will be constructed using the Group Sparse Representation method (101), a multi-region construction technique designed to avoid spurious correlations and to enhance classification model performance, implemented in the BrainNetClass toolbox (116) and built into GraphVar. Raw connectivity matrices (relative thresholding at 0.2, 0.25, 0.30, 0.35, and 0.4 density thresholds representing the 20%, 25%, 30%, 35%, and 40% strongest connections, to test robustness of findings across different thresholds; threshold values based on lower thresholds resulting in fragmented networks based on fragmentation check in GraphVar) will be used to compute local and global graph measures: 1) clustering component (nodal and global), 2) average path length (nodal and global), 3) nodal betweenness centrality, 4) efficiency (nodal and global), and 5) assortativity (global). Group comparisons for the graph measures will be performed with one-way ANOVAs (for non-density-dependent graph measures) and MANOVAs (for density-dependent graph measures with group as independent factor and density as repeated factor). Positive FDR correction ($q_{FDR} < 0.05$) will be applied (25, 100) (i.e. when the estimated number of true significances by the bootstrap and/or spline procedure > 0), otherwise standard Benjamini-Hochberg FDR will be used (102). Furthermore, Partial Least Squares (PLS) Discriminant Analysis will be implemented to test how well the graph measures are able to distinguish ME/CFS patients from healthy participants. Nonlinear iterative partial least squares (NIPALS) method implemented in 2025 JMP® Pro 18 software will be used for this as it operates effectively when the X variables are highly associated and when there are more X values than observations because of latent factor creation. N-fold cross-validation based on minimum root mean PRESS, Van der Voet $T^2$ statistics and maximal values for $Q^2$ will be applied to decide on the most suitable number of factors while preventing overfitting of the data (8,9). Because of the imbalanced classification caused by the unequal patient/healthy participant ratio, the Tomek Sampling method (10) will be applied in JMP.

2.7.1.4.2 Functional Connectivity Multivariate Pattern Analysis (fc-MVPA)

Functional connectivity multivariate pattern analyses (fc-MVPA (20)) will be performed to estimate 6 eigenpatterns characterizing the principal axes of heterogeneity in functional connectivity across subjects. From these eigenpatterns, 6 associated eigenpattern-score images will be derived for each individual subject characterizing their brain-wide functional connectome state. Eigenpatterns and eigenpattern-scores will be computed separately for each individual seed voxel as the left- and right-singular vectors, respectively, from a singular value decomposition (group-level SVD) of the matrix of functional connectivity values between this seed voxel and the rest of the brain (a matrix with one row per target voxel, and one column per subject). Individual functional connectivity values will be computed from the matrices of bivariate correlation coefficients between the BOLD timeseries from each pair of voxels, estimated using a singular value decomposition of the z-score normalized BOLD signal (subject-level SVD) with 64 components separately for each subject (11). Group-level analyses will be performed using a General Linear Model (GLM (21)). For each individual voxel a separate GLM will be estimated, with first-level connectivity measures at this voxel as dependent variables (one independent sample per subject), and group as independent variable in an F-test across the 6 estimated eigenpatterns. Voxel-level hypotheses will be evaluated using multivariate parametric statistics with random-effects across subjects and sample covariance estimation across multiple measurements. Inferences will be performed at the level of individual clusters (groups of contiguous voxels). Cluster-level inferences will be based on nonparametric statistics using Threshold Free Cluster Enhancement (TFCE), with 5000 residual-randomization iterations (22,23). Results will be familywise error corrected ($p_{FWE} < 0.05$).

### Supplementary Tables and Figures

| Observed variables | | Measured at baseline | Measured at follow-up 1 | Measured at follow-up 2 |
|---|---|---|---|---|
| Stress-response system (SRS) | HRV in rest | 1 | | 1 |
| | HRV during arithmetic task | 1 | | 1 |
| | 8 cortisol outcomes during MAST | 1 | | 1 |
| | 4 subjective stress measurements during MAST | 1 | | 1 |
| | 5 subjective stress measurements during MIST | 1 | | |
| | Wearable device physiological measurements | 1 | | 1 |
| | ESM items about stress, nervosity, arousal and valence | 1 | 1 | 1 |
| Immune system | TNF-α, IL-1β, IL-4, IL-6, IL-8, IL-10 and IFN-γ | 1 | | 1 |
| | hsCRP | 1 | | 1 |
| Brain | BOLD response during MIST | 1 | | |
| | rCBF | 1 | | |
| | Distribution volume (VT), uptake rate constant (K1) | 1 | | |
| | rsFC | 1 | | |
| | MRS | 1 | | |
| Gut | Gut-microbiota composition | 1 | | 1 |
| | SCFA | 1 | | 1 |
| Fatigue and fatigability | 6 FES scores on physical fatigue | 1 | | 1 |
| | 8 FES scores on mental fatigue | 1 | | 1 |
| | ESM items about mental fatigue, physical fatigue, sleep quality, activities and rest | 1 | 1 | 1 |
| | Scores on CIS-20 | 1 | 1 | 1 |

Supplementary Table 1: Overview variables included. HRV= heart rate variability, MAST= Maastricht Acute Stress Task, PASAT= Paced Auditory Serial Attention Task, SCFA= short-chain fatty acids, FES=fatigue and energy scale, ESM=experience sampling method, MIST= Montreal Imaging Stress Task, hsCRP=high-sensitivity C-reactive protein, BOLD=blood oxygen level dependent, rCBF=regional cerebral blood flow, VT=distribution volume, K1=uptake rate constant, rsFC=resting-state functional connectivity, MRS=magnetic resonance spectroscopy, CIS-20=Checklist individual strength

| Subject | Questions | Answer options | Reference article |
|---|---|---|---|
| **Morning questions** | | | |
| Sleep quality | At what time did you go to sleep yesterday? | Open question | (50,78, 79) |
| | At what time did you wake up this morning? | Open question | |
| | How long did you sleep last night? | Open question | |
| | This night I experienced problems with | Multiple choice: 1) Falling asleep, 2) Sleeping through the night, 3) Both, 4) Neither | |
| | When I woke up, I felt rested | NRS ranging from 0 'not at all' to 100 'absolute maximum' | |
| | After waking up, for how long did you stay in bed before getting up? | Open question | |
| **Daily questions** | | | |
| Physical fatigue | At the moment I feel physically drained | NRS ranging from 0 'none' to 10 'absolute maximum' | (80) |
| | At the moment I feel physically tired | NRS ranging from 0 'none' to 10 'absolute maximum' | |
| | At the moment I feel heaviness in the legs | NRS ranging from 0 'none' to 10 'absolute maximum' | |
| | My physical energy is | NRS ranging from 0 'none' to 10 'absolute maximum' | |
| Mental fatigue | At the moment I experience brain fog | NRS ranging from 0 'none' to 10 'absolute maximum' | |
| | At the moment I experience concentration problems | NRS ranging from 0 'none' to 10 'absolute maximum' | |
| | At the moment I feel disoriented | NRS ranging from 0 'none' to 10 'absolute maximum' | |
| | At the moment I feel mentally drained | NRS ranging from 0 'none' to 10 'absolute maximum' | |
| | At the moment I feel mentally tired | NRS ranging from 0 'none' to 10 'absolute maximum' | |
| | At the moment I experience memory problems | NRS ranging from 0 'none' to 10 'absolute maximum' | |
| Symptom focusing | Since the last beep I worried about the fatigue | NRS ranging from 0 'not at all' to 100 'absolute maximum' | (81) |
| | Since the last beep I directed my attention towards the fatigue | NRS ranging from 0 'not at all' to 100 'absolute maximum' | |

| | | | |
|---|---|---|---|
| Avoidance of activities to prevent symptoms | Since the last beep, I avoided a certain activity to avoid feeling drained | NRS ranging from 0 'not at all' to 100 'absolute maximum' | (81) |
| Valence | Choose the first manikin if you feel completely happy. Choose the last manikin if you feel completely unhappy. Choose another option if your feelings are somewhere in between the two extremes, | 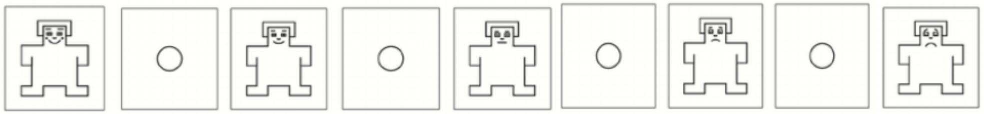 Multiple choice with inserted media | (82, 83, 84) |
| Arousal | Choose the first manikin if you feel completely calm. Choose the last manikin if you feel completely aroused. Choose another option if your feelings are somewhere in between the two extremes, | 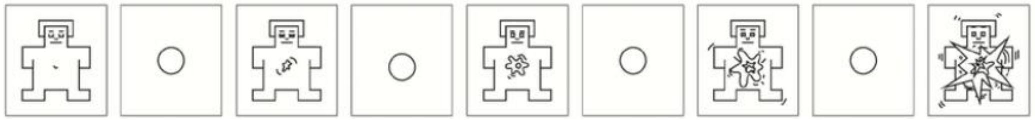 Multiple choice with inserted media | |
| Stress and nervosity | At the moment I feel stressed | NRS ranging from 0 'not at all' to 100 'absolute maximum' | (85, 86) |
| | At the moment I feel nervous | NRS ranging from 0 'not at all' to 100 'absolute maximum' | |
| | I feel comfortable in the environment in which I am right now | NRS ranging from 0 'not at all' to 100 'absolute maximum' | (87) |
| Activity | Five minutes or less before the beep I performed | Multiple choice: 1) No physical activity, 2) Light physical activity, 3) Moderate physical activity, 4) Strenuous physical activity | (88) |
| | Five minutes or less before the beep I performed mental effort | NRS ranging from 0 'not at all' to 100 'absolute maximum' | (81) |
| | Think about what you were doing 5 minutes or less before the beep: | | (89) |
| | This activity required effort | NRS ranging from 0 'not at all' to 100 'absolute maximum' | |
| | I rather did something else | NRS ranging from 0 'not at all' to 100 'absolute maximum' | |
| | I enjoyed this activity | NRS ranging from 0 'not at all' to 100 'absolute maximum' | (90) |

| | | | |
|---|---|---|---|
| Rest | Did you rest since the last beep? | Multiple choice: 1) Yes, 2) No | (88) |
| Symptoms | At the moment I suffer from shortness of breath | NRS ranging from 0 'not at all' to 100 'absolute maximum' | (91,133) |
| | At the moment I have a faster heart beat | NRS ranging from 0 'not at all' to 100 'absolute maximum' | |
| | At the moment I have a headache | NRS ranging from 0 'not at all' to 100 'absolute maximum' | |
| | At the moment I have muscle or joint pain | NRS ranging from 0 'not at all' to 100 'absolute maximum' | |
| Substance use | Since the last beep I used | Multiple choice: 1) Caffeine, 2) Nicotine, 3) Alcohol, 4) Medication, 5) Drugs, 6) Food, 7) None of the above | (92) |
| | Evening questions | | |
| Fatigue | Today I felt rested | NRS ranging from 0 'not at all' to 100 'absolute maximum' | (80) |
| | Today I felt mentally exhausted | NRS ranging from 0 'not at all' to 100 'absolute maximum' | |
| | Today I felt physically exhausted | NRS ranging from 0 'not at all' to 100 'absolute maximum' | |
| | Did you nap today? | Multiple choice: 1) Yes, before noon, 2) Yes, around noon, 3) Yes, after noon, 4) Yes, in the evening, 5) No, I did not nap today | (88) |
| Arousal | I am glad with how the day went | NRS ranging from 0 'not at all' to 100 'absolute maximum' | (93) |
| **Debriefing** | | | |
| | Was the past week a normal week? | NRS ranging from 0 to 100 | (94) |
| | Were you able to express your experiences via the questionnaires? | NRS ranging from 0 to 100 | |
| | Did the ESM influence your mood? | NRS ranging from 0 to 100 | |
| | Did the ESM period influence your routine throughout the day? | NRS ranging from 0 to 100 | |
| | Did the ESM period influence your daily contact with other people? | NRS ranging from 0 to 100 | |
| | Did any special events occur during the week? | Multiple choice: 1) Yes, 2) No | |
| | What kind of events occurred during the week? | Open questions | |

| | | |
|---|---|---|
| | Did you have problems with the wearable? | NRS ranging from 0 to 100 |

Supplementary Table 2: Ecological measurements overview questions and answer options

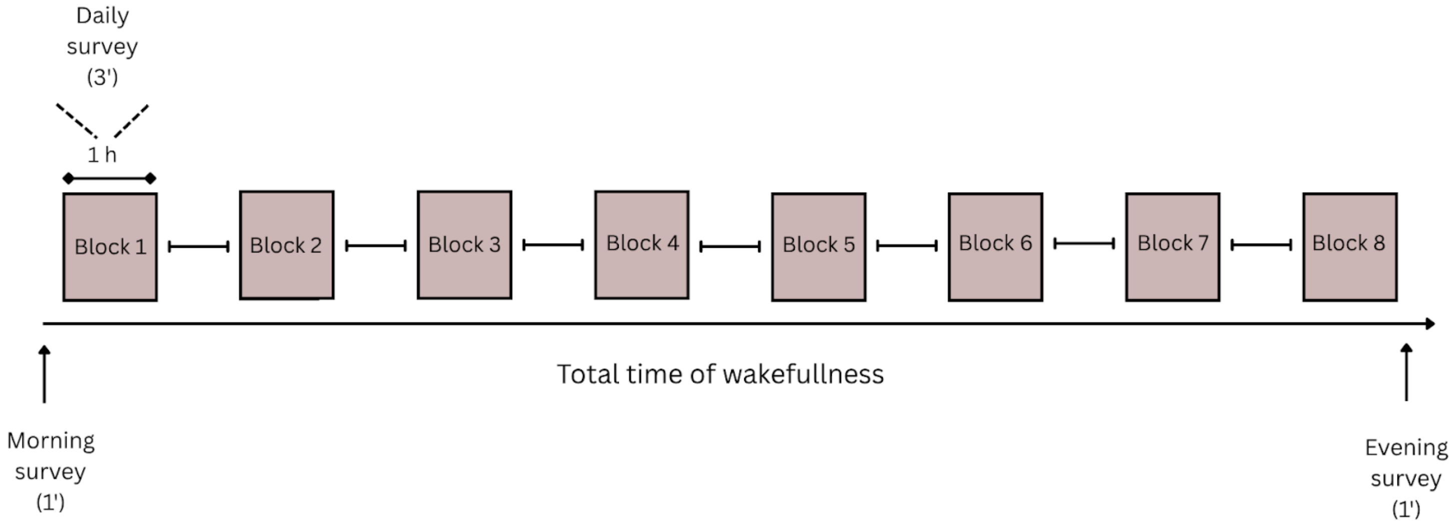

Supplementary Figure 1. Blocks 1 to 8 represent the 8 one-hour blocks during which the surveys will be sent. The blocks will be evenly distributed across the participant's entire daily wakefulness period.

| index | label descriptions | labels | labels granularity 2 | labels granularity 3 | labels granularity 4 | labels_5 |
|---|---|---|---|---|---|---|
| 1 | Primary visual cortex (Left) | Ctx_V1_L | Ctx_V1_L | striate_L | visual_early_L | Glasser2016 (Petre2023 volumetric projection) |
| 2 | Medial superior temporal area (Left) | Ctx_MST_L | Ctx_MST_L | dMT+_L | visual_MT+_L | Glasser2016 (Petre2023 volumetric projection) |
| 3 | sixth visual area (Left) | Ctx_V6_L | Ctx_V6_L | V6_L | visual_dorsal_L | Glasser2016 (Petre2023 volumetric projection) |
| 4 | second visual area (Left) | Ctx_V2_L | Ctx_V2_L | extrastriate_L | visual_early_L | Glasser2016 (Petre2023 volumetric projection) |
| 5 | third visual area (Left) | Ctx_V3_L | Ctx_V3_L | extrastriate_L | visual_early_L | Glasser2016 (Petre2023 volumetric projection) |
| 6 | fourth visual area (Left) | Ctx_V4_L | Ctx_V4_L | extrastriate_L | visual_early_L | Glasser2016 (Petre2023 volumetric projection) |
| 7 | eighth visual area (Left) | Ctx_V8_L | Ctx_V8_L | VVC_L | visual_ventral_L | Glasser2016 (Petre2023 volumetric projection) |
| 8 | primary motor cortex (Left) | Ctx_4_L | Ctx_4_L | M1_L | somatomotor_primary_L | Glasser2016 (Petre2023 volumetric projection) |

| | | | | | | |
|---|---|---|---|---|---|---|
| 9 | primary sensory cortex (Left) | Ctx_3b_L | Ctx_3b_L | S1_L | somatomotor_primary_L | Glasser2016 (Petre2023 volumetric projection) |
| 10 | frontal eye fields (Left) | Ctx_FEF_L | Ctx_FEF_L | superior_premotor_L | somatomotor_premotor_L | Glasser2016 (Petre2023 volumetric projection) |
| 11 | premotor eye field (Left) | Ctx_PEF_L | Ctx_PEF_L | inferior_premotor_L | somatomotor_premotor_L | Glasser2016 (Petre2023 volumetric projection) |
| 12 | area 55b (Left) | Ctx_55b_L | Ctx_55b_L | 55b_L | somatomotor_premotor_L | Glasser2016 (Petre2023 volumetric projection) |
| 13 | area V3A (Left) | Ctx_V3A_L | Ctx_V3A_L | supplementary_V3_L | visual_dorsal_L | Glasser2016 (Petre2023 volumetric projection) |
| 14 | RetroSplenial Complex (Left) | Ctx_RSC_L | Ctx_RSC_L | RSC_L | cingulate_posterior_L | Glasser2016 (Petre2023 volumetric projection) |
| 15 | Parieto-Occipital Sulcus Area 2 (Left) | Ctx_POS2_L | Ctx_POS2_L | parietal_occipital_sulcus_L | cingulate_posterior_L | Glasser2016 (Petre2023 volumetric projection) |
| 16 | Seventh Visual Area (Left) | Ctx_V7_L | Ctx_V7_L | V7_L | visual_dorsal_L | Glasser2016 (Petre2023 volumetric projection) |
| 17 | IntraParietal Sulcus Area 1 (Left) | Ctx_IPS1_L | Ctx_IPS1_L | ISP1_L | visual_dorsal_L | Glasser2016 (Petre2023 |

| | | | | | | |
|---|---|---|---|---|---|---|
| | | | | | | volumetric projection) |
| 18 | Fusiform Face Complex (Left) | Ctx_FFC_L | Ctx_FFC_L | FFC_L | visual_ventral_L | Glasser2016 (Petre2023 volumetric projection) |
| 19 | Area V3B (Left) | Ctx_V3B_L | Ctx_V3B_L | supplementary_V3_L | visual_dorsal_L | Glasser2016 (Petre2023 volumetric projection) |
| 20 | Area Lateral Occipital 1 (Left) | Ctx_LO1_L | Ctx_LO1_L | LO_L | visual_MT+_L | Glasser2016 (Petre2023 volumetric projection) |
| 21 | Area Lateral Occipital 2 (Left) | Ctx_LO2_L | Ctx_LO2_L | LO_L | visual_MT+_L | Glasser2016 (Petre2023 volumetric projection) |
| 22 | Posterior InferoTemporal (Left) | Ctx_PIT_L | Ctx_PIT_L | FFC_L | visual_ventral_L | Glasser2016 (Petre2023 volumetric projection) |
| 23 | Middle Temporal Area (Left) | Ctx_MT_L | Ctx_MT_L | dMT+_L | visual_MT+_L | Glasser2016 (Petre2023 volumetric projection) |
| 24 | Primary Auditory Cortex (Left) | Ctx_A1_L | Ctx_A1_L | A1_L | auditory_early_L | Glasser2016 (Petre2023 volumetric projection) |
| 25 | PeriSylvian Language Area (Left) | Ctx_PSL_L | Ctx_PSL_L | PSL_L | parietal_TPOJ_L | Glasser2016 (Petre2023 volumetric projection) |

| | | | | | | |
|---|---|---|---|---|---|---|
| 26 | Superior Frontal Language Area (Left) | Ctx_SFL_L | Ctx_SFL_L | Superior_lateral_BA8_L | cingulate_dlPFC_L | Glasser2016 (Petre2023 volumetric projection) |
| 27 | PreCuneus Visual Area (Left) | Ctx_PCV_L | Ctx_PCV_L | PCV_L | cingulate_posterior_L | Glasser2016 (Petre2023 volumetric projection) |
| 28 | Superior Temporal Visual Area (Left) | Ctx_STV_L | Ctx_STV_L | STV_L | parietal_TPOJ_L | Glasser2016 (Petre2023 volumetric projection) |
| 29 | Medial Area 7P (Left) | Ctx_7Pm_L | Ctx_7Pm_L | posterior_BA7_L | parietal_superior_lobule_L | Glasser2016 (Petre2023 volumetric projection) |
| 30 | Area 7m (Left) | Ctx_7m_L | Ctx_7m_L | precuneus_L | cingulate_posterior_L | Glasser2016 (Petre2023 volumetric projection) |
| 31 | Parieto-Occipital Sulcus Area 1 (Left) | Ctx_POS1_L | Ctx_POS1_L | parietal_occipital_sulcus_L | cingulate_posterior_L | Glasser2016 (Petre2023 volumetric projection) |
| 32 | Area 23d (Left) | Ctx_23d_L | Ctx_23d_L | anterior_precuneus_L | cingulate_posterior_L | Glasser2016 (Petre2023 volumetric projection) |
| 33 | Area ventral 23 a+b (Left) | Ctx_v23ab_L | Ctx_v23ab_L | precuneus_L | cingulate_posterior_L | Glasser2016 (Petre2023 volumetric projection) |
| 34 | Area dorsal 23 a+ (Left) | Ctx_d23ab_L | Ctx_d23ab_L | precuneus_L | cingulate_posterior_L | Glasser2016 (Petre2023 |

| | | | | | | |
|---|---|---|---|---|---|---|
| | | | | | | volumetric projection) |
| 35 | Area 31p ventral (Left) | Ctx_31pv_L | Ctx_31pv_L | precuneus_L | cingulate_posterior_L | Glasser2016 (Petre2023 volumetric projection) |
| 36 | Area 5m (Left) | Ctx_5m_L | Ctx_5m_L | BA5_L | somatomotor_paracentral_lobule_L | Glasser2016 (Petre2023 volumetric projection) |
| 37 | Area 5m ventral (Left) | Ctx_5mv_L | Ctx_5mv_L | BA5_L | somatomotor_paracentral_lobule_L | Glasser2016 (Petre2023 volumetric projection) |
| 38 | Area 23c (Left) | Ctx_23c_L | Ctx_23c_L | anterior_precuneus_L | cingulate_posterior_L | Glasser2016 (Petre2023 volumetric projection) |
| 39 | Area 5L (Left) | Ctx_5L_L | Ctx_5L_L | BA5_L | somatomotor_paracentral_lobule_L | Glasser2016 (Petre2023 volumetric projection) |
| 40 | Dorsal Area 24d (Left) | Ctx_24dd_L | Ctx_24dd_L | cingulate_motor_area_L | somatomotor_paracentral_lobule_L | Glasser2016 (Petre2023 volumetric projection) |
| 41 | Ventral Area 24d (Left) | Ctx_24dv_L | Ctx_24dv_L | cingulate_motor_area_L | somatomotor_paracentral_lobule_L | Glasser2016 (Petre2023 volumetric projection) |
| 42 | Lateral Area 7A (Left) | Ctx_7AL_L | Ctx_7AL_L | anterior_BA7_L | parietal_superior_lobule_L | Glasser2016 (Petre2023 volumetric projection) |

| | | | | | | |
|---|---|---|---|---|---|---|
| 43 | Supplementary and Cingulate Eye Field (Left) | Ctx_SCEF_L | Ctx_SCEF_L | SMA_L | somatomotor_paracentral_lobule_L | Glasser2016 (Petre2023 volumetric projection) |
| 44 | Area 6m anterior (Left) | Ctx_6ma_L | Ctx_6ma_L | SMA_L | somatomotor_paracentral_lobule_L | Glasser2016 (Petre2023 volumetric projection) |
| 45 | Medial Area 7A (Left) | Ctx_7Am_L | Ctx_7Am_L | anterior_BA7_L | parietal_superior_lobule_L | Glasser2016 (Petre2023 volumetric projection) |
| 46 | Lateral Area 7P (Left) | Ctx_7PL_L | Ctx_7PL_L | posterior_BA7_L | parietal_superior_lobule_L | Glasser2016 (Petre2023 volumetric projection) |
| 47 | Area 7 postcentral (Left) | Ctx_7PC_L | Ctx_7PC_L | postcentral_BA7_L | parietal_superior_lobule_L | Glasser2016 (Petre2023 volumetric projection) |
| 48 | Area Lateral IntraParietal ventral (Left) | Ctx_LIPv_L | Ctx_LIPv_L | dorsal_intraparietal_L | parietal_superior_lobule_L | Glasser2016 (Petre2023 volumetric projection) |
| 49 | Ventral IntraParietal Complex (Left) | Ctx_VIP_L | Ctx_VIP_L | dorsal_intraparietal_L | parietal_superior_lobule_L | Glasser2016 (Petre2023 volumetric projection) |
| 50 | Medial IntraParietal Area (Left) | Ctx_MIP_L | Ctx_MIP_L | MIP_L | parietal_superior_lobule_L | Glasser2016 (Petre2023 volumetric projection) |
| 51 | Area 1 (Left) | Ctx_1_L | Ctx_1_L | S1_L | somatomotor_primary_L | Glasser2016 (Petre2023 |

| | | | | | | |
|---|---|---|---|---|---|---|
| | | | | | | volumetric projection) |
| 52 | Area 2 (Left) | Ctx_2_L | Ctx_2_L | S1_L | somatomotor_primary_L | Glasser2016 (Petre2023 volumetric projection) |
| 53 | Area 3a (Left) | Ctx_3a_L | Ctx_3a_L | S1_L | somatomotor_primary_L | Glasser2016 (Petre2023 volumetric projection) |
| 54 | Dorsal area 6 (Left) | Ctx_6d_L | Ctx_6d_L | superior_premotor_L | somatomotor_premotor_L | Glasser2016 (Petre2023 volumetric projection) |
| 55 | Area 6mp (Left) | Ctx_6mp_L | Ctx_6mp_L | SMA_L | somatomotor_paracentral_lobule_L | Glasser2016 (Petre2023 volumetric projection) |
| 56 | Ventral Area 6 (Left) | Ctx_6v_L | Ctx_6v_L | inferior_premotor_L | somatomotor_premotor_L | Glasser2016 (Petre2023 volumetric projection) |
| 57 | Area Posterior 24 prime (Left) | Ctx_p24pr_L | Ctx_p24pr_L | midcingulate_L | cingulate_ACC_mPFC_L | Glasser2016 (Petre2023 volumetric projection) |
| 58 | Area 33 prime (Left) | Ctx_33pr_L | Ctx_33pr_L | midcingulate_L | cingulate_ACC_mPFC_L | Glasser2016 (Petre2023 volumetric projection) |
| **59** | Anterior 24 prime (Left) | Ctx_a24pr_L | Ctx_a24pr_L | midcingulate_L | cingulate_ACC_mPFC_L | Glasser2016 (Petre2023 volumetric projection) |

| | | | | | | |
|---|---|---|---|---|---|---|
| **60** | Area p32 prime (Left) | Ctx_p32pr_L | Ctx_p32pr_L | midcingulate_L | cingulate_ACC_mPFC_L | Glasser2016 (Petre2023 volumetric projection) |
| **61** | Area a24 (Left) | Ctx_a24_L | Ctx_a24_L | vmPFC_L | cingulate_ACC_mPFC_L | Glasser2016 (Petre2023 volumetric projection) |
| **62** | Area dorsal 32 (Left) | Ctx_d32_L | Ctx_d32_L | dmPFC_L | cingulate_ACC_mPFC_L | Glasser2016 (Petre2023 volumetric projection) |
| **63** | Area 8BM (Left) | Ctx_8BM_L | Ctx_8BM_L | dmPFC_L | cingulate_ACC_mPFC_L | Glasser2016 (Petre2023 volumetric projection) |
| **64** | Area p32 (Left) | Ctx_p32_L | Ctx_p32_L | vmPFC_L | cingulate_ACC_mPFC_L | Glasser2016 (Petre2023 volumetric projection) |
| **65** | Area 10r (Left) | Ctx_10r_L | Ctx_10r_L | BA10_L | cingulate_ventral_frontal_L | Glasser2016 (Petre2023 volumetric projection) |
| **66** | Area 47m (Left) | Ctx_47m_L | Ctx_47m_L | BA47_L | cingulate_ventral_frontal_L | Glasser2016 (Petre2023 volumetric projection) |
| **67** | Area 8Av (Left) | Ctx_8Av_L | Ctx_8Av_L | Inferior_lateral_BA8_L | cingulate_dlPFC_L | Glasser2016 (Petre2023 volumetric projection) |
| **68** | Area 8Ad (Left) | Ctx_8Ad_L | Ctx_8Ad_L | Superior_lateral_BA8_L | cingulate_dlPFC_L | Glasser2016 (Petre2023 |

| | | | | | | |
|---|---|---|---|---|---|---|
| | | | | | | volumetric projection) |
| **69** | Area 9 Middle (Left) | Ctx_9m_L | Ctx_9m_L | dmPFC_L | cingulate_ACC_mPFC_L | Glasser2016 (Petre2023 volumetric projection) |
| **70** | Area 88 Lateral (Left) | Ctx_8BL_L | Ctx_8BL_L | Superior_lateral_BA8_L | cingulate_dlPFC_L | Glasser2016 (Petre2023 volumetric projection) |
| **71** | Area 9 Posterior (Left) | Ctx_9p_L | Ctx_9p_L | BA9_L | cingulate_dlPFC_L | Glasser2016 (Petre2023 volumetric projection) |
| **72** | Area 10d (Left) | Ctx_10d_L | Ctx_10d_L | BA10_L | cingulate_ventral_frontal_L | Glasser2016 (Petre2023 volumetric projection) |
| **73** | Area 8C (Left) | Ctx_8C_L | Ctx_8C_L | Inferior_lateral_BA8_L | cingulate_dlPFC_L | Glasser2016 (Petre2023 volumetric projection) |
| **74** | Area 44 (Left) | Ctx_44_L | Ctx_44_L | BA44_L | cingulate_vlPFC_L | Glasser2016 (Petre2023 volumetric projection) |
| **75** | Area 45 (Left) | Ctx_45_L | Ctx_45_L | BA45_L | cingulate_vlPFC_L | Glasser2016 (Petre2023 volumetric projection) |
| **76** | Area 47 lateral (Left) | Ctx_47l_L | Ctx_47l_L | BA47_L | cingulate_ventral_frontal_L | Glasser2016 (Petre2023 volumetric projection) |

| | | | | | | |
|---|---|---|---|---|---|---|
| **77** | Area anterior 47r (Left) | Ctx_a47r_L | Ctx_a47r_L | BA47_L | cingulate_ventral_frontal_L | Glasser2016 (Petre2023 volumetric projection) |
| **78** | Rostral Area 6 (Left) | Ctx_6r_L | Ctx_6r_L | inferior_premotor_L | somatomotor_premotor_L | Glasser2016 (Petre2023 volumetric projection) |
| **79** | Area IF Ja (Left) | Ctx_IFJa_L | Ctx_IFJa_L | IFJ_L | cingulate_vlPFC_L | Glasser2016 (Petre2023 volumetric projection) |
| **80** | Area IF Jp (Left) | Ctx_IFJp_L | Ctx_IFJp_L | IFJ_L | cingulate_vlPFC_L | Glasser2016 (Petre2023 volumetric projection) |
| **81** | Area IF Sp (Left) | Ctx_IFSp_L | Ctx_IFSp_L | IFS_L | cingulate_vlPFC_L | Glasser2016 (Petre2023 volumetric projection) |
| **82** | Area IFSa (Left) | Ctx_IFSa_L | Ctx_IFSa_L | IFS_L | cingulate_vlPFC_L | Glasser2016 (Petre2023 volumetric projection) |
| **83** | Area posterior 9-46v (Left) | Ctx_p9_46v_L | Ctx_p9_46v_L | BA46_L | cingulate_dlPFC_L | Glasser2016 (Petre2023 volumetric projection) |
| **84** | Area 46 (Left) | Ctx_46_L | Ctx_46_L | BA46_L | cingulate_dlPFC_L | Glasser2016 (Petre2023 volumetric projection) |
| **85** | Area anterior 9-46v (Left) | Ctx_a9_46v_L | Ctx_a9_46v_L | BA46_L | cingulate_dlPFC_L | Glasser2016 (Petre2023 |

| | | | | | | |
|---|---|---|---|---|---|---|
| | | | | | | volumetric projection) |
| **86** | Area 9-46d (Left) | Ctx_9_46d_L | Ctx_9_46d_L | BA46_L | cingulate_dlPFC_L | Glasser2016 (Petre2023 volumetric projection) |
| **87** | Area 9 anterior (Left) | Ctx_9a_L | Ctx_9a_L | BA9_L | cingulate_dlPFC_L | Glasser2016 (Petre2023 volumetric projection) |
| **88** | Area 10v (Left) | Ctx_10v_L | Ctx_10v_L | BA10_L | cingulate_ventral_frontal_L | Glasser2016 (Petre2023 volumetric projection) |
| **89** | Area anterior 10p (Left) | Ctx_a10p_L | Ctx_a10p_L | BA10_L | cingulate_ventral_frontal_L | Glasser2016 (Petre2023 volumetric projection) |
| **90** | Polar 10p (Left) | Ctx_10pp_L | Ctx_10pp_L | BA10_L | cingulate_ventral_frontal_L | Glasser2016 (Petre2023 volumetric projection) |
| **91** | Area 11l (Left) | Ctx_11l_L | Ctx_11l_L | BA11_L | cingulate_ventral_frontal_L | Glasser2016 (Petre2023 volumetric projection) |
| **92** | Area 13l (Left) | Ctx_13l_L | Ctx_13l_L | BA13_L | cingulate_ventral_frontal_L | Glasser2016 (Petre2023 volumetric projection) |
| **93** | Orbital Frontal Complex (Left) | Ctx_OFC_L | Ctx_OFC_L | OFC_L | cingulate_ventral_frontal_L | Glasser2016 (Petre2023 volumetric projection) |

| | | | | | | |
|---|---|---|---|---|---|---|
| **94** | Area 47s (Left) | Ctx_47s_L | Ctx_47s_L | BA47_L | cingulate_ventral_frontal_L | Glasser2016 (Petre2023 volumetric projection) |
| **95** | Area Lateral IntraParietal dorsal (Left) | Ctx_LIPd_L | Ctx_LIPd_L | ventral_intraparietal_L | parietal_superior_lobule_L | Glasser2016 (Petre2023 volumetric projection) |
| **96** | Area 6 anterior (Left) | Ctx_6a_L | Ctx_6a_L | superior_premotor_L | somatomotor_premotor_L | Glasser2016 (Petre2023 volumetric projection) |
| **97** | Inferior 6-8 Transitional Area (Left) | Ctx_i6_8_L | Ctx_i6_8_L | Inferior_lateral_BA8_L | cingulate_dlPFC_L | Glasser2016 (Petre2023 volumetric projection) |
| **98** | Superior 6-8 Transitional Area (Left) | Ctx_s6_8_L | Ctx_s6_8_L | Superior_lateral_BA8_L | cingulate_dlPFC_L | Glasser2016 (Petre2023 volumetric projection) |
| **99** | Putative primary gustatory cortex (Left) | Ctx_43_L | Ctx_43_L | FOP1-BA43_L | somatomotor_operculum_L | Glasser2016 (Petre2023 volumetric projection) |
| **100** | Area OP4/PV (Left) | Ctx_OP4_L | Ctx_OP4_L | OP4_L | somatomotor_operculum_L | Glasser2016 (Petre2023 volumetric projection) |
| **101** | Area OP1/SII (Left) | Ctx_OP1_L | Ctx_OP1_L | SII+_L | somatomotor_operculum_L | Glasser2016 (Petre2023 volumetric projection) |
| **102** | Area OP2-3/VS (Left) | Ctx_OP2_3_L | Ctx_OP2_3_L | SII+_L | somatomotor_operculum_L | Glasser2016 (Petre2023 |

| | | | | | | |
|---|---|---|---|---|---|---|
| | | | | | | volumetric projection) |
| **103** | Area 52 (Left) | Ctx_52_L | Ctx_52_L | posterior_granular_insula_L | insula_posterior_L | Glasser2016 (Petre2023 volumetric projection) |
| **104** | RetroInsular Cortex (Left) | Ctx_RI_L | Ctx_RI_L | auditory+_L | auditory_early_L | Glasser2016 (Petre2023 volumetric projection) |
| **105** | Area Pfcm (Left) | Ctx_PFcm_L | Ctx_PFcm_L | SII+_L | somatomotor_operculum_L | Glasser2016 (Petre2023 volumetric projection) |
| **106** | insula_posteriorr Area 2 (Left) | Ctx_PoI2_L | Ctx_PoI2_L | inferior_dysgranular_insula_L | insula_posterior_L | Glasser2016 (Petre2023 volumetric projection) |
| **107** | Area TA2 (Left) | Ctx_TA2_L | Ctx_TA2_L | auditory_transition_area_L | auditory_association_cortex_L | Glasser2016 (Petre2023 volumetric projection) |
| **108** | Frontal Opercular Area 4 (Left) | Ctx_FOP4_L | Ctx_FOP4_L | anterior_operculum_L | insula_operculum_L | Glasser2016 (Petre2023 volumetric projection) |
| **109** | Middle Insular Area (Left) | Ctx_MI_L | Ctx_MI_L | anterior_agranular_insula_L | insula_anterior_L | Glasser2016 (Petre2023 volumetric projection) |
| **110** | Piriform olfactory cortex (Left) | Ctx_Pir_L | Ctx_Pir_L | PIR_L | insula_anterior_L | Glasser2016 (Petre2023 volumetric projection) |

| | | | | | | |
|---|---|---|---|---|---|---|
| **111** | Anterior Ventral Insular Area (Left) | Ctx_AVI_L | Ctx_AVI_L | anterior_agranular_insula_L | insula_anterior_L | Glasser2016 (Petre2023 volumetric projection) |
| **112** | Anterior Agranular Insular Complex (Left) | Ctx_AAIC_L | Ctx_AAIC_L | anterior_agranular_insula_L | insula_anterior_L | Glasser2016 (Petre2023 volumetric projection) |
| **113** | Frontal Opercular Area 1 (Left) | Ctx_FOP1_L | Ctx_FOP1_L | FOP1-BA43_L | somatomotor_operculum_L | Glasser2016 (Petre2023 volumetric projection) |
| **114** | Frontal Opercular Area 3 (Left) | Ctx_FOP3_L | Ctx_FOP3_L | posterior_operculum_L | insula_operculum_L | Glasser2016 (Petre2023 volumetric projection) |
| **115** | Frontal Opercular Area 2 (Left) | Ctx_FOP2_L | Ctx_FOP2_L | posterior_operculum_L | insula_operculum_L | Glasser2016 (Petre2023 volumetric projection) |
| **116** | Parietal area F part t (Left) | Ctx_PFt_L | Ctx_PFt_L | anterior_IPL_L | parietal_inferior_lobule_L | Glasser2016 (Petre2023 volumetric projection) |
| **117** | Anterior IntraParietal Area (Left) | Ctx_AIP_L | Ctx_AIP_L | ventral_intraparietal_L | parietal_superior_lobule_L | Glasser2016 (Petre2023 volumetric projection) |
| **118** | Entorhinal Cortex (Left) | Ctx_EC_L | Ctx_EC_L | hippocampal_formation_L | temporal_medial_L | Glasser2016 (Petre2023 volumetric projection) |
| **119** | PreSubiculum (Left) | Ctx_PreS_L | Ctx_PreS_L | hippocampal_formation_L | temporal_medial_L | Glasser2016 (Petre2023 |

| | | | | | | |
|---|---|---|---|---|---|---|
| | | | | | | volumetric projection) |
| **120** | Prostriate Area (Left) | Ctx_ProS_L | Ctx_ProS_L | parietal_occipital_sulcus_L | cingulate_posterior_L | Glasser2016 (Petre2023 volumetric projection) |
| **121** | Perirhinal Ectorhinal Cortex (Left) | Ctx_PeEc_L | Ctx_PeEc_L | PeEc_L | temporal_medial_L | Glasser2016 (Petre2023 volumetric projection) |
| **122** | Area STGa (Left) | Ctx_STGa_L | Ctx_STGa_L | auditory_transition_area_L | auditory_association_cortex_L | Glasser2016 (Petre2023 volumetric projection) |
| **123** | ParaBelt Complex (Left) | Ctx_PBelt_L | Ctx_PBelt_L | auditory+_L | auditory_early_L | Glasser2016 (Petre2023 volumetric projection) |
| **124** | Auditory 5 Complex (Left) | Ctx_A5_L | Ctx_A5_L | auditory_transition_area_L | auditory_association_cortex_L | Glasser2016 (Petre2023 volumetric projection) |
| **125** | ParaHippocampal Area 1 (Left) | Ctx_PHA1_L | Ctx_PHA1_L | PHA_L | temporal_medial_L | Glasser2016 (Petre2023 volumetric projection) |
| **126** | ParaHippocampal Area 3 (Left) | Ctx_PHA3_L | Ctx_PHA3_L | PHA_L | temporal_medial_L | Glasser2016 (Petre2023 volumetric projection) |
| **127** | Area STSd anterior (Left) | Ctx_STSda_L | Ctx_STSda_L | anterior_AAC_L | auditory_association_cortex_L | Glasser2016 (Petre2023 volumetric projection) |

| | | | | | | |
|---|---|---|---|---|---|---|
| **128** | Area STSd posterior (Left) | Ctx_STSdp_L | Ctx_STSdp_L | posterior_AAC_L | auditory_association_cortex_L | Glasser2016 (Petre2023 volumetric projection) |
| **129** | Area STSv posterior (Left) | Ctx_STSvp_L | Ctx_STSvp_L | posterior_AAC_L | auditory_association_cortex_L | Glasser2016 (Petre2023 volumetric projection) |
| **130** | Area TG dorsal (Left) | Ctx_TGd_L | Ctx_TGd_L | temporal_pole_L | temporal_lateral_L | Glasser2016 (Petre2023 volumetric projection) |
| **131** | Area TE1 anterior (Left) | Ctx_TE1a_L | Ctx_TE1a_L | temporal_pole_L | temporal_lateral_L | Glasser2016 (Petre2023 volumetric projection) |
| **132** | Area TE1 posterior (Left) | Ctx_TE1p_L | Ctx_TE1p_L | middle_temporal_gyrus_L | temporal_lateral_L | Glasser2016 (Petre2023 volumetric projection) |
| **133** | Area TE2 anterior (Left) | Ctx_TE2a_L | Ctx_TE2a_L | inferior_temporal_sulcus_L | temporal_lateral_L | Glasser2016 (Petre2023 volumetric projection) |
| **134** | Area TF (Left) | Ctx_TF_L | Ctx_TF_L | inferior_temporal_sulcus_L | temporal_lateral_L | Glasser2016 (Petre2023 volumetric projection) |
| **135** | Area TE2 posterior (Left) | Ctx_TE2p_L | Ctx_TE2p_L | inferior_temporal_sulcus_L | temporal_lateral_L | Glasser2016 (Petre2023 volumetric projection) |
| **136** | Area PHT (Left) | Ctx_PHT_L | Ctx_PHT_L | middle_temporal_gyrus_L | temporal_lateral_L | Glasser2016 (Petre2023 |

| | | | | | | |
|---|---|---|---|---|---|---|
| | | | | | | volumetric projection) |
| **137** | Area PH (Left) | Ctx_PH_L | Ctx_PH_L | PH_L | visual_MT+_L | Glasser2016 (Petre2023 volumetric projection) |
| **138** | Area TemporoParietoOccipital Junction 1 (Left) | Ctx_TPOJ1_L | Ctx_TPOJ1_L | TPOJ_L | parietal_TPOJ_L | Glasser2016 (Petre2023 volumetric projection) |
| **139** | Area TemporoParietoOccipital Junction 2 (Left) | Ctx_TPOJ2_L | Ctx_TPOJ2_L | TPOJ_L | parietal_TPOJ_L | Glasser2016 (Petre2023 volumetric projection) |
| **140** | Area TemporoParietoOccipital Junction 3 (Left) | Ctx_TPOJ3_L | Ctx_TPOJ3_L | TPOJ_L | parietal_TPOJ_L | Glasser2016 (Petre2023 volumetric projection) |
| **141** | Dorsal Transitional Visual Area (Left) | Ctx_DVT_L | Ctx_DVT_L | parietal_occipital_sulcus_L | cingulate_posterior_L | Glasser2016 (Petre2023 volumetric projection) |
| **142** | Parietal area G posterior (Left) | Ctx_PGp_L | Ctx_PGp_L | posterior_IPL_L | parietal_inferior_lobule_L | Glasser2016 (Petre2023 volumetric projection) |
| **143** | Area IntraParietal 2 (Left) | Ctx_IP2_L | Ctx_IP2_L | superior_IPL_L | parietal_inferior_lobule_L | Glasser2016 (Petre2023 volumetric projection) |
| **144** | Area IntraParietal 1 (Left) | Ctx_IP1_L | Ctx_IP1_L | superior_IPL_L | parietal_inferior_lobule_L | Glasser2016 (Petre2023 volumetric projection) |

| | | | | | | |
|---|---|---|---|---|---|---|
| **145** | Area IntraParietal 0 (Left) | Ctx_IP0_L | Ctx_IP0_L | posterior_IPL_L | parietal_inferior_lobule_L | Glasser2016 (Petre2023 volumetric projection) |
| **146** | Parietal area F operculum (Left) | Ctx_PFop_L | Ctx_PFop_L | anterior_IPL_L | parietal_inferior_lobule_L | Glasser2016 (Petre2023 volumetric projection) |
| **147** | Parietal area F (Left) | Ctx_PF_L | Ctx_PF_L | anterior_IPL_L | parietal_inferior_lobule_L | Glasser2016 (Petre2023 volumetric projection) |
| **148** | Parietal area F part m (Left) | Ctx_PFm_L | Ctx_PFm_L | middle_IPL_L | parietal_inferior_lobule_L | Glasser2016 (Petre2023 volumetric projection) |
| **149** | Parietal area G inferior (Left) | Ctx_PGi_L | Ctx_PGi_L | inferior_IPL_L | parietal_inferior_lobule_L | Glasser2016 (Petre2023 volumetric projection) |
| **150** | Parietal area G superior (Left) | Ctx_PGs_L | Ctx_PGs_L | inferior_IPL_L | parietal_inferior_lobule_L | Glasser2016 (Petre2023 volumetric projection) |
| **151** | Area V6A (Left) | Ctx_V6A_L | Ctx_V6A_L | V6_L | visual_dorsal_L | Glasser2016 (Petre2023 volumetric projection) |
| **152** | VentroMedial Visual Area 1 (Left) | Ctx_VMV1_L | Ctx_VMV1_L | VMV_L | visual_ventral_L | Glasser2016 (Petre2023 volumetric projection) |
| **153** | VentroMedial Visual Area 3 (Left) | Ctx_VMV3_L | Ctx_VMV3_L | VMV_L | visual_ventral_L | Glasser2016 (Petre2023 |

| | | | | | | |
|---|---|---|---|---|---|---|
| | | | | | | volumetric projection) |
| **154** | ParaHippocampal Area 2 (Left) | Ctx_PHA2_L | Ctx_PHA2_L | PHA_L | temporal_medial_L | Glasser2016 (Petre2023 volumetric projection) |
| **155** | Area V4t (Left) | Ctx_V4t_L | Ctx_V4t_L | vMT+_L | visual_MT+_L | Glasser2016 (Petre2023 volumetric projection) |
| **156** | Fundus of the superior temporal area (Left) | Ctx_FST_L | Ctx_FST_L | vMT+_L | visual_MT+_L | Glasser2016 (Petre2023 volumetric projection) |
| **157** | Area v3CD (Left) | Ctx_V3CD_L | Ctx_V3CD_L | supplementary_V3_L | visual_dorsal_L | Glasser2016 (Petre2023 volumetric projection) |
| **158** | Area Lateral Occipital 3 (Left) | Ctx_LO3_L | Ctx_LO3_L | LO_L | visual_MT+_L | Glasser2016 (Petre2023 volumetric projection) |
| **159** | VentroMedial Visual Area 2 (Left) | Ctx_VMV2_L | Ctx_VMV2_L | VMV_L | visual_ventral_L | Glasser2016 (Petre2023 volumetric projection) |
| **160** | Area 31pd (Left) | Ctx_31pd_L | Ctx_31pd_L | precuneus_L | cingulate_posterior_L | Glasser2016 (Petre2023 volumetric projection) |
| **161** | Area 31a (Left) | Ctx_31a_L | Ctx_31a_L | anterior_precuneus_L | cingulate_posterior_L | Glasser2016 (Petre2023 volumetric projection) |

| | | | | | | |
|---|---|---|---|---|---|---|
| **162** | Ventral Visual Complex (Left) | Ctx_VVC_L | Ctx_VVC_L | VVC_L | visual_ventral_L | Glasser2016 (Petre2023 volumetric projection) |
| **163** | Area 25 (Left) | Ctx_25_L | Ctx_25_L | infralimbic_L | cingulate_ACC_mPFC_L | Glasser2016 (Petre2023 volumetric projection) |
| **164** | Area s32 (Left) | Ctx_s32_L | Ctx_s32_L | vmPFC_L | cingulate_ACC_mPFC_L | Glasser2016 (Petre2023 volumetric projection) |
| **165** | posterior OFC Complex (Left) | Ctx_pOFC_L | Ctx_pOFC_L | OFC_L | cingulate_ventral_frontal_L | Glasser2016 (Petre2023 volumetric projection) |
| **166** | Area insula_posteriorr 1 (Left) | Ctx_PoI1_L | Ctx_PoI1_L | inferior_dysgranular_insula_L | insula_posterior_L | Glasser2016 (Petre2023 volumetric projection) |
| **167** | Insular Granular Complex (Left) | Ctx_Ig_L | Ctx_Ig_L | posterior_granular_insula_L | insula_posterior_L | Glasser2016 (Petre2023 volumetric projection) |
| **168** | Area Frontal Opercular 5 (Left) | Ctx_FOP5_L | Ctx_FOP5_L | anterior_operculum_L | insula_operculum_L | Glasser2016 (Petre2023 volumetric projection) |
| **169** | Area posterior 10p (Left) | Ctx_p10p_L | Ctx_p10p_L | BA10_L | cingulate_ventral_frontal_L | Glasser2016 (Petre2023 volumetric projection) |
| **170** | Area posterior 47r (Left) | Ctx_p47r_L | Ctx_p47r_L | BA47_L | cingulate_ventral_frontal_L | Glasser2016 (Petre2023 |

| | | | | | | |
|---|---|---|---|---|---|---|
| | | | | | | volumetric projection) |
| **171** | Area TG Ventral (Left) | Ctx_TGv_L | Ctx_TGv_L | temporal_pole_L | temporal_lateral_L | Glasser2016 (Petre2023 volumetric projection) |
| **172** | Medial Belt Complex (Left) | Ctx_MBelt_L | Ctx_MBelt_L | auditory+_L | auditory_early_L | Glasser2016 (Petre2023 volumetric projection) |
| **173** | Lateral Belt Complex (Left) | Ctx_LBelt_L | Ctx_LBelt_L | auditory+_L | auditory_early_L | Glasser2016 (Petre2023 volumetric projection) |
| **174** | Auditory 4 Complex (Left) | Ctx_A4_L | Ctx_A4_L | auditory_transition_area_L | auditory_association_cortex_L | Glasser2016 (Petre2023 volumetric projection) |
| **175** | Area STSv anterior (Left) | Ctx_STSva_L | Ctx_STSva_L | anterior_AAC_L | auditory_association_cortex_L | Glasser2016 (Petre2023 volumetric projection) |
| **176** | Area TE1 Middle (Left) | Ctx_TE1m_L | Ctx_TE1m_L | middle_temporal_gyrus_L | temporal_lateral_L | Glasser2016 (Petre2023 volumetric projection) |
| **177** | Para-Insular Area (Left) | Ctx_PI_L | Ctx_PI_L | inferior_dysgranular_insula_L | insula_posterior_L | Glasser2016 (Petre2023 volumetric projection) |
| **178** | Area anterior 32 prime (Left) | Ctx_a32pr_L | Ctx_a32pr_L | midcingulate_L | cingulate_ACC_mPFC_L | Glasser2016 (Petre2023 volumetric projection) |

| | | | | | | |
|---|---|---|---|---|---|---|
| **179** | Area posterior 24 (Left) | Ctx_p24_L | Ctx_p24_L | midcingulate_L | cingulate_ACC_mPFC_L | Glasser2016 (Petre2023 volumetric projection) |
| **180** | Primary visual cortex (Right) | Ctx_V1_R | Ctx_V1_R | striate_R | visual_early_R | Glasser2016 (Petre2023 volumetric projection) |
| **181** | Medial superior temporal area (Right) | Ctx_MST_R | Ctx_MST_R | dMT+_R | visual_MT+_R | Glasser2016 (Petre2023 volumetric projection) |
| **182** | sixth visual area (Right) | Ctx_V6_R | Ctx_V6_R | V6_R | visual_dorsal_R | Glasser2016 (Petre2023 volumetric projection) |
| **183** | second visual area (Right) | Ctx_V2_R | Ctx_V2_R | extrastriate_R | visual_early_R | Glasser2016 (Petre2023 volumetric projection) |
| **184** | third visual area (Right) | Ctx_V3_R | Ctx_V3_R | extrastriate_R | visual_early_R | Glasser2016 (Petre2023 volumetric projection) |
| **185** | fourth visual area (Right) | Ctx_V4_R | Ctx_V4_R | extrastriate_R | visual_early_R | Glasser2016 (Petre2023 volumetric projection) |
| **186** | eighth visual area (Right) | Ctx_V8_R | Ctx_V8_R | VVC_R | visual_ventral_R | Glasser2016 (Petre2023 volumetric projection) |
| **187** | primary motor cortex (Right) | Ctx_4_R | Ctx_4_R | M1_R | somatomotor_primary_R | Glasser2016 (Petre2023 |

| | | | | | | |
|---|---|---|---|---|---|---|
| | | | | | | volumetric projection) |
| **188** | primary sensory cortex (Right) | Ctx_3b_R | Ctx_3b_R | S1_R | somatomotor_primary_R | Glasser2016 (Petre2023 volumetric projection) |
| **189** | frontal eye fields (Right) | Ctx_FEF_R | Ctx_FEF_R | superior_premotor_R | somatomotor_premotor_R | Glasser2016 (Petre2023 volumetric projection) |
| **190** | premotor eye field (Right) | Ctx_PEF_R | Ctx_PEF_R | inferior_premotor_R | somatomotor_premotor_R | Glasser2016 (Petre2023 volumetric projection) |
| **191** | area 55b (Right) | Ctx_55b_R | Ctx_55b_R | 55b_R | somatomotor_premotor_R | Glasser2016 (Petre2023 volumetric projection) |
| **192** | area V3A (Right) | Ctx_V3A_R | Ctx_V3A_R | supplementary_V3_R | visual_dorsal_R | Glasser2016 (Petre2023 volumetric projection) |
| **193** | RetroSplenial Complex (Right) | Ctx_RSC_R | Ctx_RSC_R | RSC_R | cingulate_posterior_R | Glasser2016 (Petre2023 volumetric projection) |
| **194** | Parieto-Occipital Sulcus Area 2 (Right) | Ctx_POS2_R | Ctx_POS2_R | parietal_occipital_sulcus_R | cingulate_posterior_R | Glasser2016 (Petre2023 volumetric projection) |
| **195** | Seventh Visual Area (Right) | Ctx_V7_R | Ctx_V7_R | V7_R | visual_dorsal_R | Glasser2016 (Petre2023 volumetric projection) |

| | | | | | | |
|---|---|---|---|---|---|---|
| **196** | IntraParietal Sulcus Area 1 (Right) | Ctx_IPS1_R | Ctx_IPS1_R | ISP1_R | visual_dorsal_R | Glasser2016 (Petre2023 volumetric projection) |
| **197** | Fusiform Face Complex (Right) | Ctx_FFC_R | Ctx_FFC_R | FFC_R | visual_ventral_R | Glasser2016 (Petre2023 volumetric projection) |
| **198** | Area V3B (Right) | Ctx_V3B_R | Ctx_V3B_R | supplementary_V3_R | visual_dorsal_R | Glasser2016 (Petre2023 volumetric projection) |
| **199** | Area Lateral Occipital 1 (Right) | Ctx_LO1_R | Ctx_LO1_R | LO_R | visual_MT+_R | Glasser2016 (Petre2023 volumetric projection) |
| **200** | Area Lateral Occipital 2 (Right) | Ctx_LO2_R | Ctx_LO2_R | LO_R | visual_MT+_R | Glasser2016 (Petre2023 volumetric projection) |
| **201** | Posterior InferoTemporal (Right) | Ctx_PIT_R | Ctx_PIT_R | FFC_R | visual_ventral_R | Glasser2016 (Petre2023 volumetric projection) |
| **202** | Middle Temporal Area (Right) | Ctx_MT_R | Ctx_MT_R | dMT+_R | visual_MT+_R | Glasser2016 (Petre2023 volumetric projection) |
| **203** | Primary Auditory Cortex (Right) | Ctx_A1_R | Ctx_A1_R | A1_R | auditory_early_R | Glasser2016 (Petre2023 volumetric projection) |
| **204** | PeriSylvian Language Area (Right) | Ctx_PSL_R | Ctx_PSL_R | PSL_R | parietal_TPOJ_R | Glasser2016 (Petre2023 |

| | | | | | | |
|---|---|---|---|---|---|---|
| | | | | | | volumetric projection) |
| **205** | Superior Frontal Language Area (Right) | Ctx_SFL_R | Ctx_SFL_R | Superior_lateral_BA8_R | cingulate_dlPFC_R | Glasser2016 (Petre2023 volumetric projection) |
| **206** | PreCuneus Visual Area (Right) | Ctx_PCV_R | Ctx_PCV_R | PCV_R | cingulate_posterior_R | Glasser2016 (Petre2023 volumetric projection) |
| **207** | Superior Temporal Visual Area (Right) | Ctx_STV_R | Ctx_STV_R | STV_R | parietal_TPOJ_R | Glasser2016 (Petre2023 volumetric projection) |
| **208** | Medial Area 7P (Right) | Ctx_7Pm_R | Ctx_7Pm_R | posterior_BA7_R | parietal_superior_lobule_R | Glasser2016 (Petre2023 volumetric projection) |
| **209** | Area 7m (Right) | Ctx_7m_R | Ctx_7m_R | precuneus_R | cingulate_posterior_R | Glasser2016 (Petre2023 volumetric projection) |
| **210** | Parieto-Occipital Sulcus Area 1 (Right) | Ctx_POS1_R | Ctx_POS1_R | parietal_occipital_sulcus_R | cingulate_posterior_R | Glasser2016 (Petre2023 volumetric projection) |
| **211** | Area 23d (Right) | Ctx_23d_R | Ctx_23d_R | anterior_precuneus_R | cingulate_posterior_R | Glasser2016 (Petre2023 volumetric projection) |
| **212** | Area ventral 23 a+b (Right) | Ctx_v23ab_R | Ctx_v23ab_R | precuneus_R | cingulate_posterior_R | Glasser2016 (Petre2023 volumetric projection) |

| | | | | | | |
|---|---|---|---|---|---|---|
| **213** | Area dorsal 23 a+ (Right) | Ctx_d23ab_R | Ctx_d23ab_R | precuneus_R | cingulate_posterior_R | Glasser2016 (Petre2023 volumetric projection) |
| **214** | Area 31p ventral (Right) | Ctx_31pv_R | Ctx_31pv_R | precuneus_R | cingulate_posterior_R | Glasser2016 (Petre2023 volumetric projection) |
| **215** | Area 5m (Right) | Ctx_5m_R | Ctx_5m_R | BA5_R | somatomotor_paracentral_lobule_R | Glasser2016 (Petre2023 volumetric projection) |
| **216** | Area 5m ventral (Right) | Ctx_5mv_R | Ctx_5mv_R | BA5_R | somatomotor_paracentral_lobule_R | Glasser2016 (Petre2023 volumetric projection) |
| **217** | Area 23c (Right) | Ctx_23c_R | Ctx_23c_R | anterior_precuneus_R | cingulate_posterior_R | Glasser2016 (Petre2023 volumetric projection) |
| **218** | Area 5L (Right) | Ctx_5L_R | Ctx_5L_R | BA5_R | somatomotor_paracentral_lobule_R | Glasser2016 (Petre2023 volumetric projection) |
| **219** | Dorsal Area 24d (Right) | Ctx_24dd_R | Ctx_24dd_R | cingulate_motor_area_R | somatomotor_paracentral_lobule_R | Glasser2016 (Petre2023 volumetric projection) |
| **220** | Ventral Area 24d (Right) | Ctx_24dv_R | Ctx_24dv_R | cingulate_motor_area_R | somatomotor_paracentral_lobule_R | Glasser2016 (Petre2023 volumetric projection) |
| **221** | Lateral Area 7A (Right) | Ctx_7AL_R | Ctx_7AL_R | anterior_BA7_R | parietal_superior_lobule_R | Glasser2016 (Petre2023 |

| | | | | | | |
|---|---|---|---|---|---|---|
| | | | | | | volumetric projection) |
| **222** | Supplementary and Cingulate Eye Field (Right) | Ctx_SCEF_R | Ctx_SCEF_R | SMA_R | somatomotor_paracentral_lobule_R | Glasser2016 (Petre2023 volumetric projection) |
| **223** | Area 6m anterior (Right) | Ctx_6ma_R | Ctx_6ma_R | SMA_R | somatomotor_paracentral_lobule_R | Glasser2016 (Petre2023 volumetric projection) |
| **224** | Medial Area 7A (Right) | Ctx_7Am_R | Ctx_7Am_R | anterior_BA7_R | parietal_superior_lobule_R | Glasser2016 (Petre2023 volumetric projection) |
| **225** | Lateral Area 7P (Right) | Ctx_7PL_R | Ctx_7PL_R | posterior_BA7_R | parietal_superior_lobule_R | Glasser2016 (Petre2023 volumetric projection) |
| **226** | Area 7 postcentral (Right) | Ctx_7PC_R | Ctx_7PC_R | postcentral_BA7_R | parietal_superior_lobule_R | Glasser2016 (Petre2023 volumetric projection) |
| **227** | Area Lateral IntraParietal ventral (Right) | Ctx_LIPv_R | Ctx_LIPv_R | dorsal_intraparietal_R | parietal_superior_lobule_R | Glasser2016 (Petre2023 volumetric projection) |
| **228** | Ventral IntraParietal Complex (Right) | Ctx_VIP_R | Ctx_VIP_R | dorsal_intraparietal_R | parietal_superior_lobule_R | Glasser2016 (Petre2023 volumetric projection) |
| **229** | Medial IntraParietal Area (Right) | Ctx_MIP_R | Ctx_MIP_R | MIP_R | parietal_superior_lobule_R | Glasser2016 (Petre2023 volumetric projection) |

| | | | | | | |
|---|---|---|---|---|---|---|
| **230** | Area 1 (Right) | Ctx_1_R | Ctx_1_R | S1_R | somatomotor_primary_R | Glasser2016 (Petre2023 volumetric projection) |
| **231** | Area 2 (Right) | Ctx_2_R | Ctx_2_R | S1_R | somatomotor_primary_R | Glasser2016 (Petre2023 volumetric projection) |
| **232** | Area 3a (Right) | Ctx_3a_R | Ctx_3a_R | S1_R | somatomotor_primary_R | Glasser2016 (Petre2023 volumetric projection) |
| **233** | Dorsal area 6 (Right) | Ctx_6d_R | Ctx_6d_R | superior_premotor_R | somatomotor_premotor_R | Glasser2016 (Petre2023 volumetric projection) |
| **234** | Area 6mp (Right) | Ctx_6mp_R | Ctx_6mp_R | SMA_R | somatomotor_paracentral_lobule_R | Glasser2016 (Petre2023 volumetric projection) |
| **235** | Ventral Area 6 (Right) | Ctx_6v_R | Ctx_6v_R | inferior_premotor_R | somatomotor_premotor_R | Glasser2016 (Petre2023 volumetric projection) |
| **236** | Area Posterior 24 prime (Right) | Ctx_p24pr_R | Ctx_p24pr_R | midcingulate_R | cingulate_ACC_mPFC_R | Glasser2016 (Petre2023 volumetric projection) |
| **237** | Area 33 prime (Right) | Ctx_33pr_R | Ctx_33pr_R | midcingulate_R | cingulate_ACC_mPFC_R | Glasser2016 (Petre2023 volumetric projection) |
| **238** | Anterior 24 prime (Right) | Ctx_a24pr_R | Ctx_a24pr_R | midcingulate_R | cingulate_ACC_mPFC_R | Glasser2016 (Petre2023 |

| | | | | | | |
|---|---|---|---|---|---|---|
| | | | | | | volumetric projection) |
| **239** | Area p32 prime (Right) | Ctx_p32pr_R | Ctx_p32pr_R | midcingulate_R | cingulate_ACC_mPFC_R | Glasser2016 (Petre2023 volumetric projection) |
| **240** | Area a24 (Right) | Ctx_a24_R | Ctx_a24_R | vmPFC_R | cingulate_ACC_mPFC_R | Glasser2016 (Petre2023 volumetric projection) |
| **241** | Area dorsal 32 (Right) | Ctx_d32_R | Ctx_d32_R | dmPFC_R | cingulate_ACC_mPFC_R | Glasser2016 (Petre2023 volumetric projection) |
| **242** | Area 8BM (Right) | Ctx_8BM_R | Ctx_8BM_R | dmPFC_R | cingulate_ACC_mPFC_R | Glasser2016 (Petre2023 volumetric projection) |
| **243** | Area p32 (Right) | Ctx_p32_R | Ctx_p32_R | vmPFC_R | cingulate_ACC_mPFC_R | Glasser2016 (Petre2023 volumetric projection) |
| **244** | Area 10r (Right) | Ctx_10r_R | Ctx_10r_R | BA10_R | cingulate_ventral_frontal_R | Glasser2016 (Petre2023 volumetric projection) |
| **245** | Area 47m (Right) | Ctx_47m_R | Ctx_47m_R | BA47_R | cingulate_ventral_frontal_R | Glasser2016 (Petre2023 volumetric projection) |
| **246** | Area 8Av (Right) | Ctx_8Av_R | Ctx_8Av_R | Inferior_lateral_BA8_R | cingulate_dlPFC_R | Glasser2016 (Petre2023 volumetric projection) |

| | | | | | | |
|---|---|---|---|---|---|---|
| **247** | Area 8Ad (Right) | Ctx_8Ad_R | Ctx_8Ad_R | Superior_lateral_BA8_R | cingulate_dlPFC_R | Glasser2016 (Petre2023 volumetric projection) |
| **248** | Area 9 Middle (Right) | Ctx_9m_R | Ctx_9m_R | dmPFC_R | cingulate_ACC_mPFC_R | Glasser2016 (Petre2023 volumetric projection) |
| **249** | Area 88 Lateral (Right) | Ctx_8BL_R | Ctx_8BL_R | Superior_lateral_BA8_R | cingulate_dlPFC_R | Glasser2016 (Petre2023 volumetric projection) |
| **250** | Area 9 Posterior (Right) | Ctx_9p_R | Ctx_9p_R | BA9_R | cingulate_dlPFC_R | Glasser2016 (Petre2023 volumetric projection) |
| **251** | Area 10d (Right) | Ctx_10d_R | Ctx_10d_R | BA10_R | cingulate_ventral_frontal_R | Glasser2016 (Petre2023 volumetric projection) |
| **252** | Area 8C (Right) | Ctx_8C_R | Ctx_8C_R | Inferior_lateral_BA8_R | cingulate_dlPFC_R | Glasser2016 (Petre2023 volumetric projection) |
| **253** | Area 44 (Right) | Ctx_44_R | Ctx_44_R | BA44_R | cingulate_vlPFC_R | Glasser2016 (Petre2023 volumetric projection) |
| **254** | Area 45 (Right) | Ctx_45_R | Ctx_45_R | BA45_R | cingulate_vlPFC_R | Glasser2016 (Petre2023 volumetric projection) |
| **255** | Area 47 lateral (Right) | Ctx_47l_R | Ctx_47l_R | BA47_R | cingulate_ventral_frontal_R | Glasser2016 (Petre2023 |

| | | | | | | |
|---|---|---|---|---|---|---|
| | | | | | | volumetric projection) |
| **256** | Area anterior 47r (Right) | Ctx_a47r_R | Ctx_a47r_R | BA47_R | cingulate_ventral_frontal_R | Glasser2016 (Petre2023 volumetric projection) |
| **257** | Rostral Area 6 (Right) | Ctx_6r_R | Ctx_6r_R | inferior_premotor_R | somatomotor_premotor_R | Glasser2016 (Petre2023 volumetric projection) |
| **258** | Area IF Ja (Right) | Ctx_IFJa_R | Ctx_IFJa_R | IFJ_R | cingulate_vlPFC_R | Glasser2016 (Petre2023 volumetric projection) |
| **259** | Area IF Jp (Right) | Ctx_IFJp_R | Ctx_IFJp_R | IFJ_R | cingulate_vlPFC_R | Glasser2016 (Petre2023 volumetric projection) |
| **260** | Area IF Sp (Right) | Ctx_IFSp_R | Ctx_IFSp_R | IFS_R | cingulate_vlPFC_R | Glasser2016 (Petre2023 volumetric projection) |
| **261** | Area IFSa (Right) | Ctx_IFSa_R | Ctx_IFSa_R | IFS_R | cingulate_vlPFC_R | Glasser2016 (Petre2023 volumetric projection) |
| **262** | Area posterior 9-46v (Right) | Ctx_p9_46v_R | Ctx_p9_46v_R | BA46_R | cingulate_dlPFC_R | Glasser2016 (Petre2023 volumetric projection) |
| **263** | Area 46 (Right) | Ctx_46_R | Ctx_46_R | BA46_R | cingulate_dlPFC_R | Glasser2016 (Petre2023 volumetric projection) |

| | | | | | | |
|---|---|---|---|---|---|---|
| **264** | Area anterior 9-46v (Right) | Ctx_a9_46v_R | Ctx_a9_46v_R | BA46_R | cingulate_dlPFC_R | Glasser2016 (Petre2023 volumetric projection) |
| **265** | Area 9-46d (Right) | Ctx_9_46d_R | Ctx_9_46d_R | BA46_R | cingulate_dlPFC_R | Glasser2016 (Petre2023 volumetric projection) |
| **266** | Area 9 anterior (Right) | Ctx_9a_R | Ctx_9a_R | BA9_R | cingulate_dlPFC_R | Glasser2016 (Petre2023 volumetric projection) |
| **267** | Area 10v (Right) | Ctx_10v_R | Ctx_10v_R | BA10_R | cingulate_ventral_frontal_R | Glasser2016 (Petre2023 volumetric projection) |
| **268** | Area anterior 10p (Right) | Ctx_a10p_R | Ctx_a10p_R | BA10_R | cingulate_ventral_frontal_R | Glasser2016 (Petre2023 volumetric projection) |
| **269** | Polar 10p (Right) | Ctx_10pp_R | Ctx_10pp_R | BA10_R | cingulate_ventral_frontal_R | Glasser2016 (Petre2023 volumetric projection) |
| **270** | Area 11l (Right) | Ctx_11l_R | Ctx_11l_R | BA11_R | cingulate_ventral_frontal_R | Glasser2016 (Petre2023 volumetric projection) |
| **271** | Area 13l (Right) | Ctx_13l_R | Ctx_13l_R | BA13_R | cingulate_ventral_frontal_R | Glasser2016 (Petre2023 volumetric projection) |
| **272** | Orbital Frontal Complex (Right) | Ctx_OFC_R | Ctx_OFC_R | OFC_R | cingulate_ventral_frontal_R | Glasser2016 (Petre2023 |

| | | | | | | |
|---|---|---|---|---|---|---|
| | | | | | | volumetric projection) |
| **273** | Area 47s (Right) | Ctx_47s_R | Ctx_47s_R | BA47_R | cingulate_ventral_frontal_R | Glasser2016 (Petre2023 volumetric projection) |
| **274** | Area Lateral IntraParietal dorsal (Right) | Ctx_LIPd_R | Ctx_LIPd_R | ventral_intraparietal_R | parietal_superior_lobule_R | Glasser2016 (Petre2023 volumetric projection) |
| **275** | Area 6 anterior (Right) | Ctx_6a_R | Ctx_6a_R | superior_premotor_R | somatomotor_premotor_R | Glasser2016 (Petre2023 volumetric projection) |
| **276** | Inferior 6-8 Transitional Area (Right) | Ctx_i6_8_R | Ctx_i6_8_R | Inferior_lateral_BA8_R | cingulate_dlPFC_R | Glasser2016 (Petre2023 volumetric projection) |
| **277** | Superior 6-8 Transitional Area (Right) | Ctx_s6_8_R | Ctx_s6_8_R | Superior_lateral_BA8_R | cingulate_dlPFC_R | Glasser2016 (Petre2023 volumetric projection) |
| **278** | Putative primary gustatory cortex (Right) | Ctx_43_R | Ctx_43_R | FOP1-BA43_R | somatomotor_operculum_R | Glasser2016 (Petre2023 volumetric projection) |
| **279** | Area OP4/PV (Right) | Ctx_OP4_R | Ctx_OP4_R | OP4_R | somatomotor_operculum_R | Glasser2016 (Petre2023 volumetric projection) |
| **280** | Area OP1/SII (Right) | Ctx_OP1_R | Ctx_OP1_R | SII+_R | somatomotor_operculum_R | Glasser2016 (Petre2023 volumetric projection) |

| | | | | | | |
|---|---|---|---|---|---|---|
| **281** | Area OP2-3/VS (Right) | Ctx_OP2_3_R | Ctx_OP2_3_R | SII+_R | somatomotor_operculum_R | Glasser2016 (Petre2023 volumetric projection) |
| **282** | Area 52 (Right) | Ctx_52_R | Ctx_52_R | posterior_granular_insula_R | insula_posterior_R | Glasser2016 (Petre2023 volumetric projection) |
| **283** | RetroInsular Cortex (Right) | Ctx_RI_R | Ctx_RI_R | auditory+_R | auditory_early_R | Glasser2016 (Petre2023 volumetric projection) |
| **284** | Area Pfcm (Right) | Ctx_PFcm_R | Ctx_PFcm_R | SII+_R | somatomotor_operculum_R | Glasser2016 (Petre2023 volumetric projection) |
| **285** | insula_posteriorr Area 2 (Right) | Ctx_PoI2_R | Ctx_PoI2_R | inferior_dysgranular_insula_R | insula_posterior_R | Glasser2016 (Petre2023 volumetric projection) |
| **286** | Area TA2 (Right) | Ctx_TA2_R | Ctx_TA2_R | auditory_transition_area_R | auditory_association_cortex_R | Glasser2016 (Petre2023 volumetric projection) |
| **287** | Frontal Opercular Area 4 (Right) | Ctx_FOP4_R | Ctx_FOP4_R | anterior_operculum_R | insula_operculum_R | Glasser2016 (Petre2023 volumetric projection) |
| **288** | Middle Insular Area (Right) | Ctx_MI_R | Ctx_MI_R | anterior_agranular_insula_R | insula_anterior_R | Glasser2016 (Petre2023 volumetric projection) |
| **289** | Piriform olfactory cortex (Right) | Ctx_Pir_R | Ctx_Pir_R | PIR_R | insula_anterior_R | Glasser2016 (Petre2023 |

| | | | | | | |
|---|---|---|---|---|---|---|
| | | | | | | volumetric projection) |
| **290** | Anterior Ventral Insular Area (Right) | Ctx_AVI_R | Ctx_AVI_R | anterior_agranular_insula_R | insula_anterior_R | Glasser2016 (Petre2023 volumetric projection) |
| **291** | Anterior Agranular Insular Complex (Right) | Ctx_AAIC_R | Ctx_AAIC_R | anterior_agranular_insula_R | insula_anterior_R | Glasser2016 (Petre2023 volumetric projection) |
| **292** | Frontal Opercular Area 1 (Right) | Ctx_FOP1_R | Ctx_FOP1_R | FOP1-BA43_R | somatomotor_operculum_R | Glasser2016 (Petre2023 volumetric projection) |
| **293** | Frontal Opercular Area 3 (Right) | Ctx_FOP3_R | Ctx_FOP3_R | posterior_operculum_R | insula_operculum_R | Glasser2016 (Petre2023 volumetric projection) |
| **294** | Frontal Opercular Area 2 (Right) | Ctx_FOP2_R | Ctx_FOP2_R | posterior_operculum_R | insula_operculum_R | Glasser2016 (Petre2023 volumetric projection) |
| **295** | Parietal area F part t (Right) | Ctx_PFt_R | Ctx_PFt_R | anterior_IPL_R | parietal_inferior_lobule_R | Glasser2016 (Petre2023 volumetric projection) |
| **296** | Anterior IntraParietal Area (Right) | Ctx_AIP_R | Ctx_AIP_R | ventral_intraparietal_R | parietal_superior_lobule_R | Glasser2016 (Petre2023 volumetric projection) |
| **297** | Entorhinal Cortex (Right) | Ctx_EC_R | Ctx_EC_R | hippocampal_formation_R | temporal_medial_R | Glasser2016 (Petre2023 volumetric projection) |

| | | | | | | |
|---|---|---|---|---|---|---|
| **298** | PreSubiculum (Right) | Ctx_PreS_R | Ctx_PreS_R | hippocampal_formation_R | temporal_medial_R | Glasser2016 (Petre2023 volumetric projection) |
| **299** | Prostriate Area (Right) | Ctx_ProS_R | Ctx_ProS_R | parietal_occipital_sulcus_R | cingulate_posterior_R | Glasser2016 (Petre2023 volumetric projection) |
| **300** | Perirhinal Ectorhinal Cortex (Right) | Ctx_PeEc_R | Ctx_PeEc_R | PeEc_R | temporal_medial_R | Glasser2016 (Petre2023 volumetric projection) |
| **301** | Area STGa (Right) | Ctx_STGa_R | Ctx_STGa_R | auditory_transition_area_R | auditory_association_cortex_R | Glasser2016 (Petre2023 volumetric projection) |
| **302** | ParaBelt Complex (Right) | Ctx_PBelt_R | Ctx_PBelt_R | auditory+_R | auditory_early_R | Glasser2016 (Petre2023 volumetric projection) |
| **303** | Auditory 5 Complex (Right) | Ctx_A5_R | Ctx_A5_R | auditory_transition_area_R | auditory_association_cortex_R | Glasser2016 (Petre2023 volumetric projection) |
| **304** | ParaHippocampal Area 1 (Right) | Ctx_PHA1_R | Ctx_PHA1_R | PHA_R | temporal_medial_R | Glasser2016 (Petre2023 volumetric projection) |
| **305** | ParaHippocampal Area 3 (Right) | Ctx_PHA3_R | Ctx_PHA3_R | PHA_R | temporal_medial_R | Glasser2016 (Petre2023 volumetric projection) |
| **306** | Area STSd anterior (Right) | Ctx_STSda_R | Ctx_STSda_R | anterior_AAC_R | auditory_association_cortex_R | Glasser2016 (Petre2023 |

| | | | | | | |
|---|---|---|---|---|---|---|
| | | | | | | volumetric projection) |
| **307** | Area STSd posterior (Right) | Ctx_STSdp_R | Ctx_STSdp_R | posterior_AAC_R | auditory_association_cortex_R | Glasser2016 (Petre2023 volumetric projection) |
| **308** | Area STSv posterior (Right) | Ctx_STSvp_R | Ctx_STSvp_R | posterior_AAC_R | auditory_association_cortex_R | Glasser2016 (Petre2023 volumetric projection) |
| **309** | Area TG dorsal (Right) | Ctx_TGd_R | Ctx_TGd_R | temporal_pole_R | temporal_lateral_R | Glasser2016 (Petre2023 volumetric projection) |
| **310** | Area TE1 anterior (Right) | Ctx_TE1a_R | Ctx_TE1a_R | temporal_pole_R | temporal_lateral_R | Glasser2016 (Petre2023 volumetric projection) |
| **311** | Area TE1 posterior (Right) | Ctx_TE1p_R | Ctx_TE1p_R | middle_temporal_gyrus_R | temporal_lateral_R | Glasser2016 (Petre2023 volumetric projection) |
| **312** | Area TE2 anterior (Right) | Ctx_TE2a_R | Ctx_TE2a_R | inferior_temporal_sulcus_R | temporal_lateral_R | Glasser2016 (Petre2023 volumetric projection) |
| **313** | Area TF (Right) | Ctx_TF_R | Ctx_TF_R | inferior_temporal_sulcus_R | temporal_lateral_R | Glasser2016 (Petre2023 volumetric projection) |
| **314** | Area TE2 posterior (Right) | Ctx_TE2p_R | Ctx_TE2p_R | inferior_temporal_sulcus_R | temporal_lateral_R | Glasser2016 (Petre2023 volumetric projection) |

| | | | | | | |
|---|---|---|---|---|---|---|
| **315** | Area PHT (Right) | Ctx_PHT_R | Ctx_PHT_R | middle_temporal_gyrus_R | temporal_lateral_R | Glasser2016 (Petre2023 volumetric projection) |
| **316** | Area PH (Right) | Ctx_PH_R | Ctx_PH_R | PH_R | visual_MT+_R | Glasser2016 (Petre2023 volumetric projection) |
| **317** | Area TemporoParietoOccipital Junction 1 (Right) | Ctx_TPOJ1_R | Ctx_TPOJ1_R | TPOJ_R | parietal_TPOJ_R | Glasser2016 (Petre2023 volumetric projection) |
| **318** | Area TemporoParietoOccipital Junction 2 (Right) | Ctx_TPOJ2_R | Ctx_TPOJ2_R | TPOJ_R | parietal_TPOJ_R | Glasser2016 (Petre2023 volumetric projection) |
| **319** | Area TemporoParietoOccipital Junction 3 (Right) | Ctx_TPOJ3_R | Ctx_TPOJ3_R | TPOJ_R | parietal_TPOJ_R | Glasser2016 (Petre2023 volumetric projection) |
| **320** | Dorsal Transitional Visual Area (Right) | Ctx_DVT_R | Ctx_DVT_R | parietal_occipital_sulcus_R | cingulate_posterior_R | Glasser2016 (Petre2023 volumetric projection) |
| **321** | Parietal area G posterior (Right) | Ctx_PGp_R | Ctx_PGp_R | posterior_IPL_R | parietal_inferior_lobule_R | Glasser2016 (Petre2023 volumetric projection) |
| **322** | Area IntraParietal 2 (Right) | Ctx_IP2_R | Ctx_IP2_R | superior_IPL_R | parietal_inferior_lobule_R | Glasser2016 (Petre2023 volumetric projection) |
| **323** | Area IntraParietal 1 (Right) | Ctx_IP1_R | Ctx_IP1_R | superior_IPL_R | parietal_inferior_lobule_R | Glasser2016 (Petre2023 |

| | | | | | | |
|---|---|---|---|---|---|---|
| | | | | | | volumetric projection) |
| **324** | Area IntraParietal 0 (Right) | Ctx_IP0_R | Ctx_IP0_R | posterior_IPL_R | parietal_inferior_lobule_R | Glasser2016 (Petre2023 volumetric projection) |
| **325** | Parietal area F operculum (Right) | Ctx_PFop_R | Ctx_PFop_R | anterior_IPL_R | parietal_inferior_lobule_R | Glasser2016 (Petre2023 volumetric projection) |
| **326** | Parietal area F (Right) | Ctx_PF_R | Ctx_PF_R | anterior_IPL_R | parietal_inferior_lobule_R | Glasser2016 (Petre2023 volumetric projection) |
| **327** | Parietal area F part m (Right) | Ctx_PFm_R | Ctx_PFm_R | middle_IPL_R | parietal_inferior_lobule_R | Glasser2016 (Petre2023 volumetric projection) |
| **328** | Parietal area G inferior (Right) | Ctx_PGi_R | Ctx_PGi_R | inferior_IPL_R | parietal_inferior_lobule_R | Glasser2016 (Petre2023 volumetric projection) |
| **329** | Parietal area G superior (Right) | Ctx_PGs_R | Ctx_PGs_R | inferior_IPL_R | parietal_inferior_lobule_R | Glasser2016 (Petre2023 volumetric projection) |
| **330** | Area V6A (Right) | Ctx_V6A_R | Ctx_V6A_R | V6_R | visual_dorsal_R | Glasser2016 (Petre2023 volumetric projection) |
| **331** | VentroMedial Visual Area 1 (Right) | Ctx_VMV1_R | Ctx_VMV1_R | VMV_R | visual_ventral_R | Glasser2016 (Petre2023 volumetric projection) |

| | | | | | | |
|---|---|---|---|---|---|---|
| **332** | VentroMedial Visual Area 3 (Right) | Ctx_VMV3_R | Ctx_VMV3_R | VMV_R | visual_ventral_R | Glasser2016 (Petre2023 volumetric projection) |
| **333** | ParaHippocampal Area 2 (Right) | Ctx_PHA2_R | Ctx_PHA2_R | PHA_R | temporal_medial_R | Glasser2016 (Petre2023 volumetric projection) |
| **334** | Area V4t (Right) | Ctx_V4t_R | Ctx_V4t_R | vMT+_R | visual_MT+_R | Glasser2016 (Petre2023 volumetric projection) |
| **335** | Fundus of the superior temporal area (Right) | Ctx_FST_R | Ctx_FST_R | vMT+_R | visual_MT+_R | Glasser2016 (Petre2023 volumetric projection) |
| **336** | Area v3CD (Right) | Ctx_V3CD_R | Ctx_V3CD_R | supplementary_V3_R | visual_dorsal_R | Glasser2016 (Petre2023 volumetric projection) |
| **337** | Area Lateral Occipital 3 (Right) | Ctx_LO3_R | Ctx_LO3_R | LO_R | visual_MT+_R | Glasser2016 (Petre2023 volumetric projection) |
| **338** | VentroMedial Visual Area 2 (Right) | Ctx_VMV2_R | Ctx_VMV2_R | VMV_R | visual_ventral_R | Glasser2016 (Petre2023 volumetric projection) |
| **339** | Area 31pd (Right) | Ctx_31pd_R | Ctx_31pd_R | precuneus_R | cingulate_posterior_R | Glasser2016 (Petre2023 volumetric projection) |
| **340** | Area 31a (Right) | Ctx_31a_R | Ctx_31a_R | anterior_precuneus_R | cingulate_posterior_R | Glasser2016 (Petre2023 |

| | | | | | | |
|---|---|---|---|---|---|---|
| | | | | | | volumetric projection) |
| **341** | Ventral Visual Complex (Right) | Ctx_VVC_R | Ctx_VVC_R | VVC_R | visual_ventral_R | Glasser2016 (Petre2023 volumetric projection) |
| **342** | Area 25 (Right) | Ctx_25_R | Ctx_25_R | infralimbic_R | cingulate_ACC_mPFC_R | Glasser2016 (Petre2023 volumetric projection) |
| **343** | Area s32 (Right) | Ctx_s32_R | Ctx_s32_R | vmPFC_R | cingulate_ACC_mPFC_R | Glasser2016 (Petre2023 volumetric projection) |
| **344** | posterior OFC Complex (Right) | Ctx_pOFC_R | Ctx_pOFC_R | OFC_R | cingulate_ventral_frontal_R | Glasser2016 (Petre2023 volumetric projection) |
| **345** | Area insula_posteriorr 1 (Right) | Ctx_PoI1_R | Ctx_PoI1_R | inferior_dysgranular_insula_R | insula_posterior_R | Glasser2016 (Petre2023 volumetric projection) |
| **346** | Insular Granular Complex (Right) | Ctx_Ig_R | Ctx_Ig_R | posterior_granular_insula_R | insula_posterior_R | Glasser2016 (Petre2023 volumetric projection) |
| **347** | Area Frontal Opercular 5 (Right) | Ctx_FOP5_R | Ctx_FOP5_R | anterior_operculum_R | insula_operculum_R | Glasser2016 (Petre2023 volumetric projection) |
| **348** | Area posterior 10p (Right) | Ctx_p10p_R | Ctx_p10p_R | BA10_R | cingulate_ventral_frontal_R | Glasser2016 (Petre2023 volumetric projection) |

| | | | | | | |
|---|---|---|---|---|---|---|
| **349** | Area posterior 47r (Right) | Ctx_p47r_R | Ctx_p47r_R | BA47_R | cingulate_ventral_frontal_R | Glasser2016 (Petre2023 volumetric projection) |
| **350** | Area TG Ventral (Right) | Ctx_TGv_R | Ctx_TGv_R | temporal_pole_R | temporal_lateral_R | Glasser2016 (Petre2023 volumetric projection) |
| **351** | Medial Belt Complex (Right) | Ctx_MBelt_R | Ctx_MBelt_R | auditory+_R | auditory_early_R | Glasser2016 (Petre2023 volumetric projection) |
| **352** | Lateral Belt Complex (Right) | Ctx_LBelt_R | Ctx_LBelt_R | auditory+_R | auditory_early_R | Glasser2016 (Petre2023 volumetric projection) |
| **353** | Auditory 4 Complex (Right) | Ctx_A4_R | Ctx_A4_R | auditory_transition_area_R | auditory_association_cortex_R | Glasser2016 (Petre2023 volumetric projection) |
| **354** | Area STSv anterior (Right) | Ctx_STSva_R | Ctx_STSva_R | anterior_AAC_R | auditory_association_cortex_R | Glasser2016 (Petre2023 volumetric projection) |
| **355** | Area TE1 Middle (Right) | Ctx_TE1m_R | Ctx_TE1m_R | middle_temporal_gyrus_R | temporal_lateral_R | Glasser2016 (Petre2023 volumetric projection) |
| **356** | Para-Insular Area (Right) | Ctx_PI_R | Ctx_PI_R | inferior_dysgranular_insula_R | insula_posterior_R | Glasser2016 (Petre2023 volumetric projection) |
| **357** | Area anterior 32 prime (Right) | Ctx_a32pr_R | Ctx_a32pr_R | midcingulate_R | cingulate_ACC_mPFC_R | Glasser2016 (Petre2023 |

| | | | | | | |
|---|---|---|---|---|---|---|
| | | | | | | volumetric projection) |
| **358** | Area posterior 24 (Right) | Ctx_p24_R | Ctx_p24_R | midcingulate_R | cingulate_ACC_mPFC_R | Glasser2016 (Petre2023 volumetric projection) |
| **359** | Left hippocampal cornu ammonis 2 | MTL_Hipp_CA2_L | MTL_Hipp_CA23_L | Hipp_L | Hippocampal_Formation_L | Julich/Amunts |
| **360** | Left hippocampal cornu ammonis 3 | MTL_Hipp_CA3_L | MTL_Hipp_CA23_L | Hipp_L | Hippocampal_Formation_L | Julich/Amunts |
| **361** | Left hippocampal dentate gyrus | MTL_Hipp_DG_L | MTL_Hipp_DG_L | Hipp_L | Hippocampal_Formation_L | Julich/Amunts |
| **362** | Left hippocampal cornu ammonis 1 | MTL_Hipp_CA1_L | MTL_Hipp_CA1_L | Hipp_L | Hippocampal_Formation_L | Julich/Amunts |
| **363** | Left hippocampal subiculum | MTL_Hipp_Subiculum_L | MTL_Hipp_Subiculum_L | Subiculum_L | Hippocampal_Formation_L | Julich/Amunts |
| **364** | Right hippocampal cornu ammonis 2 | MTL_Hipp_CA2_R | MTL_Hipp_CA23_R | Hipp_R | Hippocampal_Formation_R | Julich/Amunts |
| **365** | Right hippocampal cornu ammonis 3 | MTL_Hipp_CA3_R | MTL_Hipp_CA23_R | Hipp_R | Hippocampal_Formation_R | Julich/Amunts |
| **366** | Right hippocampal dentate gyrus | MTL_Hipp_DG_R | MTL_Hipp_DG_R | Hipp_R | Hippocampal_Formation_R | Julich/Amunts |
| **367** | Right hippocampal cornu ammonis 1 | MTL_Hipp_CA1_R | MTL_Hipp_CA1_R | Hipp_R | Hippocampal_Formation_R | Julich/Amunts |
| **368** | Right hippocampal subiculum | MTL_Hipp_Subiculum_R | MTL_Hipp_Subiculum_R | Subiculum_R | Hippocampal_Formation_R | Julich/Amunts |
| **369** | Region 1 from SUIT_Cerebellum_MNI152NLin2009cAsym | Cblm_I_IV_L | Cblm_I_IV_L | Cblm_I_IV_V_VI_L | Cblm_cortex_L | SUIT/Diedrichsen |
| **370** | Region 2 from SUIT_Cerebellum_MNI152NLin2009cAsym | Cblm_I_IV_R | Cblm_I_IV_R | Cblm_I_IV_V_VI_R | Cblm_cortex_R | SUIT/Diedrichsen |
| **371** | Region 3 from SUIT_Cerebellum_MNI152NLin2009cAsym | Cblm_V_L | Cblm_V_L | Cblm_I_IV_V_VI_L | Cblm_cortex_L | SUIT/Diedrichsen |
| **372** | Region 4 from SUIT_Cerebellum_MNI152NLin2009cAsym | Cblm_V_R | Cblm_V_R | Cblm_I_IV_V_VI_R | Cblm_cortex_R | SUIT/Diedrichsen |
| **373** | Region 5 from SUIT_Cerebellum_MNI152NLin2009cAsym | Cblm_VI_L | Cblm_VI_L | Cblm_I_IV_V_VI_L | Cblm_cortex_L | SUIT/Diedrichsen |
| **374** | Region 6 from SUIT_Cerebellum_MNI152NLin2009cAsym | Cblm_Vermis_VI | Cblm_Vermis_VI | Cblm_vermis | Cblm_vermis | SUIT/Diedrichsen |
| **375** | Region 7 from SUIT_Cerebellum_MNI152NLin2009cAsym | Cblm_VI_R | Cblm_VI_R | Cblm_I_IV_V_VI_R | Cblm_cortex_R | SUIT/Diedrichsen |

| | | | | | | |
|---|---|---|---|---|---|---|
| **376** | Region 8 from SUIT_Cerebellum_MNI152NLin2009cAsym | Cblm_CrusI_L | Cblm_CrusI_L | Cblm_CrusI_CrusII_VIIIb_L | Cblm_cortex_L | SUIT/Diedrichsen |
| **377** | Region 10 from SUIT_Cerebellum_MNI152NLin2009cAsym | Cblm_CrusI_R | Cblm_CrusI_R | Cblm_CrusI_CrusII_VIIIb_R | Cblm_cortex_R | SUIT/Diedrichsen |
| **378** | Region 11 from SUIT_Cerebellum_MNI152NLin2009cAsym | Cblm_CrusII_L | Cblm_CrusII_L | Cblm_CrusI_CrusII_VIIIb_L | Cblm_cortex_L | SUIT/Diedrichsen |
| **379** | Region 12 from SUIT_Cerebellum_MNI152NLin2009cAsym | Cblm_Vermis_CrusII | Cblm_Vermis_CrusII | Cblm_vermis | Cblm_vermis | SUIT/Diedrichsen |
| **380** | Region 13 from SUIT_Cerebellum_MNI152NLin2009cAsym | Cblm_CrusII_R | Cblm_CrusII_R | Cblm_CrusI_CrusII_VIIIb_R | Cblm_cortex_R | SUIT/Diedrichsen |
| **381** | Region 14 from SUIT_Cerebellum_MNI152NLin2009cAsym | Cblm_VIIb_L | Cblm_VIIb_L | Cblm_CrusI_CrusII_VIIIb_L | Cblm_cortex_L | SUIT/Diedrichsen |
| **382** | Region 15 from SUIT_Cerebellum_MNI152NLin2009cAsym | Cblm_Vermis_VIIb | Cblm_Vermis_VIIb | Cblm_vermis | Cblm_vermis | SUIT/Diedrichsen |
| **383** | Region 16 from SUIT_Cerebellum_MNI152NLin2009cAsym | Cblm_VIIb_R | Cblm_VIIb_R | Cblm_CrusI_CrusII_VIIIb_R | Cblm_cortex_R | SUIT/Diedrichsen |
| **384** | Region 17 from SUIT_Cerebellum_MNI152NLin2009cAsym | Cblm_VIIIa_L | Cblm_VIIIa_L | Cblm_VIIIa_VIIb_IX_X_L | Cblm_cortex_L | SUIT/Diedrichsen |
| **385** | Region 18 from SUIT_Cerebellum_MNI152NLin2009cAsym | Cblm_Vermis_VIIIa | Cblm_Vermis_VIIIa | Cblm_vermis | Cblm_vermis | SUIT/Diedrichsen |
| **386** | Region 19 from SUIT_Cerebellum_MNI152NLin2009cAsym | Cblm_VIIIa_R | Cblm_VIIIa_R | Cblm_VIIIa_VIIb_IX_X_R | Cblm_cortex_R | SUIT/Diedrichsen |
| **387** | Region 20 from SUIT_Cerebellum_MNI152NLin2009cAsym | Cblm_VIIIb_L | Cblm_VIIIb_L | Cblm_VIIIa_VIIb_IX_X_L | Cblm_cortex_L | SUIT/Diedrichsen |
| **388** | Region 21 from SUIT_Cerebellum_MNI152NLin2009cAsym | Cblm_Vermis_VIIIb | Cblm_Vermis_VIIIb | Cblm_vermis | Cblm_vermis | SUIT/Diedrichsen |
| **389** | Region 22 from SUIT_Cerebellum_MNI152NLin2009cAsym | Cblm_VIIIb_R | Cblm_VIIIb_R | Cblm_VIIIa_VIIb_IX_X_R | Cblm_cortex_R | SUIT/Diedrichsen |
| **390** | Region 23 from SUIT_Cerebellum_MNI152NLin2009cAsym | Cblm_IX_L | Cblm_IX_L | Cblm_VIIIa_VIIb_IX_X_L | Cblm_cortex_L | SUIT/Diedrichsen |
| **391** | Region 24 from SUIT_Cerebellum_MNI152NLin2009cAsym | Cblm_Vermis_IX | Cblm_Vermis_IX | Cblm_vermis | Cblm_vermis | SUIT/Diedrichsen |
| **392** | Region 25 from SUIT_Cerebellum_MNI152NLin2009cAsym | Cblm_IX_R | Cblm_IX_R | Cblm_VIIIa_VIIb_IX_X_R | Cblm_cortex_R | SUIT/Diedrichsen |
| **393** | Region 26 from SUIT_Cerebellum_MNI152NLin2009cAsym | Cblm_X_L | Cblm_X_L | Cblm_VIIIa_VIIb_IX_X_L | Cblm_cortex_L | SUIT/Diedrichsen |
| **394** | Region 27 from SUIT_Cerebellum_MNI152NLin2009cAsym | Cblm_Vermis_X | Cblm_Vermis_X | Cblm_vermis | Cblm_vermis | SUIT/Diedrichsen |
| **395** | Region 28 from SUIT_Cerebellum_MNI152NLin2009cAsym | Cblm_X_R | Cblm_X_R | Cblm_VIIIa_VIIb_IX_X_R | Cblm_cortex_R | SUIT/Diedrichsen |

| | | | | | | |
|---|---|---|---|---|---|---|
| **396** | Extended Amygdala: Bed Nuclei of the Stria Terminalis (BNST) and Sublenticular Extended Amygdala (SLEA) (left) | BG_BST_SLEA_L | BG_BST_SLEA_L | VStriatum_L | VStriatum_L | CIT168 subcortical v1.1.0 |
| **397** | Extended Amygdala: Bed Nuclei of the Stria Terminalis (BNST) and Sublenticular Extended Amygdala (SLEA) (right) | BG_BST_SLEA_R | BG_BST_SLEA_R | VStriatum_R | VStriatum_R | CIT168 subcortical v1.1.0 |
| **398** | Region 45 from tian_3t_fmriprep20 | BG_CAU_DA_L | BG_CAU_DA_L | aCAU_L | CAU_L | Tian2020 |
| **399** | Region 18 from tian_3t_fmriprep20 | BG_CAU_DA_R | BG_CAU_DA_R | aCAU_R | CAU_R | Tian2020 |
| **400** | Region 44 from tian_3t_fmriprep20 | BG_CAU_VA_L | BG_CAU_VA_L | aCAU_L | CAU_L | Tian2020 |
| **401** | Region 17 from tian_3t_fmriprep20 | BG_CAU_VA_R | BG_CAU_VA_R | aCAU_R | CAU_R | Tian2020 |
| **402** | Region 46 from tian_3t_fmriprep20 | BG_CAU_body_L | BG_CAU_body_L | pCAU_L | CAU_L | Tian2020 |
| **403** | Region 19 from tian_3t_fmriprep20 | BG_CAU_body_R | BG_CAU_body_R | pCAU_R | CAU_R | Tian2020 |
| **404** | Region 47 from tian_3t_fmriprep20 | BG_CAU_tail_L | BG_CAU_tail_L | pCAU_L | CAU_L | Tian2020 |
| **405** | Region 20 from tian_3t_fmriprep20 | BG_CAU_tail_R | BG_CAU_tail_R | pCAU_R | CAU_R | Tian2020 |
| **406** | Globus Pallidus external segment (left) | BG_GPe_L | BG_GPe_L | GPe_L | GP_L | CIT168 subcortical v1.1.0 |
| **407** | Globus Pallidus external segment (right) | BG_GPe_R | BG_GPe_R | GPe_R | GP_R | CIT168 subcortical v1.1.0 |
| **408** | Globus Pallidus internal segment (left) | BG_GPi_L | BG_GPi_L | GPi_L | GP_L | CIT168 subcortical v1.1.0 |
| **409** | Globus Pallidus internal segment (right) | BG_GPi_R | BG_GPi_R | GPi_R | GP_R | CIT168 subcortical v1.1.0 |
| **410** | Nucleus Accumbens, putative core (left) | BG_NAc_core_L | BG_NAc_L | VStriatum_L | VStriatum_L | Cartmell2019 |
| **411** | Nucleus Accumbens, putative core (right) | BG_NAc_core_R | BG_NAc_R | VStriatum_R | VStriatum_R | Cartmell2019 |
| **412** | Nucleus Accumbens, putative shell (left) | BG_NAc_shell_L | BG_NAc_L | VStriatum_L | VStriatum_L | Cartmell2019 |
| **413** | NucleusAccumbens, putative shell (right) | BG_NAc_shell_R | BG_NAc_R | VStriatum_R | VStriatum_R | Cartmell2019 |
| **414** | Region 41 from tian_3t_fmriprep20 | BG_PUT_DA_L | BG_PUT_DA_L | aPUT_L | PUT_L | Tian2020 |
| **415** | Region 14 from tian_3t_fmriprep20 | BG_PUT_DA_R | BG_PUT_DA_R | aPUT_R | PUT_R | Tian2020 |
| **416** | Region 43 from tian_3t_fmriprep20 | BG_PUT_DP_L | BG_PUT_DP_L | pPUT_L | PUT_L | Tian2020 |
| **417** | Region 16 from tian_3t_fmriprep20 | BG_PUT_DP_R | BG_PUT_DP_R | pPUT_R | PUT_R | Tian2020 |

| | | | | | | |
|---|---|---|---|---|---|---|
| **418** | Region 40 from tian_3t_fmriprep20 | BG_PUT_VA_L | BG_PUT_VA_L | aPUT_L | PUT_L | Tian2020 |
| **419** | Region 13 from tian_3t_fmriprep20 | BG_PUT_VA_R | BG_PUT_VA_R | aPUT_R | PUT_R | Tian2020 |
| **420** | Region 42 from tian_3t_fmriprep20 | BG_PUT_VP_L | BG_PUT_VP_L | pPUT_L | PUT_L | Tian2020 |
| **421** | Region 15 from tian_3t_fmriprep20 | BG_PUT_VP_R | BG_PUT_VP_R | pPUT_R | PUT_R | Tian2020 |
| **422** | Ventral pallidum (left) | BG_VeP_L | BG_GPe_L | GPe_L | GP_L | CIT168 subcortical v1.1.0 |
| **423** | Ventral pallidum (right) | BG_VeP_R | BG_GPe_R | GPe_R | GP_R | CIT168 subcortical v1.1.0 |
| **424** | Anteroventral_(left) | Thal_AV_L | Thal_AV_L | Thal_anterior_L | Thal_Anterior_L | Iglesias2018 |
| **425** | central_medial_(left) | Thal_CeM_L | Thal_rIL_L | Thal_Intralaminar-antoventral_L | Thal_Intralaminar_L | Iglesias2018 |
| **426** | central_lateral_(left) | Thal_CL_L | Thal_CL_L | Thal_Intralaminar-anterodorsal_L | Thal_Intralaminar_L | Iglesias2018 |
| **427** | centromedian_(left) | Thal_CM_L | Thal_cIL_L | Thal_Intralaminar-posterior_L | Thal_Intralaminar_L | Iglesias2018 |
| **428** | Laterodorsal_(left) | Thal_LD_L | Thal_LD_L | Thal_dorsal_L | Thal_Lateral_L | Iglesias2018 |
| **429** | Lateral_posterior_(left) | Thal_LP_L | Thal_LP_L | Thal_dorsal_L | Thal_Lateral_L | Iglesias2018 |
| **430** | Limitans_(suprageniculate)_(left) | Thal_SG_L_L | Thal_PuM_L | Thal_Pulvinar_L | Thal_Posterior_L | Iglesias2018 |
| **431** | Mediodorsal_medial_parvocellular-like_(left) | Thal_MDl_L | Thal_MD_L | Thal_Medial_L | Thal_Medial_L | Iglesias2018 |
| **432** | Mediodorsal_medial_magnocellular-like_(left) | Thal_MDm_L | Thal_MD_L | Thal_Medial_L | Thal_Medial_L | Iglesias2018 |
| **433** | parafascicular_(left) | Thal_Pf_L | Thal_cIL_L | Thal_Intralaminar-posterior_L | Thal_Intralaminar_L | Iglesias2018 |
| **434** | Pulvinar_anterior_(left) | Thal_PuA_L | Thal_PuA_L | Thal_Pulvinar_L | Thal_Posterior_L | Iglesias2018 |
| **435** | Pulvinar_inferior_(left) | Thal_PuI_L | Thal_PuL_L | Thal_Pulvinar_L | Thal_Posterior_L | Iglesias2018 |
| **436** | Pulvinar_lateral_(left) | Thal_PuL_L | Thal_PuL_L | Thal_Pulvinar_L | Thal_Posterior_L | Iglesias2018 |
| **437** | Ventral_anterior_(left) | Thal_VA_L | Thal_VA_L | Thal_Lateral-rostral_L | Thal_Ventral_L | Iglesias2018 |
| **438** | Ventral_anterior_magnocellular_(left) | Thal_VAmc_L | Thal_VA_L | Thal_Lateral-rostral_L | Thal_Ventral_L | Iglesias2018 |

| | | | | | | |
|---|---|---|---|---|---|---|
| **439** | ventral_lateral_anterior_(left) | Thal_VLa_L | Thal_VL_L | Thal_Lateral-rostral_L | Thal_Ventral_L | Iglesias2018 |
| **440** | ventral_lateral_posterior_(left) | Thal_VLp_L | Thal_VL_L | Thal_Lateral-rostral_L | Thal_Ventral_L | Iglesias2018 |
| **441** | ventral_posterolateral_and_ventral_posteromedial_(left) | Thal_VPL_VPM_L | Thal_VPL_VPM_L | Thal_Lateral-caudal_L | Thal_Ventral_L | Iglesias2018 |
| **442** | Pulvinar_medial_(medial_subdivision)_(left) | Thal_PuMm_L | Thal_PuM_L | Thal_Pulvinar_L | Thal_Posterior_L | Iglesias2018 |
| **443** | Pulvinar_medial_(lateral_subdivision)_(left) | Thal_PuMl_L | Thal_PuM_L | Thal_Pulvinar_L | Thal_Posterior_L | Iglesias2018 |
| **444** | Anteroventral_(right) | Thal_AV_R | Thal_AV_R | Thal_anterior_R | Thal_Anterior_R | Iglesias2018 |
| **445** | central_medial_(right) | Thal_CeM_R | Thal_rIL_R | Thal_Intralaminar-antoventral_R | Thal_Intralaminar_R | Iglesias2018 |
| **446** | central_lateral_(right) | Thal_CL_R | Thal_CL_R | Thal_Intralaminar-anterodorsal_R | Thal_Intralaminar_R | Iglesias2018 |
| **447** | centromedian_(right) | Thal_CM_R | Thal_cIL_R | Thal_Intralaminar-posterior_R | Thal_Intralaminar_R | Iglesias2018 |
| **448** | Laterodorsal_(right) | Thal_LD_R | Thal_LD_R | Thal_dorsal_R | Thal_Lateral_R | Iglesias2018 |
| **449** | Lateral_posterior_(right) | Thal_LP_R | Thal_LP_R | Thal_dorsal_R | Thal_Lateral_R | Iglesias2018 |
| **450** | Limitans_(suprageniculate)_(right) | Thal_SG_R_L | Thal_PuM_R | Thal_Pulvinar_R | Thal_Posterior_R | Iglesias2018 |
| **451** | Mediodorsal_medial_parvocellular-like_(right) | Thal_MDl_R | Thal_MD_R | Thal_Medial_R | Thal_Medial_R | Iglesias2018 |
| **452** | Mediodorsal_medial_magnocellular-like_(right) | Thal_MDm_R | Thal_MD_R | Thal_Medial_R | Thal_Medial_R | Iglesias2018 |
| **453** | parafascicular_(right) | Thal_Pf_R | Thal_cIL_R | Thal_Intralaminar-posterior_R | Thal_Intralaminar_R | Iglesias2018 |
| **454** | Pulvinar_anterior_(right) | Thal_PuA_R | Thal_PuA_R | Thal_Pulvinar_R | Thal_Posterior_R | Iglesias2018 |
| **455** | Pulvinar_inferior_(right) | Thal_PuI_R | Thal_PuL_R | Thal_Pulvinar_R | Thal_Posterior_R | Iglesias2018 |
| **456** | Pulvinar_lateral_(right) | Thal_PuL_R | Thal_PuL_R | Thal_Pulvinar_R | Thal_Posterior_R | Iglesias2018 |
| **457** | Ventral_anterior_(right) | Thal_VA_R | Thal_VA_R | Thal_Lateral-rostral_R | Thal_Ventral_R | Iglesias2018 |
| **458** | Ventral_anterior_magnocellular_(right) | Thal_VAmc_R | Thal_VA_R | Thal_Lateral-rostral_R | Thal_Ventral_R | Iglesias2018 |
| **459** | ventral_lateral_anterior_(right) | Thal_VLa_R | Thal_VL_R | Thal_Lateral-rostral_R | Thal_Ventral_R | Iglesias2018 |

| | | | | | | |
|---|---|---|---|---|---|---|
| **460** | ventral_lateral_posterior_(right) | Thal_VLp_R | Thal_VL_R | Thal_Lateral-rostral_R | Thal_Ventral_R | Iglesias2018 |
| **461** | ventral_posterolateral_and_ventral_posteromedial_(right) | Thal_VPL_VPM_R | Thal_VPL_VPM_R | Thal_Lateral-caudal_R | Thal_Ventral_R | Iglesias2018 |
| **462** | Pulvinar_medial_(medial_subdivision)_(right) | Thal_PuMm_R | Thal_PuM_R | Thal_Pulvinar_R | Thal_Posterior_R | Iglesias2018 |
| **463** | Pulvinar_medial_(lateral_subdivision)_(right) | Thal_PuMl_R | Thal_PuM_R | Thal_Pulvinar_R | Thal_Posterior_R | Iglesias2018 |
| **464** | Habenula (left) | Epithal_Haben_L | Thal_PuM_L | Thal_Pulvinar_L | Thal_Posterior_L | CIT168 v1.1.0 subcortical |
| **465** | Habenula (right) | Epithal_Haben_R | Thal_PuM_R | Thal_Pulvinar_R | Thal_Posterior_R | CIT168 v1.1.0 subcortical |
| **466** | suprachiasmatic nucleus, supraoptic nucleus (left) | hypothalamus_anterior_inferior_L | hypothalamus_anterior_and_tubular_inferior_L | hypothalamus_L | hypothalamus_L | Billot/Iglesias2020 |
| **467** | preoptic area, paraventricular nucleus (left) | hypothalamus_anterior_superior_L | hypothalamus_anterior_and_tubular_superior_L | hypothalamus_L | hypothalamus_L | Billot/Iglesias2020 |
| **468** | mamillary body (medial and lateral nuclei), lateral hypothalamus, tuberomamillary nucleus (left) | hypothalamus_posterior_L | hypothalamus_posterior_L | hypothalamus_L | hypothalamus_L | Billot/Iglesias2020 |
| **469** | infundibular nucleus, ventromedial nucleus, supraoptic nucleus, lateral tubular nucleus, tuberomamillary nucleus (left) | hypothalamus_tubular_inferior_L | hypothalamus_anterior_and_tubular_inferior_L | hypothalamus_L | hypothalamus_L | Billot/Iglesias2020 |
| **470** | dorsomedial nucleus, paraventricular nucleus, lateral hypothalamus (left) | hypothalamus_tubular_superior_L | hypothalamus_anterior_and_tubular_superior_L | hypothalamus_L | hypothalamus_L | Billot/Iglesias2020 |
| **471** | suprachiasmatic nucleus, supraoptic nucleus (right) | hypothalamus_anterior_inferior_R | hypothalamus_anterior_and_tubular_inferior_R | hypothalamus_R | hypothalamus_R | Billot/Iglesias2020 |
| **472** | preoptic area, paraventricular nucleus (right) | hypothalamus_anterior_superior_R | hypothalamus_anterior_and_tubular_superior_R | hypothalamus_R | hypothalamus_R | Billot/Iglesias2020 |
| **473** | mamillary body (medial and lateral nuclei), lateral hypothalamus, tuberomamillary nucleus (right) | hypothalamus_posterior_R | hypothalamus_posterior_R | hypothalamus_R | hypothalamus_R | Billot/Iglesias2020 |
| **474** | infundibular nucleus, ventromedial nucleus, supraoptic nucleus, lateral tubular nucleus, tuberomamillary nucleus (right) | hypothalamus_tubular_inferior_R | hypothalamus_anterior_and_tubular_inferior_R | hypothalamus_R | hypothalamus_R | Billot/Iglesias2020 |
| **475** | dorsomedial nucleus, paraventricular nucleus, lateral hypothalamus (right) | hypothalamus_tubular_superior_R | hypothalamus_anterior_and_tubular_superior_R | hypothalamus_R | hypothalamus_R | Billot/Iglesias2020 |
| **476** | Parabrachial Pigmented Nucleus (left) | BStem_PBP_L | BStem_VTA_PBP_L | VTA_PBP_SN_L | Midbrain_L | CIT168 subcortical v1.1.0 |

| | | | | | | |
|---|---|---|---|---|---|---|
| **477** | Ventral Tegmental Area (left) | BStem_VTA_L | BStem_VTA_PBP_L | VTA_PBP_SN_L | Midbrain_L | CIT168 subcortical v1.1.0 |
| **478** | Subthalamic nucleus (left) | BStem_STH_R | BStem_STH_L | STH_L | Midbrain_L | CIT168 subcortical v1.1.0 |
| **479** | Parabrachial Pigmented Nucleus (right) | BStem_PBP_R | BStem_VTA_PBP_R | VTA_PBP_SN_R | Midbrain_R | CIT168 subcortical v1.1.0 |
| **480** | Ventral Tegmental Area (right) | BStem_VTA_R | BStem_VTA_PBP_R | VTA_PBP_SN_R | Midbrain_R | CIT168 subcortical v1.1.0 |
| **481** | Subthalamic nucleus (right) | BStem_STH_R | BStem_STH_R | STH_R | Midbrain_R | CIT168 subcortical v1.1.0 |
| **482** | Reuniens_(medial_ventral)_(left) | Thal_MV_Re_L | Thal_rIL_L | Thal_Intralaminar-antoventral_L | Thal_Intralaminar_L | Iglesias2018 |
| **483** | Reuniens_(medial_ventral)_(right) | Thal_MV_Re_R | Thal_rIL_R | Thal_Intralaminar-antoventral_R | Thal_Intralaminar_R | Iglesias2018 |
| **484** | Amygdala basolateral complex: lateral nucleus;Intercalated nuclei most proximal to basolateral complex: lateral nucleus; (left) | MTL_AMY_La_L | MTL_AMY_La_L | AMY_La_L | Amygdala_L | CIT168 amydala v1.0.3 |
| **485** | Amygdala basolateral complex: basal nucleus; basolateral complex: accessory basal nucleus; basolateral complex: paralaminar nucleus;Intercalated nuclei most proximal to basolateral complex: basal nucleus;Intercalated nuclei most proximal to basolateral complex: accessory basal nucleus;Intercalated nuclei most proximal to basolateral complex: paralaminar nucleus; (left) | MTL_AMY_BL_L | MTL_AMY_BL_L | AMY_BL_CeM_L | Amygdala_L | CIT168 amydala v1.0.3 |
| **486** | Amygdala central nucleus; corticomedial group: medial nucleus; corticomedial group: periamygdaloid cortex;Amygdalostriatal triansition area;Anteroir amygdaloid area;Intercalated nuclei most proximal to central nucleus;Intercalated nuclei most proximal to corticomedial group: medial nucleus;Intercalated nuclei most proximal to corticomedial group: periamygdaloid cortex;Intercalated nuclei most proximal to | MTL_AMY_CeM_L | MTL_AMY_CeM_L | AMY_BL_CeM_L | Amygdala_L | CIT168 amydala v1.0.3 |

| | | | | | | |
|---|---|---|---|---|---|---|
| | Amygdalostriatal triansition area;Intercalated nuclei most proximal to Anteroir amygdaloid area; (left) | | | | | |
| **487** | Amygdala basolateral complex: lateral nucleus;Intercalated nuclei most proximal to basolateral complex: lateral nucleus; (right) | MTL_AMY_La_R | MTL_AMY_La_R | AMY_La_R | Amygdala_R | CIT168 amydala v1.0.3 |
| **488** | Amygdala basolateral complex: basal nucleus; basolateral complex: accessory basal nucleus; basolateral complex: paralaminar nucleus;Intercalated nuclei most proximal to basolateral complex: basal nucleus;Intercalated nuclei most proximal to basolateral complex: accessory basal nucleus;Intercalated nuclei most proximal to basolateral complex: paralaminar nucleus; (right) | MTL_AMY_BL_R | MTL_AMY_BL_R | AMY_BL_CeM_R | Amygdala_R | CIT168 amydala v1.0.3 |
| **489** | Amygdala central nucleus; corticomedial group: medial nucleus; corticomedial group: periamygdaloid cortex;Amygdalostriatal triansition area;Anteroir amygdaloid area;Intercalated nuclei most proximal to central nucleus;Intercalated nuclei most proximal to corticomedial group: medial nucleus;Intercalated nuclei most proximal to corticomedial group: periamygdaloid cortex;Intercalated nuclei most proximal to Amygdalostriatal triansition area;Intercalated nuclei most proximal to Anteroir amygdaloid area; (right) | MTL_AMY_CeM_R | MTL_AMY_CeM_R | AMY_BL_CeM_R | Amygdala_R | CIT168 amydala v1.0.3 |
| **490** | lateral_geniculate_(left) | Thal_LGN_L | Thal_LGN_L | Thal_LGN_L | Thal_Posterior_L | Iglesias2018 |
| **491** | medial_geniculate_(left) | Thal_MGN_L | Thal_MGN_L | Thal_MGN_L | Thal_Posterior_L | Iglesias2018 |
| **492** | lateral_geniculate_(right) | Thal_LGN_R | Thal_LGN_R | Thal_LGN_R | Thal_Posterior_R | Iglesias2018 |
| **493** | medial_geniculate_(right) | Thal_MGN_R | Thal_MGN_R | Thal_MGN_R | Thal_Posterior_R | Iglesias2018 |
| **494** | Region 1 from Kragel2019PAG | BStem_dmPAG | BStem_PAG | PAG | Midbrain | Kragel2019 |
| **495** | Region 2 from Kragel2019PAG | BStem_vlPAG_L | BStem_PAG | PAG | Midbrain | Kragel2019 |

| | | | | | | |
|---|---|---|---|---|---|---|
| **496** | Region 3 from Kragel2019PAG | BStem_lPAG_L | BStem_PAG | PAG | Midbrain | Kragel2019 |
| **497** | Region 4 from Kragel2019PAG | BStem_vlPAG_R | BStem_PAG | PAG | Midbrain | Kragel2019 |
| **498** | Region 5 from Kragel2019PAG | BStem_lPAG_R | BStem_PAG | PAG | Midbrain | Kragel2019 |
| **499** | Dorsal Raphe Nucleus | BStem_DR_B7 | BStem_DR_B7 | Rostral Raphe (Serotonergic) | Midbrain | Levinson-Bari Limbic Brainstem Atlas |
| **500** | Locus Coreleus (left) | BStem_LC_L | BStem_LC+_L | LC+_L | Pons_L | Levinson-Bari Limbic Brainstem Atlas |
| **501** | Locus Coreleus (right) | BStem_LC_R | BStem_LC+_R | LC+_R | Pons_R | Levinson-Bari Limbic Brainstem Atlas |
| **502** | Substantia Nigra pars compacta (left) | BStem_SNc_L | BStem_SN_L | VTA_PBP_SN_L | Midbrain_L | CIT168 subcortical v1.1.0 |
| **503** | Red nucleus (left) | BStem_RN_L | BStem_RN_L | RN_L | Midbrain_L | CIT168 subcortical v1.1.0 |
| **504** | Substantia Nigra pars reticulata (left) | BStem_SNr_L | BStem_SN_L | VTA_PBP_SN_L | Midbrain_L | CIT168 subcortical v1.1.0 |
| **505** | Substantia Nigra pars compacta (right) | BStem_SNc_R | BStem_SN_R | VTA_PBP_SN_R | Midbrain_R | CIT168 subcortical v1.1.0 |
| **506** | Red nucleus (right) | BStem_RN_R | BStem_RN_R | RN_R | Midbrain_R | CIT168 subcortical v1.1.0 |
| **507** | Substantia Nigra pars reticulata (right) | BStem_SNr_R | BStem_SN_R | VTA_PBP_SN_R | Midbrain_R | CIT168 subcortical v1.1.0 |
| **508** | Caudal-rostral linear raphe | BStem_CLi_RLi | BStem_CLi_RLi | Rostral Raphe (Serotonergic) | Midbrain | Bianciardi brainstem navigator v.0.9 (ref: 2021) |
| **509** | Left cuneiform nucleus | BStem_CnF_L | BStem_PAG | PAG | Midbrain | Bianciardi brainstem |

| | | | | | | |
|---|---|---|---|---|---|---|
| | | | | | | navigator v.0.9 (ref: 2018) |
| **510** | Right cuneiform nucleus | BStem_CnF_R | BStem_PAG | PAG | Midbrain | Bianciardi brainstem navigator v.0.9 (ref: 2018) |
| **511** | Left inferior colliculus | BStem_IC_L | BStem_IC_L | Tectum_L | Midbrain_L | Bianciardi brainstem navigator v.0.9 (ref: 2019) |
| **512** | Right inferior colliculus | BStem_IC_R | BStem_IC_R | Tectum_R | Midbrain_R | Bianciardi brainstem navigator v.0.9 (ref: 2019) |
| **513** | Left inferior medullary reticular formation (lateral part) | BStem_iMRtl_L | BStem_iMRt_L | Medullary reticular formation_L | Medulla_L | Bianciardi brainstem navigator v.0.9 (ref: 2022) |
| **514** | Right inferior medullary reticular formation (lateral part) | BStem_iMRtl_R | BStem_iMRt_R | Medullary reticular formation_R | Medulla_R | Bianciardi brainstem navigator v.0.9 (ref: 2022) |
| **515** | Left inferior medullary reticular formation (medial part) | BStem_iMRtm_L | BStem_iMRt_L | Medullary reticular formation_L | Medulla_L | Bianciardi brainstem navigator v.0.9 (ref: 2022) |
| **516** | Right inferior medullary reticular formation (medial part) | BStem_iMRtm_R | BStem_iMRt_R | Medullary reticular formation_R | Medulla_R | Bianciardi brainstem navigator v.0.9 (ref: 2022) |
| **517** | Left inferior olivary nucleus | BStem_ION_L | BStem_OC_L | Olivary complex_L | Medulla_L | Bianciardi brainstem navigator v.0.9 (ref: 2015) |

| | | | | | | |
|---|---|---|---|---|---|---|
| **518** | Right inferior olivary nucleus | BStem_ION_R | BStem_OC_R | Olivary complex_R | Medulla_R | Bianciardi brainstem navigator v.0.9 (ref: 2015) |
| **519** | Left isthmic reticular formation | BStem_isRt_L | BStem_isRt_L | Rostral reticular formation_L | Midbrain_L | Bianciardi brainstem navigator v.0.9 (ref: 2021) |
| **520** | Right isthmic reticular formation | BStem_isRt_R | BStem_isRt_R | Rostral reticular formation_R | Midbrain_R | Bianciardi brainstem navigator v.0.9 (ref: 2021) |
| **521** | Left laterodorsal tegmental nucleus - central gray of the rhomboencephalon | BStem_LDTg_CGPn_L | BStem_LDTg_CGPn_L | Cholinergic nuclei_L | Pons_L | Bianciardi brainstem navigator v.0.9 (ref: 2022) |
| **522** | Right laterodorsal tegmental nucleus - central gray of the rhomboencephalon | BStem_LDTg_CGPn_R | BStem_LDTg_CGPn_R | Cholinergic nuclei_R | Pons_R | Bianciardi brainstem navigator v.0.9 (ref: 2022) |
| **523** | Left lateral parabrachial nucleus | BStem_LPB_L | BStem_LPB_L | Parabrachial nuclei_L | Pons_L | Bianciardi brainstem navigator v.0.9 (ref: 2020) |
| **524** | Right lateral parabrachial nucleus | BStem_LPB_R | BStem_LPB_R | Parabrachial nuclei_R | Pons_R | Bianciardi brainstem navigator v.0.9 (ref: 2020) |
| **525** | Left microcellular tegmental nucleus – prabigeminal nucleus | BStem_MiTg_PBG_L | BStem_MiTg_PBG_L | Tectum_L | Midbrain_L | Bianciardi brainstem navigator v.0.9 (ref: 2021) |
| **526** | Right microcellular tegmental nucleus – prabigeminal nucleus | BStem_MiTg_PBG_R | BStem_MiTg_PBG_R | Tectum_R | Midbrain_R | Bianciardi brainstem navigator |

| | | | | | | |
|---|---|---|---|---|---|---|
| | | | | | | v.0.9 (ref: 2021) |
| **527** | Median raphe | BStem_MnR_B6_B8 | BStem_MnR | Rostral Raphe (Serotonergic) | Midbrain | Bianciardi brainstem navigator v.0.9 (ref: 2015) |
| **528** | Left medial parabrachial nucleus | BStem_MPB_L | BStem_MPB_L | Parabrachial nuclei_L | Pons_L | Bianciardi brainstem navigator v.0.9 (ref: 2020) |
| **529** | Right medial parabrachial nucleus | BStem_MPB_R | BStem_MPB_R | Parabrachial nuclei_R | Pons_R | Bianciardi brainstem navigator v.0.9 (ref: 2020) |
| **530** | Left mesencephalic reticular formation (anterior part) | BStem_mRta_L | BStem_mRt_L | Rostral reticular formation_L | Midbrain_L | Bianciardi brainstem navigator v.0.9 (ref: 2021) |
| **531** | Right mesencephalic reticular formation (anterior part) | BStem_mRta_R | BStem_mRt_R | Rostral reticular formation_R | Midbrain_R | Bianciardi brainstem navigator v.0.9 (ref: 2021) |
| **532** | Left mesencephalic reticular formation (dorsal part) | BStem_mRtd_L | BStem_mRt_L | Rostral reticular formation_L | Midbrain_L | Bianciardi brainstem navigator v.0.9 (ref: 2021) |
| **533** | Right mesencephalic reticular formation (dorsal part) | BStem_mRtd_R | BStem_mRt_R | Rostral reticular formation_R | Midbrain_R | Bianciardi brainstem navigator v.0.9 (ref: 2021) |
| **534** | Left mesencephalic reticular formation (lateral part) | BStem_mRtl_L | BStem_mRt_L | Rostral reticular formation_L | Midbrain_L | Bianciardi brainstem navigator v.0.9 (ref: 2021) |

| | | | | | | |
|---|---|---|---|---|---|---|
| **535** | Right mesencephalic reticular formation (lateral part) | BStem_mRtl_R | BStem_mRt_R | Rostral reticular formation_R | Midbrain_R | Bianciardi brainstem navigator v.0.9 (ref: 2021) |
| **536** | Left parvicellular reticular nucleus alpha part | BStem_PCRtA_L | BStem_PCRtA_L | Medullary reticular formation_L | Medulla_L | Bianciardi brainstem navigator v.0.9 (ref: 2022) |
| **537** | Right parvicellular reticular nucleus alpha part | BStem_PCRtA_R | BStem_PCRtA_R | Medullary reticular formation_R | Medulla_R | Bianciardi brainstem navigator v.0.9 (ref: 2022) |
| **538** | Paramedian nucleus | BStem_PMnR_B6_B8 | BStem_MnR | Rostral Raphe (Serotonergic) | Midbrain | Bianciardi brainstem navigator v.0.9 (ref: 2018) |
| **539** | Left pontine reticular nucleus, oral and caudal parts (pontis oralis and caudalis) | BStem_PnO_PnC_B5_L | BStem_PnO_PnC_L | L_PnO_PnC_L | Pons_L | Bianciardi brainstem navigator v.0.9 (ref: 2022) |
| **540** | Right pontine reticular nucleus, oral and caudal parts (pontis oralis and caudalis) | BStem_PnO_PnC_B5_R | BStem_PnO_PnC_R | R_PnO_PnC_R | Pons_R | Bianciardi brainstem navigator v.0.9 (ref: 2022) |
| **541** | Left pedunculotegmental nucleus (also called pedunculopontine nucleus) | BStem_PTg_L | BStem_PTg_L | Cholinergic nuclei_L | Pons_L | Bianciardi brainstem navigator v.0.9 (ref: 2018) |
| **542** | Right pedunculotegmental nucleus (also called pedunculopontine nucleus) | BStem_PTg_R | BStem_PTg_R | Cholinergic nuclei_R | Pons_R | Bianciardi brainstem navigator v.0.9 (ref: 2018) |
| **543** | Raphe magnus | BStem_RMg_B3 | BStem_RObPaMg | Medullary Raphe (Serotonergic) | Medulla | Bianciardi brainstem navigator |

| | | | | | | |
|---|---|---|---|---|---|---|
| | | | | | | v.0.9 (ref: 2015) |
| **544** | Raphe obscurus | BStem_ROb_B2 | BStem_RObPaMg | Medullary Raphe (Serotonergic) | Medulla | Bianciardi brainstem navigator v.0.9 (ref: 2022) |
| **545** | Raphe pallidus | BStem_RPa_B1 | BStem_RObPaMg | Medullary Raphe (Serotonergic) | Medulla | Bianciardi brainstem navigator v.0.9 (ref: 2022) |
| **546** | Left superior colliculus | BStem_SC_L | BStem_SC_L | Tectum_L | Midbrain_L | Bianciardi brainstem navigator v.0.9 (ref: 2019) |
| **547** | Right superior colliculus | BStem_SC_R | BStem_SC_R | Tectum_R | Midbrain_R | Bianciardi brainstem navigator v.0.9 (ref: 2019) |
| **548** | Left superior medullary reticular formation (lateral part) | BStem_sMRtl_L | BStem_sMRt_L | Medullary reticular formation_L | Medulla_L | Bianciardi brainstem navigator v.0.9 (ref: 2022) |
| **549** | Right superior medullary reticular formation (lateral part) | BStem_sMRtl_R | BStem_sMRt_R | Medullary reticular formation_R | Medulla_R | Bianciardi brainstem navigator v.0.9 (ref: 2022) |
| **550** | Left superior medullary reticular formation (medial part) | BStem_sMRtm_L | BStem_sMRt_L | Medullary reticular formation_L | Medulla_L | Bianciardi brainstem navigator v.0.9 (ref: 2022) |
| **551** | Right superior medullary reticular formation (medial part) | BStem_sMRtm_R | BStem_sMRt_R | Medullary reticular formation_R | Medulla_R | Bianciardi brainstem navigator v.0.9 (ref: 2022) |

| | | | | | | |
|---|---|---|---|---|---|---|
| **552** | Left superior olivary complex | BStem_SOC_L | BStem_OC_L | Olivary complex_L | Medulla_L | Bianciardi brainstem navigator v.0.9 (ref: 2019) |
| **553** | Right superior olivary complex | BStem_SOC_R | BStem_OC_R | Olivary complex_R | Medulla_R | Bianciardi brainstem navigator v.0.9 (ref: 2019) |
| **554** | Left subcoeruleus | BStem_SubC_L | BStem_LC+_L | LC+_L | Pons_L | Bianciardi brainstem navigator v.0.9 (ref: 2022) |
| **555** | Right subcoeruleus | BStem_SubC_R | BStem_LC+_R | LC+_R | Pons_R | Bianciardi brainstem navigator v.0.9 (ref: 2022) |
| **556** | Left vestibular nuclei complex | BStem_Ve_L | BStem_Ve_L | Cranial_nuclei_L | Medulla_L | Bianciardi brainstem navigator v.0.9 (ref: 2020) |
| **557** | Right vestibular nuclei complex | BStem_Ve_R | BStem_Ve_R | Cranial_nuclei_R | Medulla_R | Bianciardi brainstem navigator v.0.9 (ref: 2020) |
| **558** | Left viscero-sensory-motor nuclei complex | BStem_VSM_L | BStem_VSM_L | Cranial_nuclei_L | Medulla_L | Bianciardi brainstem navigator v.0.9 (ref: 2020) |
| **559** | Right viscero-sensory-motor nuclei complex | BStem_VSM_R | BStem_VSM_R | Cranial_nuclei_R | Medulla_R | Bianciardi brainstem navigator v.0.9 (ref: 2020) |
| **560** | Midbrain left rostral dorsal | BStem_Midbrd_L | BStem_Shen_Midb_Lrd | Shen_Midb_Lrd | Midbrain_L | Shen268 |
| **561** | Midbrain left caudal | BStem_Midbc_L | BStem_Shen_Midb_Lc | Shen_Midb_Lc | Midbrain_L | Shen268 |

| | | | | | | |
|---|---|---|---|---|---|---|
| **562** | Midbrain right rostral dorsal | BStem_Midbrd_R | BStem_Shen_Midb_Rrd | Shen_Midb_Rrd | Midbrain_R | Shen268 |
| **563** | Midbrain right caudal dorsal | BStem_Midbcd_R | BStem_Shen_Midb_Rcd | Shen_Midb_Rcd | Midbrain_R | Shen268 |
| **564** | Pons left rostral dorsal | BStem_Ponsrd_L | BStem_Shen_Pons_Lrd | Shen_Pons_Lrd | Pons_L | Shen268 |
| **565** | Pons left cudal dorsal | BStem_Ponscd_L | BStem_Shen_Pons_Lcd | Shen_Pons_Lcd | Pons_L | Shen268 |
| **566** | Pons left ventral | BStem_Ponsv_L | BStem_Shen_Pons_Lv | Shen_Pons_Lv | Pons_L | Shen268 |
| **567** | Pons right rostral ventral | BStem_Ponsrv_R | BStem_Shen_Pons_Rrv | Shen_Pons_Rrv | Pons_R | Shen268 |
| **568** | Pons right caudal ventral | BStem_Ponscv_R | BStem_Shen_Pons_Rcv | Shen_Pons_Rcv | Pons_R | Shen268 |
| **569** | Pons right caudal dorsal | BStem_Ponscd_R | BStem_Shen_Pons_Rcd | Shen_Pons_Rcd | Pons_R | Shen268 |
| **570** | Medulla left | BStem_Med_L | BStem_Shen_Med_L | Shen_Med_L | Medulla_L | Shen268 |
| **571** | Medulla right | BStem_Med_R | BStem_Shen_Med_R | Shen_Med_R | Medulla_R | Shen268 |

Supplementary Table 3: Brain parcels and their atlas labels included in the whole-brain parcel-wise analysis.

| Brain Regions | Kogler et al., 2015 | Mothersill et al., 2016 | Berretz et al., 2021 | Qiu et al., 2022 | Neurosynth | Frequency | $f_{sig}/f_{total}$ | CanlabAtlas 2024 |
|---|---|---|---|---|---|---|---|---|
| Inferior frontal gyrus/ventral lateral prefrontal cortex | 1 | | 1 | 1 | 1 | 4 | 80% | label2: Ctx_44, Ctx_45, Ctx_IFSp |
| Medial prefrontal cortex | | | | | 1 | 1 | 20% | |
| Anterior mid-cingulate cortex | | | | | 1 | 1 | 20% | |
| Posterior cingulate cortex | | | | | 1 | 1 | 20% | |
| Ventral medial prefrontal cortex | | | | | 1 | 1 | 20% | |
| Anterior insula | 1 | 1 | 1 | 1 | 1 | 5 | 100% | label3: anterior_agranular_insula |
| Precentral/Premotor Cortex | | | | 1 | 1 | 2 | 40% | |
| Inferior temporal gyrus | | | 1 | | | 1 | 20% | |
| Middle temporal gyrus | | | 1 | | | 1 | 20% | |
| Superior temporal gyrus | 1 | | 1 | | | 2 | 40% | |
| Para-hippocampal cortex | | | 1 | | 1 | 2 | 40% | |
| Fusiform gyrus | | | | 1 | | 1 | 20% | |
| Occipital Gyrus/V3 | | | 1 | 1 | | 2 | 40% | |
| Amygdala | | 1 | | 1 | 1 | 3 | 60% | labels4: Amygdala |
| Hippocampus | | | | | 1 | 1 | 20% | |
| Caudate | | | | | 1 | 2 | 40% | |
| Putamen | | | | | 1 | 2 | 40% | |
| Accumbens | | | | | 1 | 1 | 20% | |
| Thalamus | | 1 | | 1 | 1 | 3 | 60% | labels4: Thal |

| | | | | | | | |
|---|---|---|---|---|---|---|---|
| Brain Stem | | | | 1 | 1 | 2 | 40% |

Supplementary Table 4: Brain regions showing significance in stress meta-analyses. Selected regions of interest (ROIs) are highlighted

| **Brain Regions** | **Kim et al., 2015** | **Boissoneault et al., 2016** | **Wortinger et al., 2016/2017** | **Boissoneault et al., 2018** | **Manca et al., 2021** | **Li et al., 2022** | **Su et al., 2023** | $f_{total}$ | **CanlabAtlas 2024** |
|---|---|---|---|---|---|---|---|---|---|
| aMCC | | 1 | | | 1 | | 1 | 3 | labels2: Ctx_33pr, Ctx_a24pr, Ctx_p32p, Ctx_a32pr |
| OFC | | | | | 1 | | | 1 | |
| Frontal pole | | | | | 1 | 1 | | 2 | labels2: Ctx_10d, Ctx_a10p, Ctx_p10p |
| aINS | | | | | 1 (INS) | 1 | | 2 | labels3: anterior_agranular_insula |
| pINS | | 1 | 1 | | 1 (INS) | | | 3 | labels4: insula_posterior |
| Premotor | | 1 | | | 1 | 1 | | 3 | labels2: Ctx_FEF, Ctx_PEF, Ctx_55b, Ctx_6d, Ctx_6v, Ctx_6r, Ctx_6a |
| Somatomotor_paracentral | | 1 | | | | 1 | | 2 | labels3: cingulate_motor_area, SMA, BA5 |
| dlPFC | | 1 (SFG) | | | 1 | 1 | | 3 | labels3: BA9, BA46 |
| dmPFC | 1 | 1 (SFG) | | 1 | 1 | | | 4 | labels3: dmPFC |
| Operculum | | | | | | 1 | | 1 | |
| M1 | | | 1 | | 1 | | | 2 | labels2: Ctx_4 |
| S1 | | | | | 1 | 1 | | 2 | labels3: S1 |
| PCC/Precuneus | 1 | 1 | | 1 | 1 | 1 | 1 | 6 | labels2: Ctx_23d, Ctx_v23ab, Ctx_d23ab, Ctx_31pv, Ctx_23c, Ctx_31pd, Ctx_31a |
| SPL | | 1 | 1 | | | | | 2 | labels4: parietal_superior_lobule |
| IPL | | 1 | | 1 | 1 | 1 | | 4 | labels4: parietal_inferior_lobule |
| TPOJ | | | 1 | | 1 | | | 2 | labels3: TPOJ |
| Early Auditory | | | 1 | | | | | 1 | |
| AAC | | | | | 1 | 1 | | 2 | labels3: anterior_AAC, posterior_AAC |
| STG | | | | | 1 | | | 1 | |
| MTG | 1 | | | | 1 | | | 2 | labels3: middle_temporal_gyrus |
| ParaHPC | | 1 | | | | | | 1 | |
| Inferior Occipital | | | | | 1 | | | 1 | |

| | | | | | | | | | |
|---|---|---|---|---|---|---|---|---|---|
| V1 | | 1 | | | | | | 1 | |
| V3 | | | | | | 1 | | 1 | |
| V8 | | | | | 1 | | | 1 | |
| Globus Pallidus | | 1 | | | 1 | | | 2 | labels4: GP |
| Putamen | | | 1 | | | | | 1 | |
| Thalamus | | | 1 | | 1 | | | 2 | labels4: Thal |
| Hypothalamus | | | | | 1 | | | 1 | |
| Caudate | | | 1 | | 1 | | | 2 | labels4: CAU |
| Medulla | | | | | 1 | | | 1 | |
| Pons | | | | | | | 1 | 1 | |

Supplementary Table 5: Brain regions showing altered resting state functional connectivity in ME/CFS studies. Selected regions of interest (ROIs) are highlighted. aMCC, anterior mid-cingulate cortex; OFC, orbitofrontal cortex; aINS/pINS, anterior/posterior insula; dl/dmPFC, dorsal lateral/dorsal medial prefrontal cortex; PCC, posterior cingulate cortex; SPL/IPL, superior/inferior parietal lobe; TPOJ, temporal parietal occipital junction; AAC, auditory association cortex; STG/MTG, superior/middle temporal gyrus; ParaHPC, para-hippocampal gyrus, M1, primary motor cortex; S1, primary somatosensory cortex.

| Brain regions | Hannestad et al., 2013 | Holmes et al., 2018 | Li et al., 2018 | Richards et al., 2018 | Setiawan et al., 2015 | Setiawan et al., 2018 | Joo et al., 2021 | Cakmak et al., 2022 | f | $f_{sig}/f_{defined}$(%) | CanlabAtlas 2024 |
|---|---|---|---|---|---|---|---|---|---|---|---|
| | ROI | ROI | ROI | ROI | ROI | ROI | ROI | ROI | | | |
| aMCC$_{sig}$ | | 1 | 1 | 1 | ACC... | ACC... | ACC... | | 6 | 75% | labels2: Ctx_33pr, Ctx_a24pr, Ctx_p32pr,Ctx_a32pr |
| aMCC$_{defined}$ | Frontal | 1 | Frontal | 1 | 1 | 1 | ACC... | 1 | 8 | | |
| sgACC$_{sig}$ | | | 1 | 1 | | | | 1 | 3 | 75% | labels2: Ctx_25 |
| sgACC$_{defined}$ | Frontal | | Frontal | 1 | | | | 1 | 4 | | |
| pgACC$_{sig}$ | | | | | | | | | 0 | 0% | |
| pgACC$_{defined}$ | Frontal | | Frontal | | | | | 1 | 3 | | |
| OFC$_{sig}$ | | | 1 | | 1 | 1 | | | 3 | 60% | |
| OFC$_{defined}$ | Frontal | | Frontal | | 1 | 1 | 1 | | 5 | | |
| Frontal pole$_{sig}$ | | | 1 | | 1 | 1 | | | 3 | 60% | |
| Frontal pole$_{defined}$ | Frontal | SFG | Frontal | | 1 | 1 | | | 5 | | |
| dmPFC$_{sig}$ | | | 1 | | 1 | 1 | | | 3 | 60% | |
| dmPFC$_{defined}$ | Frontal | SFG | Frontal | | 1 | 1 | | | 5 | | |
| dlPFC$_{sig}$ | | | 1 | | 1 | 1 | | | 3 | 50% | |
| dlPFC$_{defined}$ | Frontal | MFG | Frontal | | 1 | 1 | 1 | | 6 | | |
| vlPFC/IFG$_{sig}$ | | | 1 | | 1 | 1 | | | 3 | 60% | |
| vlPFC/IFG$_{defined}$ | | MFG/IFG | Frontal | | 1 | 1 | 1 | | 5 | | |
| PCC$_{sig}$ | | | | | | | 1 | | 1 | 100% biased | |
| PCC$_{defined}$ | | | | | | | 1 | | 1 | | |
| aINS & pINS$_{sig}$ | | 1 | | | 1 | 1 | | 1 | 4 | 80% | labels4: insula_posterior; label3: anterior_agranular_insula |

| | | | | | | | | | | | |
|---|---|---|---|---|---|---|---|---|---|---|---|
| aINS & pINS$_{defined}$ | | 1 | | | 1 | 1 | 1 | 1 | 5 | | |
| Temporal cortex$_{sig}$ | | | 1 | | | 1 | | | 2 | 40% | |
| Temporal cortex$_{defined}$ | 1 | | 1 | | 1 | 1 | 1 | | 5 | | |
| ParaHPC$_{sig}$ | | | | | | | | | 1 | 100% biased | |
| ParaHPC$_{defined}$ | | | | | | | | | 1 | | |
| Parietal$_{sig}$ | | | | | | 1 | | | 1 | 33% | |
| Parietal$_{defined}$ | 1 | | | | 1 | 1 | | | 3 | | |
| Occipital$_{sig}$ | | | | | | 1 | | | 1 | 33% | |
| Occipital$_{defined}$ | 1 | | | | 1 | 1 | | | 3 | | |
| Thalamus$_{sig}$ | | | | | 1 | 1 | | | 2 | 67% | |
| Thalamus$_{defined}$ | 1 | | | | 1 | 1 | | | 3 | | |
| Hipp$_{sig}$ | | | 1 | | 1 | 1 | | | 3 | 75% | label4: Hippocampal_Formation |
| Hipp$_{defined}$ | | | 1 | | 1 | 1 | 1 | | 4 | | |
| Putamen$_{sig}$ | | | | | 1 | 1 | | | 2 | 67% | |
| Putamen$_{defined}$ | 1 | | | | 1 | 1 | | | 3 | | |
| Caudate$_{sig}$ | | | | | | 1 | | | 1 | 33% | |
| Caudate$_{defined}$ | 1 | | | | 1 | 1 | | | 3 | | |
| Striatum$_{sig}$ | | | | | 1 | 1 | | | 2 | 100% biased | |
| Striatum$_{defined}$ | | | | | 1 | 1 | | | 2 | | |
| Cerebellum$_{sig}$ | | | | | | | | | 0 | 0% | |
| Cerebellum$_{defined}$ | 1 | | | | | | | | 1 | | |

Supplementary Table 6: Brain regions showing significant case-control differences in major depressive disorder TSPO studies. Selected regions of interest (ROIs) are highlighted. aMCC, anterior mid-cingulate cortex; sgACC, subgenual anterior cingulate cortex; aINS/pINS, anterior/posterior insula; Hipp, Hippocampus.

| **Brain regions** | **Albrecht et al., 2019a** | **Albrecht et al., 2019b** | **Albrecht et al., 2021** | **Hadjikhani et al., 2020** | **Jeon et al., 2017** | **Loggia et al., 2015** | **Matsudaira et al., 2020** | **Nakatomi et al., 2014** | **Mueller et al., 2023** | **Torrado-Carvajal et al., 2021** | **f** | **$f_{sig}/f_{defined}$ (%)** | **CanlabAtlas 2024** |
|---|---|---|---|---|---|---|---|---|---|---|---|---|---|
| | **ROI** | **WholeBrain** | **WholeBrain** | **ROI** | **ROI** | **WholeBrain** | **WholeBrain** | **WholeBrain** | **ROI** | **WholeBrain** | | | |
| aMCC/mPFC $_{sig}$ | | 1 | 1 | | | | | Cingulate? | | | 3 | 60% | |

| | | | | | | | | | | | | | |
|---|---|---|---|---|---|---|---|---|---|---|---|---|---|
| aMCC/mPFC$_{defined}$ | | 1 | 1 | | Cingul-ate | | | 1 | 1 | | 5 | | |
| pgACC$_{sig}$ | | | 1 | | | | | | | | 1 | 50% | |
| pgACC$_{defined}$ | | | 1 | | Cingul-ate | | | | | | 2 | | |
| vlPFC$_{sig}$ | | 1 | | | | | | | | | 1 | 100% biased | |
| vlPFC$_{defined}$ | | 1 | | | | | | | | | 1 | | |
| dmPFC$_{sig}$ | | 1 | | | | | | | | | 1 | 100% biased | |
| dmPFC$_{defined}$ | | 1 | | | | | | | | | 1 | | |
| dlPFC$_{sig}$ | | 1 | 1 | | | | | | | | 2 | 100% biased | |
| dlPFC$_{defined}$ | | 1 | 1 | | | | | | | | 2 | | |
| vmPFC$_{sig}$ | | | | | | | 1 | | | | 1 | 100% biased | |
| vmPFC$_{defined}$ | | | | | | | 1 | | | | 1 | | |
| aINS/pINS$_{sig}$ | 1 (p) | 1 | | | | | 1 | | | | 3 | 75% | labels4: insula_posterior; label3: anterior_agranular_insula |
| aINS/pINS$_{defined}$ | 1 | 1 | | | 1 | | 1 | | | | 4 | | |
| M1 & Premotor$_{sig}$ | | 1 (M1) | | | | | 1 | | | | 2 | 67% | |
| M1 & Premotor$_{defined}$ | | 1 | | | Prece n-tral | | 1 | | | | 3 | | |
| Postcentral$_{sig}$ | | | | | | 1 | | | 1 | | 2 | 67% | |
| Postcentral$_{defined}$ | | | | | 1 | 1 | | | 1 | | 3 | | |
| S1$_{sig}$ | 1 (S1) | 1 (S1) | | | | 1 | | | | | 3 | 75% | Labels3: S1 |

| | | | | | | | | | | | | | |
|---|---|---|---|---|---|---|---|---|---|---|---|---|---|
| S1$_{defined}$ | 1 | 1 | | | 1 | 1 | | | | | 4 | | |
| PCC$_{sig}$ | | 1 | | | | | | | | | 1 | 100% biased | |
| PCC$_{defined}$ | | 1 | | | | | | | | | 1 | | |
| Precuneus$_{sig}$ | | 1 | | | | | | | 1 | | 2 | 100% biased | |
| Precuneus$_{defined}$ | | 1 | | | | | | | 1 | | 2 | | |
| Occipital$_{sig}$ | 1 (V1) | | | 1 | | | | | 1 | | 3 | 100% | Too broad |
| Occipital$_{defined}$ | 1 | | | 1 | | | | | 1 | | 3 | | |
| Parietal$_{sig}$ | | 1 (s) | | | | | | | 1 | | 2 | 100% biased | |
| Parietal$_{defined}$ | | 1 | | | | | | | 1 | | 2 | | |
| Supramarginal$_{sig}$ | | 1 | | | | | | | 1 | | 2 | 100% biased | |
| Supramarginal$_{defined}$ | | 1 | | | | | | | 1 | | 2 | | |
| Temporal$_{sig}$ | | | | | | | 1 (i, m, s) | | 1 | | 2 | 100% biased | |
| Temporal$_{defined}$ | | | | | | | 1 | | 1 | | 2 | | |
| Fusiform$_{sig}$ | | | | | | | 1 | | | | 1 | 100% biased | |
| Fusiform$_{defined}$ | | | | | | | 1 | | | | 1 | | |
| ParaHPC$_{sig}$ | | | | | | | 1 | | | | 1 | 100% biased | |
| ParaHPC$_{defined}$ | | | | | | | 1 | | | | 1 | | |
| Thalamus$_{sig}$ | 1 (left) | | | | 1 | 1 | 1 | 1 | | 1 | 6 | 86% | Labels4: Thal |
| Thalamus$_{defined}$ | 1 | | | | 1 | 1 | 1 | 1 | 1 | 1 | 7 | | |
| Putamen$_{sig}$ | | | | | 1 | | | | | | 1 | 100% biased | |
| Putamen$_{defined}$ | | | | | 1 | | | | | | 1 | | |

| | | | | | | | | | | | | |
|---|---|---|---|---|---|---|---|---|---|---|---|---|
| Caudate$_{sig}$ | | | | | 1 | | | | | | 1 | 100% biased |
| Caudate$_{defined}$ | | | | | 1 | | | | | | 1 | |
| Pallidum$_{sig}$ | | | | | | | | | | | 0 | 0% |
| Pallidum$_{defined}$ | | | | | 1 | | | | | | 1 | |
| Accumbens$_{sig}$ | | | | | 1 | | | | | | 1 | 100% biased |
| Accumbens$_{defined}$ | | | | | 1 | | | | | | 1 | |
| Amygdala$_{sig}$ | | | | | | | 1 | | | | 1 | 33% |
| Amygdala$_{defined}$ | | | | | 1 | | 1 | 1 | | | 3 | |
| Hipp$_{sig}$ | | | | | | | 1 | 1 | | | 2 | 67% |
| Hipp$_{defined}$ | | | | | | | 1 | 1 | 1 | | 3 | |
| Midbrain$_{sig}$ | | | | | | | | 1 | | | 1 | 100% biased |
| Midbrain$_{defined}$ | | | | | | | | 1 | | | 1 | |
| Pons$_{sig}$ | | | | | | | | 1 | | | 1 | 100% biased |
| Pons$_{defined}$ | | | | | | | | 1 | | | 1 | |
| Cerebellum$_{sig}$ | | | | | | | | | | | 0 | 0% |
| Cerebellum$_{defined}$ | | | | | | | | | 1 | | 1 | |

Supplementary Table 7: Brain regions showing significance in chronic pain/functional somatic syndromes TSPO studies. Selected regions of interest (ROIs) are highlighted. aMCC, anterior mid-cingulate cortex; pgACC, perigenual anterior cingulate cortex; dm/dl/vmPFC, dorsal medial/dorsal lateral/ventral medial prefrontal cortex; PCC, posterior cingulate cortex; aINS/pINS, anterior/posterior insula; ParaHPC, parahippocampal gyrus; Hipp, hippocampus.
